# Depression Measurement in Autistic Adults and Adults Who Did Not Meet Autism Criteria: Retrospective Psychometric Evaluation of the Patient Health Questionnaire-9 in Telehealth Psychotherapy

**DOI:** 10.64898/2026.11.24.26368910

**Authors:** Kelly Whaling, Lindsay Olson, Meghan Fraley, Hannah Jones, Julia Heinly, Micah Mazurek, Rachel Loftin

## Abstract

**Background:** The Patient Health Questionnaire-9 (PHQ-9) is widely used to measure depressive symptoms, but its performance in autistic adults remains understudied. In autism-focused telehealth care, it is unclear whether PHQ-9 scores can be interpreted similarly for adults who meet autism criteria and those who do not, or across demographic subgroups of autistic adults.

**Objective:** This study evaluated PHQ-9 reliability, internal structure, and associations with anxiety to inform score interpretation in psychotherapy with autistic adults. Total-score comparability and item 9 endorsement were compared between adults who met autism criteria and adults assessed in the same service who did not. An exploratory aim was to assess score comparability across subgroups of diagnosed adults.

**Methods:** This retrospective psychometric study used the first eligible check-in with paired PHQ-9 and Generalized Anxiety Disorder-7 (GAD-7) scores in 2 nonoverlapping cohorts. The original cohort included 3,471 clients diagnosed with autism and 484 who completed the same service’s assessment without meeting diagnostic criteria. The replication cohort included 7,406 and 1,194 clients, respectively. Ordinal factor models assessed structure and whether item responses were comparable across groups at the same severity. Graded response models compared expected totals at matched latent severity across all 56 sets of 3 anchor items from items 1–8. Logistic models compared item 9 endorsement (response ≥1) at matched Patient Health Questionnaire-8 (PHQ-8) scores.

**Results:** Across anchor sets, diagnosed-minus-comparison differences in expected PHQ-9 totals, averaged over latent severity, ranged from −0.46 to 0.35 points in the original cohort and −0.47 to 0.49 points in replication. Model-specific 95% interval endpoints spanned −0.80 to 0.59 and −0.68 to 0.63 points, respectively. The largest upper bounds for the average absolute difference on the 0–27 scale were 0.502 and 0.524 points, respectively. Cronbach’s alpha ranged from 0.84 to 0.86. The correlated two-factor model fit better than the one-factor model, though absolute fit remained limited. PHQ-9 and GAD-7 correlations ranged from 0.69 to 0.74. PHQ-9 totals discriminated weakly against recorded depressive-disorder codes (area under the curve 0.60–0.65). In replication, scalar invariance was rejected in the primary comparison, with item-level differences of 0.10 points or less, and in 6 of 7 subgroup contrasts. At PHQ-8=5, the odds ratios for item 9 endorsement were 1.42 (95% CI 0.77–2.61) in the original cohort and 0.66 (95% CI 0.49–0.88) in replication. The joint between-cohort test yielded χ²(2)=5.78, P=.06.

**Conclusions:** The PHQ-9 total appears useful as a group-level summary of depressive symptoms at this service. Average modeled differences were small relative to within-group variation. Item 9 estimates at PHQ-8=5 pointed in opposite directions across cohorts. Comparability with nonautistic adults, diagnostic cutoffs, sensitivity to change, and applicability to other settings and to adults with higher support or different communication needs require further study.

## Introduction

Depression and suicidality are common in autistic adults [1,2]. A meta-analysis of 36 studies of autistic and possibly autistic people without co-occurring intellectual disability estimated pooled prevalences of 34.2% for suicidal ideation and 24.3% for suicide attempts and behaviors [3]. The Patient Health Questionnaire-9 (PHQ-9) is widely used to assess depressive symptoms and inform clinical care [4]. Sleep disruption, fatigue, and difficulty concentrating are symptoms assessed by the PHQ-9 that can also occur in the context of autism or autistic burnout [5,6]. How respondents understand and answer particular items may also depend on characteristics associated with autism like the literal interpretation of wording, interoceptive differences, or difficulty identifying and describing emotions (alexithymia) [7–11]. These considerations motivate evaluating how the PHQ-9 performs in autistic adults and whether its items function similarly across subgroups defined by characteristics like gender or age.

Emerging research on depression measurement in autistic youth and adults [11,12] provides a foundation for evaluating how the PHQ-9 performs in routine telehealth psychotherapy. A feasibility study examined agreement between screening and diagnostic instruments in autistic adults [13]. In community samples, a single PHQ-9 total score summarized depressive symptoms in autistic adults even though the items reflected two symptom domains [14]. Among university students, most items functioned similarly for self-reported autistic and self-reported nonautistic respondents; the exception was item 9, which asks about thoughts of death or self-harm [15]. Work with the Beck Depression Inventory-II in autistic adults suggests that item-level differences may have little effect on expected total scores [16], but whether the same holds for the PHQ-9 is unknown. If item-level differences offset one another, total scores may look comparable even when responses to particular symptoms differ; if they do not, two people with similar depression could fall on opposite sides of a screening cutoff. Conclusions about item bias also depend on which items are assumed to function equivalently, so a finding is more credible when it holds across alternative assumptions.

Elevated rates of depression and suicidality among autistic adults make it important to evaluate the tools used to monitor depressive symptoms in the settings where they receive care. Telehealth use in mental health care has grown [17,18], and autistic adults have reported positive experiences with remote care [19–23]. Because symptom questionnaires are routinely administered electronically in telehealth [24–27], autistic adults may complete the PHQ-9 repeatedly over the course of treatment, with scores used to track changes in depressive symptoms. Large general-population studies have found similar PHQ-9 scores across administration modes [28]. To our knowledge, however, no studies have evaluated the psychometric performance of the electronically administered PHQ-9 among autistic adults receiving routine telehealth psychotherapy. We addressed this gap in an autism-focused telehealth service where adults who met autism criteria and adults who did not completed the PHQ-9 as part of the same routine care workflow.

The aim of this study was to determine whether adults at the same estimated level of depression severity had similar expected PHQ-9 total scores regardless of autism assessment outcome. An exploratory aim was to examine whether PHQ-9 items and total scores functioned similarly across subgroups of clients diagnosed with autism. Additional analyses examined how sensitive the estimated group difference was to the choice of anchor items, evaluated the PHQ-9’s internal structure and reliability, and assessed associations with anxiety and wellbeing. Item 9 was examined separately because it showed noninvariance in prior work with autistic adults, and because suicidal ideation and suicide attempts are common in this population [1–3]. The analysis tested whether clients with the same PHQ-8 score differed in their likelihood of endorsing thoughts of death or self-harm.

## Methods

### Study Design and Setting

This retrospective psychometric study used deidentified electronic health record data from a national telehealth service providing autism assessments and neurodivergent-affirming psychotherapy. Licensed psychologists conducted structured autism assessments [29]. Licensed master’s-level clinicians delivered psychotherapy using the service’s autism-adapted care model [30], a flexible cognitive behavioral approach incorporating skills from dialectical behavior therapy and acceptance and commitment therapy. Clinicians used explicit, concrete language and adapted the pace and delivery to clients’ sensory needs and communication preferences.

In service-wide records of completed appointments, clients who received assessments had addresses in all 50 US states and the District of Columbia. Clients who participated in psychotherapy had addresses in 38 states. At the first completed appointment, 92.2% of assessment clients and 96.2% of psychotherapy clients used in-network insurance. Self-pay accounted for 6.2% and 2.8%, respectively. Clients in the original and replication cohorts had addresses in 28 and 35 US states, respectively.

The PHQ-9 and the Generalized Anxiety Disorder-7 (GAD-7) were administered together electronically as part of routine care. Completion was encouraged and optional. Before the October 2025 rollout of the measurement-informed care workflow, a check-in was requested before each psychotherapy appointment. After the rollout, a check-in was requested at intake and approximately monthly if none had been completed in the preceding 4 weeks. The original cohort spans both schedules. The replication cohort was observed under the later schedule. The baseline analytic file included one PHQ-9 administration per client, defined as the first available PHQ-9 during the observation window. A separate descriptive item 9 follow-up used the second eligible paired administration when available.

### Positionality

The author team included clinicians and researchers with autistic, attention-deficit/hyperactivity disorder (ADHD), AuDHD (co-occurring autism and ADHD), otherwise neurodivergent, disabled, and neurotypical perspectives. The research question was shaped by clinical and community concerns about whether standard depression measures function adequately in autism-adapted care while also recognizing that standard measures may not fully capture autistic experiences of depression. The organization also maintains clinical and research advisory structures that include autism specialists and autistic advocates or community members who provide input on research priorities and designs. These perspectives informed the framing of the study, but the present analysis was not community-based research.

### Participants

The original analytic cohort included clients whose first eligible record was dated May 1, 2025, through March 31, 2026. The replication cohort included clients whose first eligible record was dated April 1, 2026, through October 31, 2026; no client was in both cohorts. An eligible record was a completed paired PHQ-9 and GAD-7 submission linked to a psychotherapy appointment. The two groups in the assessment-outcome analyses were clients diagnosed with autism through the service’s structured assessment and clients who completed the same in-service assessment and did not meet autism diagnostic criteria. Table S1 shows how clients were distributed across all categories. Among clients with a qualifying in-service assessment outcome, 3,471 of 3,955 (87.8%) in the original cohort and 7,406 of 8,600 (86.1%) in the replication cohort met autism criteria.

### Measures

Depressive symptoms were measured with the PHQ-9 [4]. The PHQ-9 includes nine items rated from 0 to 3 for the preceding 2 weeks, with total scores ranging from 0 to 27. Higher scores indicate greater depressive symptom severity. Conventional severity bands were used descriptively: minimal (0-4), mild (5-9), moderate (10-14), moderately severe (15-19), and severe (20-27) [4]. Anxiety symptoms were measured with the GAD-7 [31]. The GAD-7 includes seven items rated from 0 to 3, with total scores ranging from 0 to 21. Higher scores indicate greater anxiety symptom severity. Conventional severity bands were used descriptively: minimal (0-4), mild (5-9), moderate (10-14), and severe (15-21) [31]. Two single-item routine-care ratings were also collected: stress and emotional functioning (observed range 1–7) and overall wellbeing (observed range 0–10). Higher scores indicated better functioning or wellbeing.

### Data Analytic Plan

This psychometric evaluation combined classical test theory, factor analysis, and item response theory. Classical test theory was used to estimate internal consistency and the standard error of measurement (SEM). Factor analysis was used to examine the structure of depressive symptoms, estimate general-factor saturation, assess the distinction between depression and anxiety, and test measurement invariance by evaluating whether item parameters could be held equal across groups. Item response theory was used to assess measurement precision across severity, check local dependence, screen for differential item functioning (DIF), and estimate whether item-level differences accumulated into differences in expected PHQ-9 total scores at matched severity (differential test functioning). Because these comparisons require placing both groups on a common scale, a subset of items (anchors) was constrained to have equal parameters across groups, and the remaining items were free to differ. Item 9 endorsement was also compared between groups at matched observed PHQ-8 scores using logistic regression. Welch 95% confidence intervals were calculated for unstandardized mean differences. Standardized mean differences were expressed as Hedges g, calculated using the pooled standard deviation and a small-sample correction, with approximate normal-theory 95% confidence intervals. P values were rounded from unrounded output and are reported to 2 decimal places in the main text (3 when P<.01) and to 3 decimal places in Multimedia Appendix 1.

The study was not preregistered. After the initial analyses, a later, nonoverlapping cohort was added as a replication sample to assess whether the main findings held across cohorts. Because expected total-score differences can depend on which items serve as anchors, the range across all 56 possible three-item anchor sets from items 1–8 was adopted after the initial analyses as the main summary (see Expected Total Scores Across Anchor Sets). No equivalence margin was prespecified, so these comparisons are descriptive and are not formal equivalence tests. Magnitude was interpreted against two reference values chosen post hoc: the alpha-based SEM and 1 point, the smallest possible difference between two observed PHQ-9 totals. Subgroup analyses within the diagnosed group, partial invariance and subsampling checks, and additional item 9 and alternative-model analyses were exploratory (Multimedia Appendix 1, Table S8). Additional analytic specifications and decision rules are detailed in Multimedia Appendix 1. Reporting was informed by the Consensus-based Standards for the Selection of Health Measurement Instruments (COSMIN) reporting guideline [32].

### Reliability and Structure

Cronbach’s alpha is reported descriptively by assessment group and cohort. Under COSMIN, internal consistency is rated indeterminate when evidence of unidimensionality is insufficient [33]. The SEM estimates the typical size of measurement error in an observed score. The alpha-based SEM was calculated as the SD at the first eligible check-in multiplied by √(1 − alpha).

One-factor and correlated two-factor confirmatory factor analysis (CFA) models were fitted to the four-category items using polychoric correlations and weighted least squares mean-and variance-adjusted (WLSMV) estimation with theta parameterization [34]. Following Tibubos et al [35], items 1, 2, 6, 7, and 9 loaded on a cognitive-affective factor, and items 3, 4, 5, and 8 loaded on a somatic factor. Item assignments vary across published models [36–38]. Wald 95% confidence intervals for the PHQ-9 latent factor correlations were based on the standardized CFA estimates and their standard errors.

Models were evaluated for convergence, admissibility, loadings, factor correlations, and fit. Reported indices comprised the scaled chi-square statistic, scaled and robust versions of the comparative fit index (CFI), Tucker–Lewis index (TLI), and root mean square error of approximation (RMSEA), and one standardized root mean square residual (SRMR) per model and group. The published COSMIN criterion [33] and a separate RMSEA criterion of <0.06 were evaluated. The COSMIN criterion was met when CFI or TLI was >0.95, RMSEA was <0.06, or SRMR was <0.08. The separate criterion was applied regardless of the other fit indices [39,40]. Table 1 reports selected indices and Multimedia Appendix 1, Table S3 gives the full results. A sensitivity model estimated residual covariances for sleep with fatigue and concentration with psychomotor change.

**Table 1.** Reliability and factor structure of the PHQ-9 by cohort and assessment group.

| **Measure<sup>a</sup>** | **Original cohort** |  | **Replication cohort** |  |
|---|---|---|---|---|
|  | **Diagnosed<br>(n=3,471)** | **Assessed comparison<br>(n=484)** | **Diagnosed<br>(n=7,406)** | **Assessed comparison<br>(n=1,194)** |
| **Reliability** |  |  |  |  |
| Cronbach alpha | 0.857 | 0.862 | 0.847 | 0.838 |
| Alpha-based SEM, points<sup>b</sup> | 2.18 | 2.11 | 2.23 | 2.16 |
| GRM sum-score reliability | 0.861 | 0.868 | 0.849 | 0.843 |
| ωT, one-factor CFA<sup>c</sup> | 0.905 | 0.916 | 0.897 | 0.887 |
| ωT, two-factor CFA<sup>c</sup> | 0.912 | 0.923 | 0.905 | 0.895 |
| **General-factor saturation (Schmid–Leiman)** |  |  |  |  |
| ωT | 0.926 | 0.933 | 0.917 | 0.912 |
| ωH | 0.775 | 0.772 | 0.742 | 0.696 |
| ωH/ωT<sup>d</sup> | 0.838 | 0.828 | 0.809 | 0.763 |
| ECV | 0.690 | 0.689 | 0.656 | 0.598 |
| **One-factor CFA** |  |  |  |  |
| Scaled CFI | 0.955 | 0.955 | 0.944 | 0.930 |
| Scaled RMSEA | 0.114 | 0.119 | 0.120 | 0.127 |
| Robust CFI | 0.879 | 0.863 | 0.856 | 0.823 |
| Robust RMSEA | 0.146 | 0.166 | 0.154 | 0.166 |
| SRMR | 0.062 | 0.071 | 0.068 | 0.077 |
| **Two-factor CFA** |  |  |  |  |
| Latent factor correlation | 0.852 | 0.838 | 0.825 | 0.834 |
| Scaled CFI | 0.970 | 0.971 | 0.962 | 0.947 |
| Scaled RMSEA | 0.095 | 0.098 | 0.100 | 0.113 |
| Robust CFI | 0.917 | 0.909 | 0.905 | 0.867 |
| Robust RMSEA | 0.123 | 0.138 | 0.127 | 0.146 |
| SRMR | 0.053 | 0.059 | 0.057 | 0.070 |

<sup>a</sup>CFA, confirmatory factor analysis. CFI, comparative fit index. ECV, explained common variance. GRM, graded response model. PHQ-9, Patient Health Questionnaire-9. RMSEA, root mean square error of approximation. SEM, standard error of measurement. SRMR, standardized root mean square residual. ωH, omega hierarchical. ωT, omega total.
<sup>b</sup>Calculated as the PHQ-9 standard deviation at the first eligible check-in × √(1 − alpha).
<sup>c</sup>Estimated on the latent response scale.
<sup>d</sup>Calculated from unrounded ωH and ωT.

Omega total for the underlying continuous item responses (ωT) was estimated from the one- and two-factor CFA models. General-factor saturation describes how much of the variance in total scores reflects a depression dimension shared by all nine items. Higher values support using a total score. A Schmid–Leiman decomposition used minimum-residual extraction and oblimin rotation to estimate general-factor saturation without prespecified item assignments.

Three group factors were specified in both assessment groups in each cohort. Omega hierarchical (ωH), explained common variance (ECV), the percentage of uncontaminated correlations (PUC), and the proportion of reliable total-score variance attributable to the general factor (ωH/ωT) are reported descriptively. The ωH and ECV estimates were compared descriptively with reference values of 0.70 and 0.60, respectively, which apply when PUC is below 0.80 [41–43].

Unidimensional graded response models evaluated score precision. Conditional reliability and test information were examined across latent severity. Overall item response theory (IRT) model fit was assessed with the limited-information C2 statistic and its RMSEA.

Because residual associations between items can inflate information and reliability estimates [44], Yen’s Q3 was computed from each group’s graded response model, and mean-centered Q3 (each Q3 minus the group’s mean across all 36 item pairs) above 0.20 was flagged descriptively in both cohorts. Original-cohort parametric bootstrap reference values were obtained by simulating and refitting 500 datasets per group and are reported in Multimedia Appendix 1, Table S7 [45].

Pearson correlations quantified associations of PHQ-9 totals with GAD-7 totals and with the two routine-care ratings. Joint ordinal CFAs compared one distress factor with separate depression and anxiety factors. The heterotrait–monotrait ratio (HTMT) was calculated from polychoric correlations. Percentile 95% confidence intervals were based on 2,000 client-level bootstrap resamples attempted per group, using those that converged (Multimedia Appendix 1, Table S9). The point estimates and upper limits were compared descriptively with 0.85 and 0.90, respectively [46].

### Invariance and Differential Item Functioning

Observed PHQ-9 totals ranged from 0 to 27. For the 56-anchor comparison, item 9 responses 2 and 3 were collapsed in both cohorts, yielding a model score range of 0–26. Cohort-specific analyses collapsed adjacent categories when a group-by-category cell contained fewer than 10 clients. Thus, the principal replication invariance model and the replication model using anchors selected by iterative purification retained all four item 9 categories.

Invariance across assessment groups was evaluated in the correlated two-factor ordinal CFA using polychoric correlations and Wu–Estabrook identification by adding equality constraints sequentially for thresholds, loadings, latent-response intercepts, and residual variances [47]. Thresholds mark transitions between response categories. Loadings reflect how strongly an item relates to the underlying factor. Intercepts account for systematic group differences in the underlying item response at the same severity, with equal intercepts indicating scalar invariance in this sequence. Invariance testing used the scaled-shifted chi-square difference test (P<.05). Changes in CFI, RMSEA, and SRMR were reported descriptively against benchmarks developed for continuous indicators (a CFI decrease of ≥0.010, an RMSEA increase of ≥0.015, and an SRMR increase of ≥0.030 at the loading step or ≥0.010 at the intercept and residual steps) [48,49].

Where scalar (intercept) invariance was rejected, intercept constraints were released in descending order of the score-test χ² until the difference test against the threshold-and-loading model no longer rejected equality. The practical impact of item noninvariance was evaluated in PHQ-9 points using expected-score differences [50–52]. Multimedia Appendix 1, Table S11 describes subgroup analyses and their cohort-specific testing procedures. Group differences in the somatic and cognitive-affective factor means were contrasted, using the partial scalar model in replication. Multimedia Appendix 1, Table S23 shows the domain-mean model with the diagnosed group as the reference.

To assess sensitivity to sample size, the scalar invariance test was repeated in 500 replication-cohort subsamples drawn at the original group sizes. The rejection proportion used an exact binomial 95% confidence interval.

In multigroup graded response models, the assessed comparison group’s latent mean and variance were fixed at 0 and 1. The other group’s latent mean and variance were estimated. Anchor items had equal discrimination and thresholds across groups. The alternative sets selected by iterative purification comprised three items with the least evidence of DIF among those unflagged throughout purification and in the final screen [53,54]. Likelihood-ratio DIF tests used Benjamini–Hochberg adjustment across the nine items within each contrast.

### Expected Total Scores Across Anchor Sets

Signed and unsigned differential test functioning were computed from the model-implied difference in expected totals at 61 latent severity values from −6 to 6 [50–52]. The signed statistic averages those differences and the unsigned statistic averages their absolute values. Weights were the mean posterior probabilities across clients. To calculate the upper bound on the original 0–27 scale, one group’s collapsed item 9 responses were assigned a score of 2 and the other group’s a score of 3, then the assignment was reversed. The larger absolute difference at each severity was averaged with the same weights. The bound uses fitted probabilities without sampling uncertainty. Each anchor set used 2,000 parameter draws for its 95% percentile interval. For the timing sensitivity and subgroup models, one-sided 95% upper confidence limits were obtained as the 95th percentile of the draw-specific averages of the largest absolute expected-score differences on the 0–27 scale. The alpha-based SEM provided descriptive context for magnitude.

All 56 three-item sets from items 1–8 were fitted in each cohort to quantify sensitivity to anchor choice. The reported range of 95% endpoints summarizes separate model-specific intervals and has no simultaneous coverage interpretation. The diagnosed group’s latent mean was also estimated under each anchor set in multigroup graded response models fitted by expectation-maximization with 61 quadrature points from −6 to 6. Standard errors came from the observed information matrix computed by Richardson extrapolation. The 95% intervals were Wald intervals. These models collapsed item 9 responses 2 and 3 in the original cohort and retained all four categories in replication. Fits were required to converge with a positive definite information matrix.

Targeted checks refitted models after excluding one item from each of two locally dependent pairs. In the original cohort, separate models used items 4, 6, and 7 individually as the sole anchor to assess how the estimated group difference depended on anchor choice (Multimedia Appendix 1, Table S17). The seven-item refits used 2,000 parameter draws for 95% percentile intervals. Primary expected-score estimates are reported with 95% confidence intervals. Three exploratory sensitivity analyses in the main text (combined cohorts, women only, and men versus women) and several supplementary estimates in Multimedia Appendix 1 are reported with central 90% percentile intervals, as originally specified. These intervals are narrower than the 95% intervals used elsewhere and should not be compared with them directly (Multimedia Appendix 1, Tables S19 and S20).

### Item 9

Item 9 was examined separately because endorsement has been associated with subsequent suicidal behavior [55] and prior research in autistic adults found loading noninvariance [15]. Any endorsement (responses 1–3) was compared with none using logistic regression with assessment group, PHQ-8 score (sum of items 1–8) centered at 10 [56], and their interaction. Odds ratios and risk differences were evaluated at PHQ-8 scores of 5, 10, and 15.

Wald intervals were computed for odds ratios. For each model, 4 million coefficient vectors were drawn from a multivariate normal distribution centered at the maximum likelihood estimates with their estimated covariance matrix. The 2.5th and 97.5th percentiles of the resulting risk differences formed the 95% confidence interval. For each group, adequate local support required at least five endorsers and five nonendorsers within two PHQ-8 points. The joint 2-df likelihood-ratio test used the cohort × group and cohort × group × PHQ-8 terms.

Exploratory item 9 checks examined raw endorsement, alternative logistic models, and item 9 loading contrasts from ordinal two-factor models. For the loading analysis, item 9 responses 2 and 3 were collapsed in both cohorts, and thresholds were constrained to equality across assessment groups. A Wald test compared the group odds ratios between cohorts at PHQ-8=5. The loading contrasts used delta-method Wald intervals. Repeat endorsement was examined among first-check-in endorsers with a second eligible check-in. The group-by-PHQ-8 interaction was expressed as the ratio of odds ratios per 5-point increase in PHQ-8.

### Comparisons Within Diagnosed Clients

Seven pairwise contrasts were examined among clients diagnosed with autism. They compared men with women, gender-diverse clients with women, clients younger than 35 years with those aged 35 years or older, Black with White clients, multiracial with White clients, Hispanic or Latine with non-Hispanic or Latine clients, and clients with versus without a recorded ADHD billing code among those with treatment-plan records. Benjamini–Hochberg adjustment was applied to the nine item-level DIF tests within each contrast, with Holm adjustment as a sensitivity check (Multimedia Appendix 1, Tables S11–S14 and S19). Both cohorts included omnibus tests for gender and race. The pairwise follow-up tests differed. In the original cohort, the joint test constrained thresholds, loadings, and intercepts. In the replication cohort, the adjacent-step test added intercept constraints to the threshold-and-loading model. Differences in which contrasts reached significance may partly reflect the different constraints tested.

### Sensitivity Analyses and Subgroup Analyses

Sensitivity analyses examined intake-linked first check-ins, assessment outcomes dated by the first eligible check-in, combined-cohort estimates under alternative anchor sets, and exclusion of unusual response patterns or all-zero profiles. An original-cohort analysis restricted to women assessed whether the expected-score difference persisted when both assessment groups were limited to women. The PHQ-9 association with recorded depressive-disorder codes was summarized using areas under the receiver operating characteristic curve (AUC). DeLong 95% confidence intervals for these areas were calculated.

### Software

Analyses used R 4.5.3 [57] and incorporated the following packages: lavaan 0.7-3 [58], semTools 0.5-8 [59], psych 2.6.9 [60], mirt 1.48 [61], and GPArotation 2026.8-2 [62]. Item 9 logistic models used stats::glm, with splines::ns for natural-spline terms. Firth sensitivity models used logistf 1.26.1 [63] with penalized profile-likelihood intervals. The coefficient draws for the risk-difference intervals used MASS::mvrnorm (MASS 7.3-65) [64]. Latent means across anchor sets were estimated using mirt::multipleGroup. The receiver operating characteristic analyses used pROC 1.19.0.1 [65].

## Ethical Considerations

The University of Virginia Institutional Review Board for Health Sciences Research and HIPAA Privacy Board (Study Tracking No. 303492) determined that this secondary analysis of existing deidentified data met exemption criteria. No new participant data were collected. Clients were offered the option to opt out of research use of their deidentified data without penalty or effect on their clinical care. Participants received no compensation. The study followed the ethical principles of the Belmont Report [66] and the Declaration of Helsinki [67]. Records of clients who opted out of research use were excluded before the research team received the data. The research extract did not include a count of those exclusions, so the number of clients who opted out during the study period was unavailable to the research team.

## Results

### Participants and Score Distributions

The original cohort included 5,077 psychotherapy clients with an eligible check-in, and the nonoverlapping replication cohort included 10,579 (Figure 1). The primary comparison included 3,955 clients in the original cohort (3,471 diagnosed with autism through the service and 484 who completed the service’s assessment without receiving a diagnosis) and 8,600 in the replication cohort (7,406 diagnosed and 1,194 assessed without a diagnosis). Together, the two cohorts contributed 12,555 unique clients to the primary comparisons. The other 1,122 clients in the original cohort and 1,979 in the replication cohort lacked a recorded result from the service’s autism assessment that would place them in either comparison group. Their assessment categories are shown in Figure 1 and Multimedia Appendix 1, Table S1.

**Figure 1.**
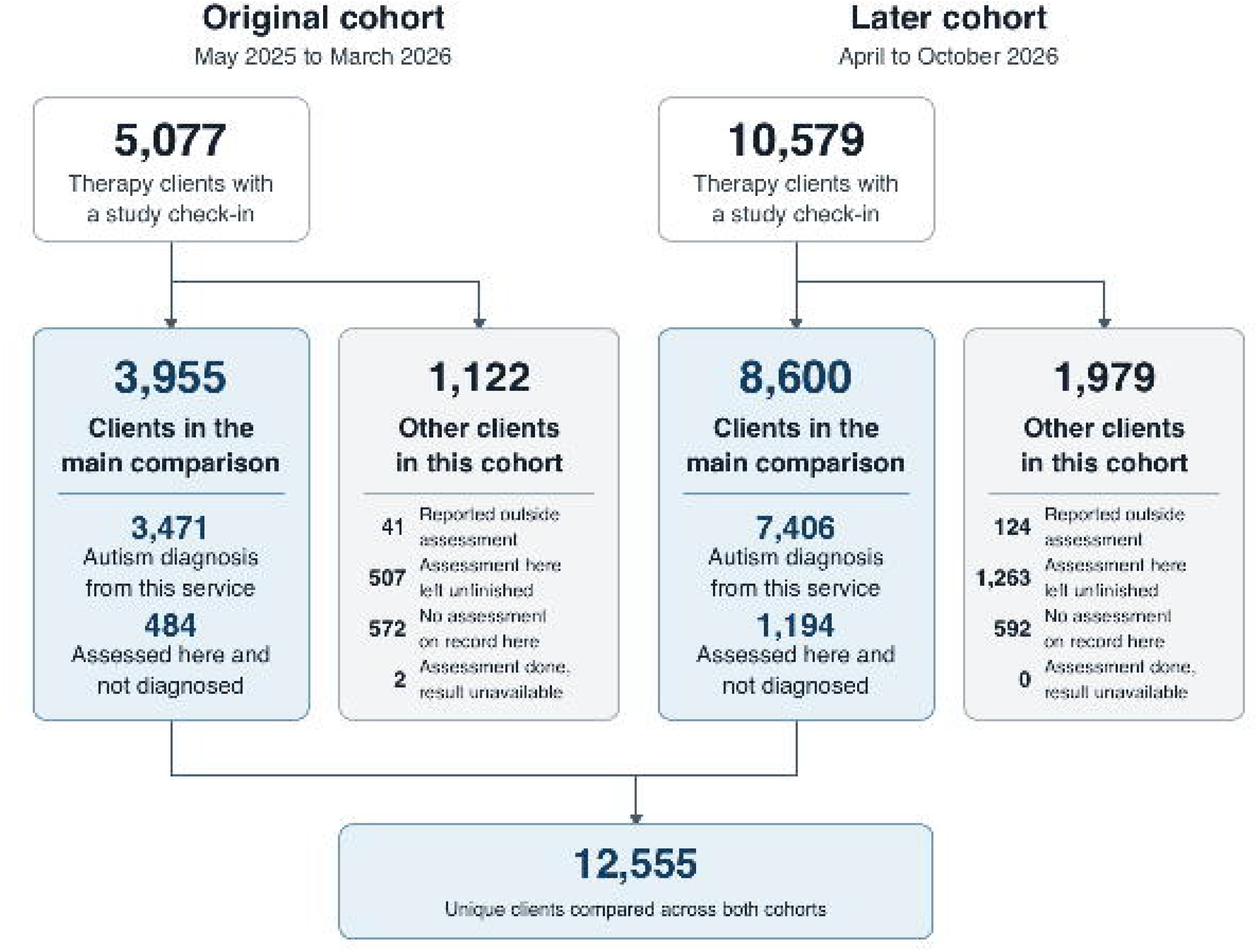
Client flow into the primary assessment comparison in the original and nonoverlapping replication cohorts.

Across cohorts, women accounted for 62.4%–66.8% of diagnosed clients and 71.0%–72.3% of assessed comparison clients. Black clients accounted for 9.5%–11.3% and 13.2%–17.6% of these groups, respectively, and Hispanic or Latine clients accounted for 15.5%–16.0% and 17.8%–18.1%, respectively (Multimedia Appendix 1, Table S1).

The first eligible check-in was linked to therapy intake for 2,509 of 3,955 clients (63.4%) in the original primary comparison and 8,022 of 8,600 (93.3%) in replication. Through March 31 and October 31, 2026, respectively, 2,128 (53.8%) and 4,544 (52.8%) had a second eligible check-in. Among those clients, intervals between first and second check-ins were a median of 33 days (IQR 29–42) and 31 days (IQR 29–37), respectively.

Observed mean PHQ-9 scores were higher among diagnosed clients than among assessed comparison clients in both cohorts. In the original cohort, the mean PHQ-9 score at the first eligible check-in was 9.39 (SD 5.77) in diagnosed clients and 8.44 (SD 5.67) in assessed comparison clients (difference 0.95 points, 95% CI 0.41 to 1.49; Hedges g=0.17, 95% CI 0.07 to 0.26). Scores of 10 or higher occurred in 1,503/3,471 diagnosed clients (43.3%) and 175/484 assessed comparison clients (36.2%). In the replication cohort, the means were 9.70 (SD 5.69) and 8.90 (SD 5.37) (difference 0.80 points, 95% CI 0.47 to 1.13; Hedges g=0.14, 95% CI 0.08 to 0.20). Scores of 10 or higher occurred in 3,361/7,406 (45.4%) and 476/1,194 (39.9%). Demographics, score distributions, and item response distributions are in Multimedia Appendix 1 (Tables S1, S2, and S25, respectively).

### Reliability and Structure

Cronbach’s alpha was between 0.84 and 0.86 in all four groups (0.857 and 0.862 in the original diagnosed and assessed comparison groups; 0.847 and 0.838 in the replication cohort). Alpha-based SEMs were 2.18 and 2.11 PHQ-9 points in the original cohort and 2.23 and 2.16 in the replication cohort (Table 1).

The correlated two-factor CFA provided a better fit than the one-factor CFA in both cohorts. Absolute fit remained limited. Scaled RMSEA was 0.095 to 0.113 for the two-factor model and 0.114 to 0.127 for the one-factor model. Two-factor SRMR was 0.053 to 0.070 and scaled CFI was 0.947 to 0.971. Robust two-factor indices were less favorable (RMSEA 0.123 to 0.146 and CFI 0.867 to 0.917). Model fit improved across groups when residual covariances were estimated for sleep–fatigue and concentration–psychomotor change, with scaled RMSEA of 0.078 to 0.092 (Table 1 and Multimedia Appendix 1, Tables S3–S4 and Extended Results). Both models met the published COSMIN criterion on SRMR in every group. Scaled and robust RMSEA values exceeded 0.06.

The cognitive-affective and somatic factors were strongly correlated: 0.85 (95% CI 0.83 to 0.87) in original diagnosed and 0.84 (0.79 to 0.89) in original assessed comparison clients, and 0.83 (0.81 to 0.84) and 0.83 (0.80 to 0.87) in the replication cohort.

In the Schmid–Leiman decomposition, the general factor accounted for 84% and 83% of model-estimated reliable total-score variance in diagnosed and assessed comparison clients in the original cohort and 81% and 76% in the replication cohort. Across the four groups, ωH ranged from 0.696 to 0.775 and ECV from 0.598 to 0.690. PUC was 0.556 under the two-domain CFA assignment and 0.722 under the dominant-loading assignment in the Schmid–Leiman solution (Multimedia Appendix 1, Table S4). The replication assessed comparison group had ωH=0.696 and ECV=0.598, both below the descriptive reference values. The confirmatory bifactor model for the original diagnosed group did not yield a reproducible solution across starting values (Multimedia Appendix 1, Tables S3–S4).

In the original cohort, conditional reliability was at least 0.80 across approximately 3–22 model-implied PHQ-9 points. Information was greatest at moderate to high latent severity (Multimedia Appendix 1, Figure S1). The graded response models showed limited fit, with C2 RMSEA values of 0.103–0.117 across the four groups (Multimedia Appendix 1, Tables S5–S6).

Sleep with fatigue and concentration with psychomotor change remained associated after adjustment for latent severity. Mean-centered Q3 for sleep–fatigue ranged from 0.28 to 0.30 across all four groups. Concentration–psychomotor change exceeded the 0.20 flag in three of four groups (0.24–0.29). In the original cohort, the maximum mean-centered Q3 was 0.28 for sleep–fatigue in diagnosed clients and 0.32 for depressed mood–worthlessness in assessed comparison clients. The corresponding 95th percentiles of the bootstrap maximum were 0.10 and 0.18 (Multimedia Appendix 1, Table S7).

PHQ-9–GAD-7 Pearson correlations ranged from 0.69 to 0.74. The joint 16-item CFA fit better with separate depression and anxiety factors than with one general distress factor. Latent correlations ranged from 0.76 to 0.82, and HTMT estimates from 0.80 to 0.84, with upper 95% limits of 0.81 to 0.88. All 2,000 bootstrap resamples converged in three groups, and 1,995 of 2,000 converged in the original assessed comparison group. PHQ-9 totals were inversely correlated with the stress and emotional functioning rating (r=−0.60 to −0.50) and overall wellbeing (r=−0.50 to −0.37) across cohort-by-assessment groups (Multimedia Appendix 1, Table S9).

### Invariance and Differential Item Functioning

The invariance analysis examined whether the PHQ-9 measurement parameters could be held equal across the diagnosed and assessed comparison groups. The configural model provided the starting point and showed limited fit in both cohorts (Multimedia Appendix 1, Table S10). Equality constraints were then added in stages to test thresholds, loadings, intercepts, and residual variances (Table 2).

**Table 2.** Sequential measurement invariance tests of the correlated two-factor Patient Health Questionnaire-9 model across autism assessment groups.

| **Added constraint** | **Scaled and shifted Δχ<sup>2a</sup>** | **df<sup>b</sup>** | **P value** | **ΔCFI<sup>c</sup>** | **ΔRMSEA<sup>d</sup>** | **ΔSRMR<sup>e</sup>** |
|---|---|---|---|---|---|---|
| **Original cohort (diagnosed n=3,471, assessed comparison n=484)** |  |  |  |  |  |  |
| Thresholds | 9.27 | 8<sup>f</sup> | .32 | −0.0010 | −0.0051 | 0.0000 |
| Loadings (metric) | 8.20 | 7 | .31 | 0.0035 | −0.0097 | 0.0001 |
| Intercepts (scalar) | 3.65 | 7 | .82 | 0.0040 | −0.0096 | 0.0000 |
| Residuals (strict) | 20.60 | 9 | .01 | 0.0027 | −0.0078 | 0.0009 |
| **Replication cohort (diagnosed n=7,406, assessed comparison n=1,194)** |  |  |  |  |  |  |
| Thresholds | 8.39 | 9 | .50 | −0.0017 | −0.0058 | 0.0000 |
| Loadings (metric) | 8.15 | 7 | .32 | 0.0054 | −0.0113 | 0.0000 |
| Intercepts (scalar) | 22.03 | 7 | .003 | 0.0047 | −0.0096 | 0.0002 |
| Residuals (strict) | 29.90 | 9 | <.001 | 0.0022 | −0.0068 | 0.0006 |

<sup>a</sup>Scaled and shifted chi-square difference. Each model was compared with the preceding model. The threshold model was compared with the configural baseline. Absolute configural fit is reported in Multimedia Appendix 1, Table S10.
<sup>b</sup>df: degrees of freedom for the difference test.
<sup>c</sup>CFI: comparative fit index. Changes are the constrained model minus the preceding model. A negative ΔCFI or a positive ΔRMSEA or ΔSRMR indicates worse fit. Changes in CFI and RMSEA use scaled indices.
<sup>d</sup>RMSEA: root mean square error of approximation.
<sup>e</sup>SRMR: standardized root mean square residual.
<sup>f</sup>The original cohort combined item 9 responses 2 and 3, so the threshold test had 8 degrees of freedom. The replication cohort retained all 4 categories, yielding 9 degrees of freedom.

The scaled-shifted chi-square difference tests did not detect threshold or loading differences in either cohort. At the intercept step, scalar invariance was not rejected in the original cohort (Δχ²(7)=3.65, P=.82) and was rejected in the replication cohort (Δχ²(7)=22.03, P=.003). A partial invariance model then tested whether the remaining intercepts could be held equal after freeing those for depressed mood and anhedonia. The remaining constraints were not rejected relative to the threshold-and-loading model (Δχ²(5)=10.03, P=.07). Strict invariance was rejected in both cohorts when residual variances were constrained (original P=.01, replication P<.001).

Three sensitivity checks probed the replication intercept result. It persisted when item 9 responses 2 and 3 were pooled to match the original cohort’s coding (Δχ²(7)=21.87, P=.003). Allowing residual covariances between sleep and fatigue and between concentration and psychomotor change left the intercept decision unchanged in both cohorts. To examine sensitivity to sample size, the replication cohort was also subsampled to the original cohort’s group sizes. The intercept constraints were rejected in 193 of 500 subsamples (38.6%, exact binomial 95% CI 34.3%–43.0%).

Changes in approximate fit were small throughout the sequence. CFI decreased only at the threshold step, by 0.0010 in the original cohort and 0.0017 in replication. The largest SRMR increases were 0.0009 and 0.0006, respectively, and RMSEA decreased at every step. No change reached a descriptive fit-change cutoff, including at the replication intercept step (Table 2 and Multimedia Appendix 1, Table S10).

Item-level DIF analyses examined whether the assessment groups were expected to score differently on individual PHQ-9 items at the same modeled depression severity. After Benjamini–Hochberg adjustment, no items were flagged in the original cohort. Three were flagged in replication: depressed mood, fatigue, and psychomotor change. Their estimated differences (diagnosed minus assessed comparison) were −0.08, 0.04, and 0.10 PHQ-9 points, respectively. With Holm adjustment, only psychomotor change (item 8) was flagged in the replication cohort, and no items were flagged in the original cohort (Multimedia Appendix 1, Table S12). For context, the estimated difference for psychomotor change in the original cohort was 0.08 points, close to the replication estimate. Anhedonia, whose intercept was freed in the replication partial-invariance model, had an estimated item-score difference of −0.07 points in that cohort. Item 9 was not flagged in either cohort (Multimedia Appendix 1, Tables S12–S13).

### Expected Total Scores Across Anchor Sets

The anchor sensitivity analysis examined whether the diagnosed and assessed comparison groups had different expected PHQ-9 totals at the same latent depression severity, and how much those differences depended on the items used to link the groups. All 56 three-item anchor sets from items 1–8 were evaluated in each cohort. Item 9 responses 2 and 3 were combined in both cohorts for this comparison (fitted score range 0–26). Positive differences indicate higher expected totals in the diagnosed group.

Averaged over latent severity, signed differences stayed within about half a PHQ-9 point across all 56 sets. Estimates ranged from −0.46 to 0.35 points in the original cohort and from −0.47 to 0.49 points in the replication cohort (Figure 2). Estimates were negative for 22 of 56 sets in the original cohort and 18 of 56 in the replication cohort. The same anchor set yielded estimates with the same sign in both cohorts for 44 of 56 sets, and the paired estimates were positively correlated (Spearman ρ=0.74).

**Figure 2.**
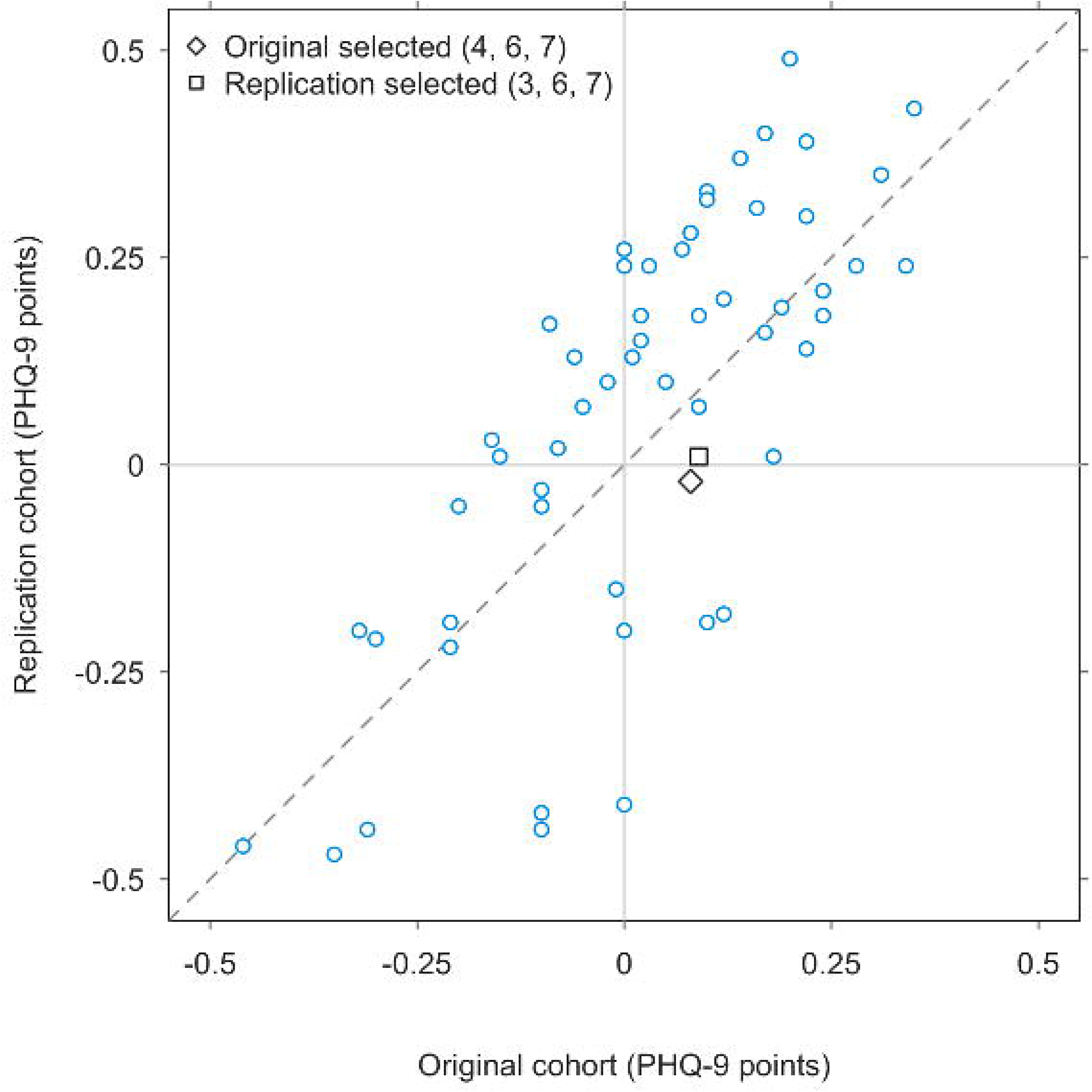
Signed expected PHQ-9 total-score differences (diagnosed minus assessed comparison) in the original and replication cohorts across 56 three-item anchor sets. Note: Each point pairs the severity-averaged estimates from the same anchor set in both cohorts. The dashed line marks equal estimates. The diamond and square mark the anchor sets selected by iterative purification in the original and replication cohorts, respectively. Item 9 responses 2 and 3 were combined in both cohorts (fitted score range 0–26).

Across the model-specific 95% intervals, endpoints ranged from −0.80 to 0.59 points in the original cohort and from −0.68 to 0.63 points in the replication cohort. Each interval was conditional on its anchor set, and the endpoint ranges summarize those intervals descriptively. Accounting for the combined item 9 response category, the largest upper bounds for the average absolute difference on the original 0–27 scale were 0.502 points in the original cohort and 0.524 points in the replication cohort (Multimedia Appendix 1, Table S16). For context, the alpha-based SEM for individual scores ranged from 2.11 to 2.23 points (Table 1).

To examine whether anchor choice mattered near the screening cutoff, the diagnosed group’s expected total was evaluated at the severity where the assessed comparison group’s expected total was 10. Across anchor sets, the diagnosed group’s expected total ranged from 9.57 to 10.51 in the original cohort and from 9.52 to 10.53 in the replication cohort. Anchor choice also affected the estimated size of the latent mean difference. The diagnosed group had the higher estimated latent mean under all 56 sets in each cohort, with differences of 0.118 to 0.263 SDs in the original cohort and 0.046 to 0.250 SDs in the replication cohort, in units of the assessed comparison group’s latent SD. Model-specific 95% Wald intervals excluded zero for 56 and 49 sets, respectively. The cutoff and latent mean analyses used cohort-specific item 9 coding, combining responses 2 and 3 in the original cohort and retaining all four categories in the replication cohort. Under the anchor sets selected by iterative purification, the severity-averaged signed difference was 0.077 points (95% CI −0.207 to 0.325) in the original cohort and 0.002 points (95% CI −0.187 to 0.201) in the replication cohort (Figure 3 and Multimedia Appendix 1, Table S14). The corresponding latent mean differences were 0.175 SDs (95% CI 0.062 to 0.289) and 0.144 SDs (95% CI 0.066 to 0.223).

**Figure 3.**
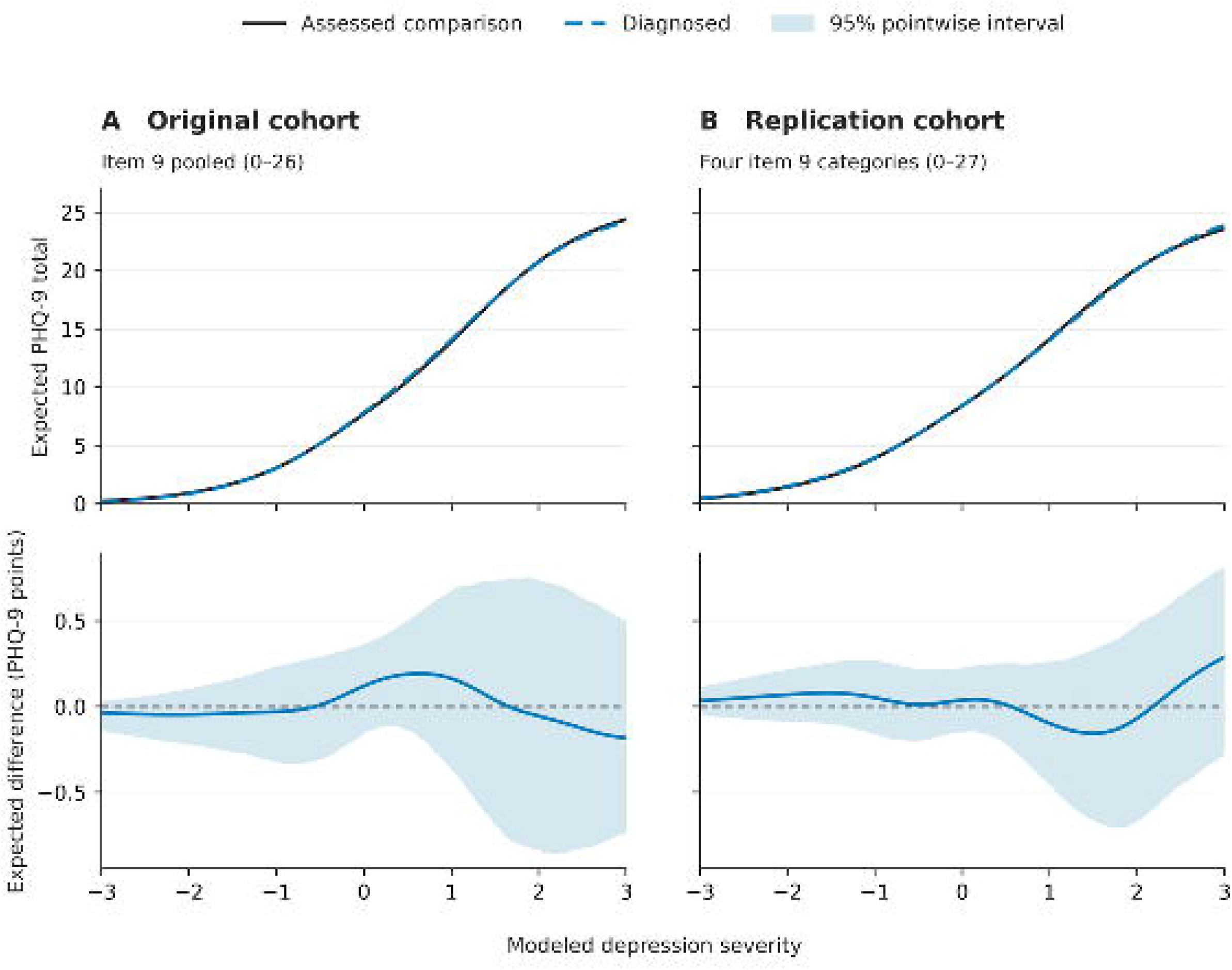
Expected PHQ-9 totals by assessment group (top) and diagnosed-minus-assessed comparison differences (bottom) across latent severity values from −3 to 3. Note: Shading shows pointwise 95% intervals conditional on the anchors selected by iterative purification. The original cohort used items 4, 6, and 7 as anchors and combined item 9 responses 2 and 3 (fitted range 0–26). The replication cohort used items 3, 6, and 7 as anchors and retained all four item 9 categories (fitted range 0–27).

Two sensitivity analyses examined how the signed difference changed when the model used a single anchor or excluded items from locally dependent pairs. In the original cohort, each of the three selected anchors was tested individually. Item 7 yielded the largest difference in absolute value (+0.65 points), above the range across the 56 three-item sets. After fatigue and psychomotor change were excluded, one from each locally dependent pair, the signed difference on the seven-item scale was −0.027 points (95% CI −0.248 to 0.163) in the original cohort and −0.135 points (95% CI −0.284 to 0.002) in the replication cohort (Multimedia Appendix 1, Tables S14–S17). Anchor sets with more somatic items tended to yield lower signed differences (Multimedia Appendix 1, Table S16). To examine group differences by symptom domain, the two-factor CFA estimated the difference between the somatic and cognitive-affective latent mean contrasts. The diagnosed-minus-comparison contrast was larger for the somatic factor by 0.076 SDs (95% CI −0.013 to 0.165) in the original cohort and 0.056 SDs (95% CI −0.018 to 0.129) in the replication cohort’s partial scalar model, in units of the assessed comparison group’s latent SD (Multimedia Appendix 1, Extended Results).

### Item 9

At the first eligible check-in, 531 of 3,471 diagnosed clients (15.3%) and 58 of 484 assessed comparison clients (12.0%) endorsed item 9 (response 1–3) in the original cohort. In the replication cohort, the corresponding counts were 1,172 of 7,406 (15.8%) and 171 of 1,194 (14.3%).

At the same PHQ-8 score, the direction of the fitted group difference varied by cohort (Table 3). At scores of 5 and 10, estimated endorsement was higher among diagnosed clients in the original cohort and lower among diagnosed clients in the replication cohort. At a score of 15, estimated endorsement was higher among diagnosed clients in both cohorts. Of the six risk differences in Table 3, only the replication estimate at a score of 5 had a 95% CI that excluded zero (−2.2 percentage points, 95% CI −4.2 to −0.6). The joint test assessed whether the level or slope of the group contrast differed between cohorts and yielded χ²(2)=5.78 (P=.06). At each reported score, both groups had at least five endorsers and five nonendorsers within two PHQ-8 points (Multimedia Appendix 1, Table S26). The smallest endorsement count was 6 of 182 assessed comparison clients with scores of 3–7 in the original cohort.

**Table 3.** Model-estimated endorsement of Patient Health Questionnaire-9 item 9 (thoughts of death or self-harm) by autism assessment group at selected Patient Health Questionnaire-8 scores.

| **PHQ-8<sup>a</sup> score** | **Diagnosed, %<sup>b</sup>** | **Assessed comparison, %<sup>b</sup>** | **Risk difference, percentage points (95% CI<sup>c</sup>)<sup>d</sup>** | **Odds ratio (95% CI)<sup>e</sup>** |
|---|---|---|---|---|
| **Original cohort (diagnosed n=3,471, assessed comparison n=484)** |  |  |  |  |
| 5 | 4.24 | 3.02 | 1.2 (−1.1 to 2.8) | 1.42 (0.77 to 2.61) |
| 10 | 11.86 | 9.79 | 2.1 (−1.8 to 5.1) | 1.24 (0.85 to 1.81) |
| 15 | 29.01 | 27.45 | 1.6 (−5.9 to 8.2) | 1.08 (0.76 to 1.54) |
| **Replication cohort (diagnosed n=7,406, assessed comparison n=1,194)** |  |  |  |  |
| 5 | 4.51 | 6.72 | −2.2 (−4.2 to −0.6) | 0.66 (0.49 to 0.88) |
| 10 | 12.10 | 13.92 | −1.8 (−4.2 to 0.4) | 0.85 (0.70 to 1.04) |
| 15 | 28.65 | 26.63 | 2.0 (−2.4 to 6.1) | 1.11 (0.89 to 1.37) |

<sup>a</sup>PHQ-8: Patient Health Questionnaire-8 (sum of items 1–8).
<sup>b</sup>Model-estimated percentage endorsing item 9 (responses 1–3 vs 0). Cohort-specific logistic models included assessment group, PHQ-8 score centered at 10, and their interaction.
<sup>c</sup>CI: confidence interval.
<sup>d</sup>Diagnosed minus assessed comparison. The 95% CIs are the 2.5th and 97.5th percentiles of risk differences from 4 million simulated coefficient draws per model. Differences were calculated before rounding.
<sup>e</sup>Odds in the diagnosed group relative to the assessed comparison group. The 95% CIs are Wald intervals.

After the fitted contrasts at a PHQ-8 score of 5 were observed to point in opposite directions, a post hoc test compared the cohort-specific odds ratios at that score. The odds ratios for diagnosed versus assessed comparison clients were 1.42 (95% CI 0.77 to 2.61) in the original cohort and 0.66 (95% CI 0.49 to 0.88) in the replication cohort. The original-to-replication ratio was 2.17 (95% CI 1.10 to 4.27, unadjusted Wald P=.03). Among original-cohort clients with a score of exactly 5, endorsement was lower in diagnosed clients (7 of 234, 3.0%) than in assessed comparison clients (2 of 32, 6.3%). These counts describe clients at a single score. The fitted models used the full observed score range, and the quadratic, natural-spline, and Firth models preserved the direction of the fitted contrast in each cohort. For each 5-point increase in PHQ-8 score, the diagnosed-versus-comparison odds ratio was multiplied by 0.87 (95% CI 0.63 to 1.20) in the original cohort and by 1.30 (95% CI 1.09 to 1.55) in the replication cohort (Multimedia Appendix 1, Table S18).

The ordinal two-factor models examined whether the item 9 loading differed between assessment groups. The standardized loading was lower in diagnosed clients in the original cohort (diagnosed minus assessed comparison, −0.078) and higher in the replication cohort (0.136). The replication-minus-original difference between these contrasts was 0.215 (95% CI 0.088 to 0.341, unadjusted P<.001). On the unstandardized scale, the corresponding estimate was 0.359, with a 95% CI that included zero (−0.101 to 0.818, unadjusted P=.13).

Among clients who endorsed item 9 at the first eligible check-in and had a second eligible check-in, 153 of 279 diagnosed clients (54.8%) and 17 of 29 assessed comparison clients (58.6%) endorsed it again in the original cohort. In the replication cohort, 304 of 646 diagnosed clients (47.1%) and 36 of 85 assessed comparison clients (42.4%) endorsed it again (Multimedia Appendix 1, Table S18, Panel G).

### Sensitivity Analyses and Subgroup Analyses

To limit variation in therapy exposure before measurement, the invariance analysis was repeated among the 2,509 original-cohort clients whose first eligible check-in was linked to therapy intake (63.4%). The scalar invariance test yielded P=.79, compared with P=.82 in the primary analysis (Multimedia Appendix 1, Table S11). To assess whether the results depended on using assessment reports issued after the first check-in, the analysis was restricted to 2,272 original-cohort clients with reports dated on or before that check-in. The signed expected-score difference was −0.02 points (95% CI −0.47 to 0.37). The one-sided 95% upper confidence limit for the average absolute difference was 0.96 points and incorporated sampling uncertainty (Multimedia Appendix 1, Table S20).

To examine whether the expected-score difference was sensitive to the choice of anchors in a common sample, the two cohorts were combined. The signed difference was 0.01 points (90% CI −0.12 to 0.13) with the original cohort’s anchors (items 4, 6, and 7) and 0.03 points (90% CI −0.10 to 0.16) with the replication cohort’s anchors (items 3, 6, and 7) (Multimedia Appendix 1, Table S20).

Among women in the original cohort, the expected PHQ-9 total-score difference was −0.054 points (diagnosed minus assessed comparison, 90% CI −0.347 to 0.204). The upper bound on the average absolute difference on the original 0–27 scale was 0.428 points (Multimedia Appendix 1, Table S20).

The observed original-cohort mean difference was 0.99 points after excluding 165 profiles flagged for repeated nonzero responses, multivariate outliers, or poor item-response model fit, and 0.88 points after separately excluding 108 all-zero profiles. The primary comparison yielded a difference of 0.95 points (Multimedia Appendix 1, Table S22). Under broader group definitions, signed expected-score differences relative to the assessed comparison group were 0.09 points (95% CI −0.19 to 0.35) after adding clients with prior outside assessments to the diagnosed group and 0.02 points (95% CI −0.23 to 0.24) after also including clients with incomplete assessments or no in-service assessment record (Multimedia Appendix 1, Table S14).

Among 9,974 clients with treatment-plan records, the AUC for PHQ-9 totals against recorded depressive-disorder codes ranged from 0.598 (DeLong 95% CI 0.555 to 0.641) in the replication assessed comparison group to 0.650 (95% CI 0.593 to 0.707) in the original assessed comparison group. The corresponding estimates among diagnosed clients were 0.628 (95% CI 0.606 to 0.649) in the original cohort and 0.647 (95% CI 0.631 to 0.662) in the replication cohort (Multimedia Appendix 1, Table S24).

Seven exploratory subgroup contrasts were examined among diagnosed clients. In the original cohort, scalar invariance was rejected in the omnibus gender test and in the age and recorded ADHD-code comparisons (all P<.001). The omnibus race test yielded P=.09. For the original-cohort Black-versus-White and multiracial-versus-White pairwise comparisons, interpretation was limited to descriptive fit changes because model nesting could not be confirmed. In the replication cohort, scalar invariance was rejected in six of seven pairwise contrasts, with multiracial versus White clients as the exception. In all six rejected contrasts, freeing one to three intercepts yielded P values above .05 for the remaining constraints. The closest result to the threshold was the Hispanic or Latine comparison (P=.05; .0503 unrounded) (Multimedia Appendix 1, Table S11 and Extended Results).

Three-anchor models were available for six contrasts in each cohort. Men versus women was the exception. Across the six three-anchor contrasts, the one-sided 95% upper confidence limits for the average absolute difference ranged from 0.32 to 0.93 points in the original cohort and from 0.28 to 0.51 points in the replication cohort. For men versus women, the signed estimate using items 1 and 7 together was −0.38 points (90% CI −0.61 to −0.15) in the original cohort and −0.31 points (90% CI −0.47 to −0.15) in the replication cohort. Using item 1 alone yielded negative estimates in both cohorts, and using item 7 alone yielded positive estimates in both. The direction also varied with anchor choice for Black versus White clients (Multimedia Appendix 1, Tables S14 and S19).

Across the seven final DIF screens, Benjamini–Hochberg adjustment within each nine-item comparison flagged 10 of 63 item tests in the original cohort and 12 of 63 in the replication cohort. In the original cohort, Holm adjustment within each subgroup contrast flagged 7 of 63 item tests (Multimedia Appendix 1, Table S12).

## Discussion

### Summary of Principal Findings

Across two cohorts, expected PHQ-9 totals at the same modeled depression severity differed by about half a point or less on average between clients diagnosed with autism and clients assessed without a diagnosis. Both groups received psychotherapy through the same autism-focused telehealth service. The magnitude of the average signed difference remained small across all 56 three-item anchor sets in each cohort, although its direction varied with anchor choice. The largest upper bound for the average absolute difference on the original 0–27 scale, accounting for the collapsed item 9 categories, was 0.524 points. Item-level differences and rejection of scalar invariance in the replication cohort coexisted with small differences in expected totals, a pattern also reported for the Beck Depression Inventory-II in autistic and nonautistic adults [16]. No equivalence margin was prespecified; the differences are described as small relative to the SEM (2.11 to 2.23 points) and to a 1-point difference in observed totals.

### Reliability, Structure, and Precision

The cognitive-affective and somatic factors were highly correlated (0.83 to 0.85), and the general factor accounted for 76% to 84% of reliable total-score variance in the exploratory Schmid–Leiman models. This supports interpreting the PHQ-9 total as a group-level summary of depressive symptoms in this sample, consistent with findings in autistic adults in the community [14] and in a large psychotherapy sample [68]. The better fit of the two-factor model also supports distinguishing cognitive-affective and somatic symptom clusters when describing the structure of item responses [37].

General-factor estimates were weakest in the replication assessed comparison group, where omega hierarchical was 0.696 and explained common variance was 0.598, just below the respective descriptive benchmarks of 0.70 and 0.60 [42,43]. In the original cohort, omega hierarchical was similar in diagnosed and assessed comparison clients (0.775 and 0.772). The two-factor models met the published COSMIN criterion on the standardized root mean square residual, while the root mean square error of approximation exceeded 0.06 [33]. Residual associations between sleep and fatigue and between concentration and psychomotor change suggest symptom clustering beyond the modeled factors and may inflate model-based estimates of precision. The confirmatory bifactor solution for the original diagnosed group was unstable across starting values, so the general-factor interpretation rests primarily on the exploratory Schmid–Leiman estimates.

The PHQ-9 and GAD-7 item responses were better described by two correlated dimensions than by a single dimension. This supports interpreting them as measures of related aspects of distress.

### Comparability by Assessment Outcome

The threshold and loading tests provided little evidence of differences between assessment groups in either cohort. The scalar invariance test yielded P=.82 in the original cohort and rejected equality in the replication cohort (P=.003). After freeing the depressed mood and anhedonia intercepts in the replication cohort, the comparison with the threshold-and-loading model yielded P=.07. Strict invariance was rejected in both cohorts. When the replication cohort was subsampled at the original cohort’s group sizes, 193 of 500 subsamples (38.6%) rejected scalar invariance, suggesting that sample size may partly explain the different test results (Multimedia Appendix 1, Extended Results).

The rejection of scalar invariance in the replication cohort was statistically detectable but practically negligible. No fit index reached a conventional cutoff for noninvariance at the intercept step. The items involved differed by 0.10 points or less each, and these differences partly canceled, leaving expected totals within about half a point. With 8,600 clients, the chi-square difference test could detect differences too small to affect interpretation of the total score.

The expected-score analyses showed how item-level differences translated to the total. In the replication cohort, lower expected scores for depressed mood and anhedonia were partly offset by higher expected scores for fatigue and psychomotor change (Multimedia Appendix 1, Table S13). Depressed mood was the only item identified by both the intercept-release analysis and the item response model’s differential item functioning screen. The two analyses assessed different model parameters.

Observed mean PHQ-9 scores were 0.95 points higher among diagnosed clients in the original cohort and 0.80 points higher in the replication cohort. Both gaps exceeded the magnitude of the corresponding average signed expected-score differences under every three-item anchor set examined. The estimated latent mean was higher in the diagnosed group under every set, although seven of 56 model-specific 95% intervals included zero in the replication cohort. This pattern is consistent with higher modeled depressive symptom severity among diagnosed clients and depends on the assumptions used to link the groups. Iterative purification can select items with undetected differential item functioning [54], and a response tendency shared across items could be absorbed into the latent mean difference.

At the modeled severity where the assessed comparison group’s expected total was 10, the diagnosed group’s expected total ranged from approximately 9.5 to 10.5 across anchor sets. The range crossed 10 in both cohorts. These comparisons describe expected scores at a common modeled severity. Individual screening classifications require separate evaluation. Within diagnosed clients, several exploratory subgroup contrasts showed item noninvariance, and the direction of the signed total-score difference depended on anchor choice for men versus women and Black versus White clients.

The single-group graded response models showed poor absolute fit (C2 RMSEA 0.103–0.117) and residual dependence for the sleep–fatigue and concentration–psychomotor pairs. In the two-factor CFA, allowing residual covariances for both pairs led to the same intercept-equality decisions in both cohorts. After dropping fatigue and psychomotor change, the estimated differences on the seven-item scale were −0.027 points (95% CI −0.248 to 0.163) in the original cohort and −0.135 points (95% CI −0.284 to 0.002) in the replication cohort. The expected-score estimates remain conditional on the fitted graded response models.

### Item 9

At a PHQ-8 score of 5, fitted item 9 endorsement was lower among diagnosed clients in the replication cohort. The original-cohort estimate pointed in the opposite direction and was imprecise. The joint test of whether the fitted contrasts differed between cohorts yielded P=.06. The post hoc comparison at a score of 5 yielded an unadjusted Wald P=.03 and addressed the narrower question of how the contrasts differed at that score. In the original cohort, 7 of 234 diagnosed clients (3.0%) and 2 of 32 assessed comparison clients (6.3%) endorsed item 9 at exactly that score. This raw contrast pointed in the opposite direction from the fitted original-cohort contrast, and the small number of endorsements limits its interpretation (Multimedia Appendix 1, Table S18).

Robeson et al [15] reported item 9 loading noninvariance in a university sample. The graded response model screens in this study did not flag item 9 in either cohort. In the separate ordinal loading analysis, the between-cohort contrast was clearer on the standardized scale, with the unstandardized interval including zero. The evidence for item 9 noninvariance by assessment outcome in this service remains limited.

Roughly half of first-check-in endorsers who contributed a second eligible questionnaire endorsed item 9 again. These observations describe the subset who completed a second questionnaire during active psychotherapy and may reflect persistence of the experiences captured by item 9 as well as changes in symptoms or reporting (Multimedia Appendix 1, Table S18, Panel G).

Suicidality measures developed in general populations have limited validation in autistic adults, and standard questions may be interpreted differently [69,70]. The Suicidal Behaviours Questionnaire–Autism Spectrum Conditions was developed with autistic adults for research and may help structure conversations about suicidal thoughts and support needs [71]. The present analyses addressed item endorsement and comparability by assessment outcome. Suicide-risk prediction and screening accuracy were outside the study’s scope.

### Association With Recorded Depressive-Disorder Codes

PHQ-9 totals distinguished poorly between clients with and without a recorded depressive-disorder code (AUC 0.598–0.650). These values fell below the COSMIN benchmark of 0.70 for comparisons against a reference standard [33]. The treatment-plan codes included remission diagnoses and could have been recorded before or after the PHQ-9. Clinicians could also see PHQ-9 scores when assigning codes. The analysis therefore describes how well scores distinguished recorded code status. Criterion validity and the diagnostic performance of PHQ-9 cutoffs in this setting remain uncertain.

### Clinical Implications

These findings extend prior research on the PHQ-9 in autistic adults [14] to an autism-focused telehealth psychotherapy service. Factor structure and reliability were examined separately among clients who received an autism diagnosis through the service and clients who completed the same assessment without receiving a diagnosis. Score functioning was compared between these groups and, in exploratory analyses, across subgroups of diagnosed clients. The findings support using the PHQ-9 total to summarize depressive symptoms at the group level in this setting. The better-fitting two-factor model met the COSMIN criterion for structural validity based on the standardized root mean square residual [33], while its RMSEA exceeded the present study’s conservative cutoff of 0.06. When observed group differences are small, their interpretation should take into account the size of the modeled score differences and how their direction changed with the items used to link the groups.

The psychometric findings for item 9 were mixed. Robeson et al [15] reported a group difference in the item 9 factor loading in a university sample. In the present study, the omnibus loading tests provided little evidence of loading differences, while an exploratory item-specific analysis suggested that the standardized loading contrast varied between cohorts. Estimated differences in endorsement at matched PHQ-8 scores ranged from 1.2 to 2.2 percentage points in absolute magnitude and pointed in opposite directions across cohorts at scores of 5 and 10. This pattern provides little basis for interpreting an endorsement differently by autism assessment outcome. In either group, clinicians should review item 9 separately from the total and ask what the client meant by an endorsement, even when the overall score is low. Direct follow-up can clarify whether the client is reporting a wish to be dead, thoughts of suicide, or thoughts of self-harm.

For interpreting change in an individual’s score, single-administration reliability estimates implied a 95% minimal detectable change of 5.8 to 6.2 points using coefficient alpha and 5.7 to 6.1 points using graded response model reliability, assuming independent errors with equal variance. These estimates were similar to the pooled value of 5.72 points reported by Wang et al [72] in samples that excluded people seeking mental health care. The estimates provide a descriptive reference for measurement error. Establishing a reliable-change threshold for this service would require repeat assessments when symptoms are expected to remain stable. Responsiveness and the smallest change that matters to clients require separate evidence.

### Strengths, Limitations, and Future Directions

The primary comparison was based on structured, in-service autism assessments conducted by licensed psychologists with specialized training in diagnosing autism in adults [29]. Both groups sought an autism assessment and received psychotherapy through the same service, reducing differences in referral context and care setting. The primary comparisons included 12,555 clients across two nonoverlapping cohorts, with the replication cohort more than twice the size of the original. Expressing modeled differences in PHQ-9 points made their magnitude interpretable alongside the statistical tests. Evaluating all 56 three-item anchor sets in each cohort showed how estimates changed with the choice of items used to link the groups. Subsampling the replication cohort at the original group sizes suggested that sample size may partly explain why the scalar invariance tests yielded different results. Neurodivergent members of the author team and advisory groups that included autistic community members informed the research question and framing.

The study was not preregistered. The replication cohort and the main summary across anchor sets were added after the initial analyses, and the subgroup and follow-up item 9 analyses were exploratory. Future studies should prespecify the primary comparisons, the analysis of anchor choice, and the item 9 follow-up tests before examining the data.

Both cohorts came from one telehealth service and included adults who sought an autism assessment and received psychotherapy there. Clients assessed without a diagnosis may share autistic traits or other clinical characteristics with diagnosed clients [7]. Classification error could reduce the ability to detect item-level differences between assessment groups. In simulations, detection declined under some scenarios that introduced errors into the assessment labels (Multimedia Appendix 1, Table S21). The assessment groups differed in gender and race composition, and the analyses did not adjust for those differences. Some items also functioned differently by gender among diagnosed clients. Differences in group composition could therefore contribute to or obscure item-level differences by assessment outcome. In the original cohort, the estimated total-score difference remained close to zero when the analysis was limited to women (−0.054 points, 90% CI −0.347 to 0.204). A corresponding analysis was unavailable in the replication cohort. Three exploratory sensitivity analyses used 90% rather than 95% intervals, which understates their uncertainty relative to the other estimates. Replication across services and care models should document how diagnoses were determined, characterize both groups clinically, and examine whether results hold after accounting for demographic and clinical differences.

Questionnaire completion was optional. The 5,077 clients in the original analytic cohort represented approximately 66.5% of the 7,633 unique clients with at least one completed, nondeleted psychotherapy appointment during the same window. The aggregate appointment count was not screened for test accounts or reconciled against all study eligibility criteria. The extract also excluded clients who opted out of research use. The sample therefore reflects both who entered care and who contributed eligible data. Participation also required telehealth access and the ability to complete self-report measures, and autism support needs were unavailable in the extract. Future studies should document participation at each stage, examine who is represented in the analytic sample, and use accessible recruitment and assessment procedures to include autistic adults with intellectual disability, minimal or no speech, or higher support needs.

The factor and item response models showed misfit and residual item dependence, and the invariance tests were conditional on the fitted configural model. Model-specific intervals quantify sampling uncertainty for a given model and anchor set, the spread across anchor sets shows sensitivity to the linking items, and the 0.524-point bound quantifies uncertainty from scoring the collapsed item 9 categories separately from sampling uncertainty. Future analyses should test whether the expected-score results hold under models that better account for residual associations between items. A response tendency shared across all items could be absorbed into the estimated latent mean difference. Comparing self-reports with an independent source, such as an informant report or clinician-rated depression interview, could help investigate this possibility [73]. Cognitive interviews with autistic adults could address a related question that the psychometric analyses leave open: whether the items are understood as intended and capture depression as autistic adults experience it.

Assessment outcomes were sometimes recorded after the first eligible check-in. Restricting the original cohort to clients whose outcome was recorded by that check-in yielded a signed difference of −0.02 points (95% CI −0.47 to 0.37) and a one-sided 95% upper confidence limit of 0.96 points for the average absolute difference. That limit incorporates sampling uncertainty. The first eligible check-in was linked to therapy intake for 63.4% of original-cohort clients and 93.3% of replication-cohort clients, and the check-in schedule changed during the original observation window. Timing in care may therefore contribute to between-cohort differences, including those for item 9. The extract began at the first eligible check-in and contained only complete PHQ-9 and GAD-7 pairs. Future studies should retain the dates of therapy entry, autism assessment, and questionnaire administration, along with records of PHQ-9-only completion, so that timing and questionnaire availability can be examined directly.

Repeat check-ins occurred during active psychotherapy, when changes in scores could reflect symptom change or measurement error. Only one original-cohort pair and no replication-cohort pairs had first and second administrations 5–14 days apart, so short-interval test–retest reliability could not be estimated (Multimedia Appendix 1, Extended Results). Repeat administrations during a period of independently established symptom stability would allow direct estimation of test–retest reliability and minimal detectable change. Responsiveness and meaningful individual change should be evaluated separately against external indicators of clinical change and clients’ accounts of whether improvement mattered to them.

Item 9 was analyzed as any endorsement versus none, with clients matched on observed PHQ-8 scores. Clients with the same PHQ-8 total can differ in symptom profile and underlying severity. Such differences can increase false-positive findings in item comparisons when groups differ in estimated latent means, as they did here [74]. Follow-up tests used unadjusted P values, and unstandardized loadings were scaled against each cohort’s own reference group, limiting direct comparison. Future studies should prespecify the item 9 contrasts and their multiplicity adjustment, examine comparability on a common scale, and retain response-frequency information where counts permit. An independent assessment of suicidal thoughts and self-harm could clarify what an endorsement captures. Adequate enrollment of assessed comparison clients at low PHQ-8 scores would improve precision where counts were sparse.

The treatment-plan analysis used recorded depressive-disorder codes, including remission codes, without a timing restriction relative to the questionnaire. Clinicians could see the PHQ-9 scores, which may have strengthened the observed association. A structured depression interview conducted close to questionnaire completion by an interviewer blinded to the score would provide an independent criterion for evaluating diagnostic accuracy and cutoff performance (Multimedia Appendix 1, Table S24).

Within diagnosed clients, subgroup analyses examined one characteristic at a time. Some subgroups were small, and two original-cohort intercept tests were uninterpretable because model nesting could not be confirmed. Future studies should recruit enough clients to support those comparisons, verify model nesting before interpreting difference tests, and use intersectional analyses to examine whether item functioning varies across combinations of characteristics [75]. Future studies could also examine the PHQ-9-AASPIRE, an adaptation of the PHQ-9 developed for autistic adults, with the same psychometric approach [76].

## Conclusions

In a large evaluation study consisting of two cohorts within a national telehealth service, the PHQ-9 total score summarized a common depression dimension and measured depressive symptoms comparably in clients diagnosed with autism and clients assessed without a diagnosis. Scalar invariance was rejected in the larger replication cohort, but the item-level differences behind that result were 0.10 points or less each and had no practical effect on the total. Among diagnosed clients, scalar invariance was also rejected in most subgroup comparisons, but the effect on expected totals was again under 1 point; these exploratory results need replication. Consistent with earlier research [14], these findings support using the PHQ-9 total as a brief summary of self-reported depressive symptoms and a starting point for discussing them in outpatient psychotherapy with autistic adults.

Group differences in item 9 endorsement were small and varied in direction across cohorts. Because item 9 asks about thoughts of death or self-harm, clinicians should review the response separately and follow up directly on any endorsement, even when the total is low. Further research should examine how autistic adults interpret item 9, how well standard cutoffs perform, and how much change in a score is meaningful, and should include adults with intellectual disability, minimal or no speech, or higher support needs. Services can continue using the PHQ-9 while evaluating whether adaptations such as the PHQ-9-AASPIRE could complement or replace it in routine care. Autistic adults’ experiences of depression should guide both the evaluation of these measures and decisions about their use.

## Acknowledgments

The authors thank the clients whose deidentified routine-care data made this study possible and the clinicians and operations staff who supported routine outcome monitoring in care.

## Data Availability

The clinical data analyzed in this study are not publicly available because they were collected during routine mental health care. Although the analytic dataset was deidentified, public sharing could create privacy risks and would be inconsistent with the trust placed in the clinical service by therapy clients. Analysis code used to reproduce the analyses is available upon request to.

## Authors’ Contributions

KW contributed to conceptualization, methodology, formal analysis, investigation, data curation, visualization, project administration, and writing (original draft, review, and editing). LO contributed to conceptualization, methodology, investigation, clinical interpretation, and writing (original draft, review, and editing). MF and HJ each contributed to investigation, clinical interpretation, and writing (original draft, review, and editing). JH contributed to methodology, investigation, clinical interpretation, and writing (review and editing). MM contributed to methodology, supervision, clinical and scientific interpretation, and writing (review and editing). RL contributed to conceptualization, methodology, resources, project administration, supervision, clinical and scientific interpretation, and writing (review and editing). All authors reviewed and approved the final manuscript.

## Generative AI Disclosure

The authors used Grammarly (embedded in Microsoft Word and Google Docs) for spelling, grammar, and language checks. The authors also used enterprise workspaces of ChatGPT (OpenAI) and Claude (Anthropic), configured for organizational privacy and security, to assist with manuscript formatting tasks like converting references from APA author-date style to JMIR/AMA numbered format, renumbering in-text bracketed citations after edits to maintain accurate sequential order, and converting plain-text characters into proper mathematical and scientific notation (eg, converting “x” to the chi symbol, χ). Claude (Anthropic) was also used minimally during revisions to assist with basic code debugging (eg, identifying syntax errors when analysis scripts failed to run). Generative AI tools were not used to make scientific decisions, conduct statistical analyses, interpret results, verify references, or generate scientific content. The authors reviewed and edited all AI-assisted outputs and take full responsibility for the final content. No generative AI tool is listed as an author.

## Abbreviations

ADHD: attention-deficit/hyperactivity disorder
AUC: area under the receiver operating characteristic curve
AuDHD: co-occurring autism and ADHD
CFA: confirmatory factor analysis
CFI: comparative fit index
COSMIN: Consensus-based Standards for the Selection of Health Measurement Instruments
DIF: differential item functioning
ECV: explained common variance
GAD-7: Generalized Anxiety Disorder-7
GRM: graded response model
HTMT: heterotrait-monotrait ratio
IRT: item response theory
PHQ-8: Patient Health Questionnaire-8
PHQ-9: Patient Health Questionnaire-9
PUC: percentage of uncontaminated correlations
RMSEA: root mean square error of approximation
SEM: standard error of measurement
SRMR: standardized root mean square residual
TLI: Tucker-Lewis index
WLSMV: weighted least squares mean- and variance-adjusted
ωH: omega hierarchical
ωT: omega total

## Multimedia Appendices

Multimedia Appendix 1: Supplementary methods and results, including Tables S1–S26 and Figure S1.

