## Supplementary Materials for "Depression Measurement in Autistic Adults and Adults Who Did Not Meet Autism Criteria: Retrospective Psychometric Evaluation of the Patient Health Questionnaire-9 in Telehealth Psychotherapy"

### Multimedia Appendix 1

#### Supplementary Methods and Results

Index of supplementary tables and figure

| Main-text section | Appendix tables and figure |
|---|---|
| Participants and observed scores | S1 Participants, ascertainment, and check-in timing; S2 Score distributions and percentiles; S25 Item response distributions and polychoric correlations |
| Analysis plan | S8 Analysis aims and supporting results |
| Reliability and structure | S3 Model fit; S4 Reliability and dimensionality; S5 Graded response and Rasch results; S6 Item fit; S7 Local dependence; S9 Anxiety and wellbeing associations; Figure S1 Test information and standard error |
| Invariance and DIF | S10 Assessment-outcome invariance; S11 Subgroup and other invariance checks; S12 DIF screening; S13 Item-level expected-score effects |
| Expected total scores | S14 Integrated score differences; S15 Anchors, coding, and simulation draws; S16 All three-item anchor sets; S17 Targeted alternative anchors; S23 Domain factor means |
| Item 9 | S18 Item 9 analyses; S26 Item 9 local support counts |
| Sensitivity and subgroup analyses | S19 Men-versus-women anchor models; S20 Timing, composition, and coding; S21 Detection simulations; S22 Additional clinical comparisons; S24 Recorded depressive-disorder codes |

Supplementary methods and results for the PHQ-9 psychometric evaluation. Tables S1–S26 and Figure S1 cover the original and nonoverlapping replication cohorts. Model-specific response coding and interval levels are indicated. P values are reported to three decimal places where the unrounded value was available and to two decimal places otherwise; four decimals appear only where rounding would cross .05 (.0497 and .0503). The main text reports the same values to two decimal places when P≥.01. Software and package versions are reported in the main manuscript (Methods, Software).

Abbreviations

| Abbreviation | Definition |
|---|---|
| ADHD | attention-deficit/hyperactivity disorder |
| AIC | Akaike information criterion |
| AUC | area under the receiver operating characteristic curve |
| BH | Benjamini–Hochberg |
| BIC | Bayesian information criterion |
| C2 | limited-information C2 goodness-of-fit statistic |
| CFA | confirmatory factor analysis |
| CFI | comparative fit index |
| CI | confidence interval |

| Abbreviation | Definition |
|---|---|
| COSMIN | Consensus-based Standards for the Selection of Health Measurement Instruments |
| DIF | differential item functioning |
| dMACS | differential mean and covariance structure effect size |
| EAP | expected a posteriori |
| ECV | explained common variance |
| GAD-7 | Generalized Anxiety Disorder-7 |
| GRM | graded response model |
| HTMT | heterotrait–monotrait ratio |
| ICC | intraclass correlation coefficient |
| IQR | interquartile range |
| IRT | item response theory |
| PHQ-8 | Patient Health Questionnaire-8 |
| PHQ-9 | Patient Health Questionnaire-9 |
| PUC | percentage of uncontaminated correlations |
| RMSEA | root mean square error of approximation |
| SEM | standard error of measurement |
| SL | Schmid–Leiman |
| SRMR | standardized root mean square residual |
| TLI | Tucker–Lewis index |
| ULSMV | unweighted least squares mean- and variance-adjusted |
| WLSMV | weighted least squares mean- and variance-adjusted |
| ωH | omega hierarchical |
| ωT | omega total |

### Supplementary methods

#### Sample definitions, timing, and missing information

Assessment groups used the available in-service outcome regardless of timing. The assessment-before-baseline check restricted clients to a dated diagnostic report on or before their first questionnaire; intake-only analyses used questionnaires linked to therapy intake. Broader cohorts added prior outside assessment and then assessment-not-completed and no-assessment-record categories. A self-identified-only subgroup was unavailable as a separate category. ADHD and anxiety-code comparisons required available treatment-plan records; missing codes were never assigned to code absence. Clinical means and percentiles used original 0–27 scoring.

**Structure, precision, and local dependence**

Ordinal CFA used WLSMV and theta parameterization unless designated ULSMV. The conventional bifactor model was evaluated at a default start and 100 randomized starts in each original-cohort assessment group. The exploratory Schmid–Leiman decomposition specified three group factors, minimum-residual extraction and oblimin rotation, with cross-loadings estimated; the three-group-factor specification was carried forward to the replication cohort. Confirmatory PUC applies to the confirmatory model only. Graded response item parameters, information and S-$X^2$ fit supplement the main analyses. Yen Q3 used EAP trait estimates; adjusted Q3 subtracts the mean across all 36 unique item pairs. Values >.20 were descriptive flags in both cohorts. Parametric-bootstrap critical values were generated only in the original cohort, using 500 simulated and refitted datasets per group [1,2].

**Invariance, DIF, and category handling**

Adjacent-model tests used the ordered-categorical sequence specified in the main text. Approximate-fit flags were scaled CFI decrease ≥.010, RMSEA increase ≥.015, and SRMR increase ≥.030 at loadings or ≥.010 at intercepts [3]. Item- and total-score effects were estimated regardless of the equality-test decision. Eligible anchors excluded item 9 and every item ever flagged or finally flagged; the three eligible items with the largest final P values defined a screening-selected sensitivity model for the assessment-group comparison. The ordinal CFA category maps and the GRM category maps are reported separately in Table S15 because the two model families used different maps. Original clinical totals always retain their original response categories.

Expected-score differences were integrated over the combined empirical posterior marginal severity distribution. Signed effects permit cancellation across severity and items;

unsigned whole-test effects prevent cancellation across severity. Unsigned item effects instead take the absolute difference before integration at item level. All parameter-draw intervals share the model-and-anchor condition stated in the main Methods; the common-coding enumeration supplies point estimates and model-specific 95% intervals. Draw counts, category maps, C2 fit and independently seeded endpoint gaps are in Table S15. Original-scale envelopes bound the possible contributions of merged response categories.

**Additional sensitivities and numerical checks**

Exploratory partial-invariance checks used score tests and sequentially released up to three constraints, targeting the largest standardized score-test statistic. The main ordinal sequences were repeated with ULSMV, 100 balanced subsamples, and 1,000 label permutations. Response screens considered nonzero straightlining, all-maximum profiles, Mahalanobis upper-tail P<.001, and person-fit flags; sensitivity exclusions separated flagged from all-zero profiles. Principal models included all eligible clients.

Each cohort contributed 56 fitted models based on three-item anchor sets drawn from items 1–8. For the main comparison, both cohorts pooled item 9 responses 2 and 3 (fitted range 0–26, Table S16, Panel C). Panel A shows the corresponding fits using each cohort's original coding (0–26 in the original cohort and 0–27 in the replication cohort). The focal latent mean and variance were free. The 0–27 upper bounds were calculated from fitted category probabilities by scoring the pooled response as 2 in one group and 3 in the other, reversing the assignment, then averaging the larger absolute difference across severity. The common-coding run used 2,000 accepted draws per model and central 95% percentiles (2.5th and 97.5th). Each model used a fixed seed of 20261120 plus its anchor-set index in the original cohort or 20261220 plus the index in the replication cohort. The native-coding enumeration reports point estimates.

For men versus women, the joint model constrained items 1 and 7 and freed all other item parameters, with women's trait mean and variance fixed at 0 and 1. Estimation used 61 quadrature points and two batches of 4,000 parameter draws. Each likelihood-ratio comparison freed one anchor's four parameters, leaving the other fixed. AIC and Bayesian information criterion (BIC) were reported alongside the tests. The two-anchor model departed from the primary three-anchor rule.

**Item 9 and association follow-ups**

Binary item 9 models used any endorsement versus none in every cohort, PHQ-8 centered at 10, group and their interaction. Pooled risk differences were standardized to the cohort mix. Logistic maximum-likelihood estimates used coefficient covariance for odds ratios and coefficient simulation for risk-difference intervals. The binary models were fitted with stats::glm in R 4.5.3. For each model, MASS::mvrnorm (MASS 7.3-65) generated two batches of 2 million coefficient vectors from a multivariate normal distribution centered at the maximum likelihood estimates with their estimated covariance matrix. The batches were combined before calculating the 2.5th and 97.5th percentiles of the resulting risk differences. The batch seeds were 20261124 and 20261125 for the original cohort, 20261126 and 20261127 for replication, and 20261128 and 20261129 for the pooled model. Additional ordinal two-factor models estimated group-specific item 9 loadings under threshold-invariant identification; standardized differences used delta-method uncertainty. Continuous-indicator dMACS analyses and cumulative-logit checks for 0/1/2/3 and 0/1/2+ coding addressed separate parameter questions. Proportional-odds diagnostics were evaluated by term; follow-ups allowed group, PHQ-8 and interaction effects to vary by threshold. Natural-spline models used splines::ns. The Firth models in Panel E used logistf 1.26.1 with penalized profile-likelihood intervals.

HTMT used original-category polychoric matrices without smoothing and 2,000 attempted client-level bootstrap samples within each assessment group and cohort. Finite ratios from nonpositive-definite matrices were included and counted separately from failures. Percentile intervals were two-sided 95%; .85 was the descriptive reference. Joint ordinal CFAs compared a single distress factor with separate correlated depression and anxiety factors, using the same specified model in both cohorts. Wellbeing analyses used paired observations for each available rating.

**Simulations and domain models**

The detection simulations generated ordinal responses from a fitted two-factor model for 3,955 clients with item 9 pooled, with 500 replicates per condition. Injections affected items 2 and 5: a 25% proportional loading increase or uniform threshold shifts of .15/.30 latent SD. Scenarios used no label perturbation, 2% symmetric perturbation, or directional diagnosed-to-reference/reference-to-diagnosed proportions 11.8%/18.8% and 21.5%/35.3%, informed by disagreement scenarios [4]. Detection was an adjacent nested-model test at the loading or intercept step as appropriate. Threshold-step results for uniform shifts are identification diagnostics because the threshold model can absorb those shifts. Reported outputs include Monte Carlo SEs, intervals and approximate-fit flags (Table S21).

The domain-mean model constrained thresholds, loadings and intercepts with item 9 pooled; diagnosed factor means were zero and assessed comparison means were estimated in diagnosed SD units. Table S23 reports estimates with the diagnosed group as the reference. Extended Results gives the contrast with the assessed comparison group as the reference.

### Supplementary results

#### Participants and ascertainment

First-administration analyses used one record per client; eligible administrations exclude same-day repeats. The portal requires every item before submission, so all first administrations were complete.

**Table S1.** Participants, ascertainment, and check-in timing

**Panel A. Sample accounting by cohort**

| Classification | Original, clients | Original, administrations | Replication, clients | Replication, administrations |
|---|---|---|---|---|
| Diagnosed | 3,471 | 7,312 | 7,406 | 16,134 |
| Reported prior outside assessment | 41 | 88 | 124 | 239 |
| Assessment not completed | 507 | 991 | 1,263 | 1,673 |
| Assessed comparison | 484 | 1,021 | 1,194 | 2,410 |
| No in-service assessment record | 572 | 1,347 | 592 | 1,303 |
| Completed assessment outcome missing | 2 | 7 | 0 | 0 |
| Total | 5,077 | 10,766 | 10,579 | 21,759 |

**Panel B. Primary cohort counts**

| Cohort | Diagnosed | Assessed comparison | Primary total |
|---|---|---|---|
| Original | 3,471 | 484 | 3,955 |
| Replication | 7,406 | 1,194 | 8,600 |

The extract could not isolate self-identification alone, and a prior outside assessment may not indicate a prior autism diagnosis.

**Panel C. Original demographic summary**

| Characteristic | Diagnosed, n (%) | Assessed comparison, n (%) |
|---|---|---|
| Age, mean (SD), years | 35.70 (9.49) | 38.08 (9.68) |
| Age missing, n | 1 | 0 |
| Women | 2,167 (62.4) | 350 (72.3) |
| Men | 755 (21.8) | 104 (21.5) |
| Gender-diverse | 424 (12.2) | 21 (4.3) |
| Gender unclassified or missing | 125 (3.6) | 9 (1.9) |
| White | 2,443 (70.4) | 328 (67.8) |
| Black | 331 (9.5) | 64 (13.2) |
| Multiracial | 213 (6.1) | 35 (7.2) |

| Characteristic | Diagnosed, n (%) | Assessed comparison, n (%) |
|---|---|---|
| Asian | 86 (2.5) | 13 (2.7) |
| Other race categories, prefer not, or missing | 398 (11.5) | 44 (9.1) |
| Hispanic or Latine | 554 (16.0) | 86 (17.8) |
| Ethnicity missing | 144 (4.1) | 7 (1.4) |
| Treatment-plan codes available | 2,951 (85.0) | 405 (83.7) |
| Attention-deficit/hyperactivity disorder code among those with codes | 551 (18.7) | 89 (22.0) |
| Baseline at therapy intake | 2,219 (63.9) | 290 (59.9) |
| Depressive-disorder code (F32/F33), among those with codes | 954 (32.3) | 134 (33.1) |

Gender and race were missing for 558 of 572 no-assessment-record clients (97.6%).

##### Panel D. Replication cohort age and treatment-plan codes

| Group | n | Age, mean (SD) | Missing age | Codes available | Depressive code, n (%) | ADHD code, n (%) |
|---|---|---|---|---|---|---|
| Diagnosed | 7,406 | 36.20 (9.83) | 2 | 5,755 | 1,482 (25.8) | 831 (14.4) |
| Assessed comparison | 1,194 | 38.90 (10.04) | 0 | 863 | 229 (26.5) | 150 (17.4) |

Percentages use clients with available treatment-plan records.

##### Panel E. Replication cohort demographic accounting

| Variable | Recorded category | Diagnosed, n (%) | Assessed comparison, n (%) |
|---|---|---|---|
| Gender | Gender-diverse | 826 (11.2) | 86 (7.2) |
| Gender | Men | 1,557 (21.0) | 253 (21.2) |
| Gender | Missing | 32 (0.4) | 2 (0.2) |
| Gender | Unclassified | 45 (0.6) | 5 (0.4) |
| Gender | Women | 4,946 (66.8) | 848 (71.0) |
| Race | Asian | 210 (2.8) | 39 (3.3) |
| Race | Black | 840 (11.3) | 210 (17.6) |
| Race | Missing | 32 (0.4) | 2 (0.2) |
| Race | Multiracial | 417 (5.6) | 69 (5.8) |
| Race | Native American | 44 (0.6) | 10 (0.8) |
| Race | Native Hawaiian / Pacific Islander | 8 (0.1) | 3 (0.3) |
| Race | Prefer not to answer | 313 (4.2) | 49 (4.1) |
| Race | Self-described only | 266 (3.6) | 47 (3.9) |
| Race | White | 5,276 (71.2) | 765 (64.1) |
| Ethnicity | Not Hispanic or Latine | 6,052 (81.7) | 951 (79.6) |
| Ethnicity | Hispanic or Latine | 1,151 (15.5) | 216 (18.1) |
| Ethnicity | Missing | 203 (2.7) | 27 (2.3) |

##### Panel F. Intake linkage and follow-up in the primary comparison

| Cohort | Assessment group | First check-in at intake, n/N (%) | Second check-in by window close, n/N (%) | Days between check-ins, median (IQR) |
|---|---|---|---|---|
| Original | Primary comparison | 2,509/3,955 (63.4) | 2,128/3,955 (53.8) | 33 (29–42) |
| Original | Diagnosed | 2,219/3,471 (63.9) | 1,868/3,471 (53.8) | 33 (28–41) |

| Cohort | Assessment group | First check-in at intake, n/N (%) | Second check-in by window close, n/N (%) | Days between check-ins, median (IQR) |
|---|---|---|---|---|
| Original | Assessed comparison | 290/484 (59.9) | 260/484 (53.7) | 34 (29–42) |
| Replication | Primary comparison | 8,022/8,600 (93.3) | 4,544/8,600 (52.8) | 31 (29–37) |
| Replication | Diagnosed | 6,915/7,406 (93.4) | 3,962/7,406 (53.5) | 31 (29–38) |
| Replication | Assessed comparison | 1,107/1,194 (92.7) | 582/1,194 (48.7) | 31 (28–37) |

Observation windows closed March 31 and October 31, 2026.

The 5,077 clients in the original analytic cohort represented approximately 66.5% of the 7,633 unique clients with at least one completed, nondeleted psychotherapy appointment during the same window. The aggregate appointment count was not screened for test accounts or reconciled against all study eligibility criteria.

#### Score distributions and percentiles

**Table S2.** Score distributions and percentiles

**Panel A. Original score bands**

| Score band | Diagnosed, n (%) | Assessed comparison, n (%) |
|---|---|---|
| Minimal, 0–4 | 762 (22.0) | 135 (27.9) |
| Mild, 5–9 | 1,206 (34.7) | 174 (36.0) |
| Moderate, 10–14 | 830 (23.9) | 98 (20.2) |
| Moderately severe, 15–19 | 436 (12.6) | 55 (11.4) |
| Severe, 20–27 | 237 (6.8) | 22 (4.5) |

**Panel B. Empirical percentiles**

| Group | 5th / 10th | 25th / 50th | 75th / 90th / 95th |
|---|---|---|---|
| The diagnosed group | 1 / 3 | 5 / 8 | 13 / 18 / 21 |
| The assessed comparison group | 1 / 2 | 4 / 7 | 12 / 17 / 19 |

**Panel C. Original item means**

| Item | Diagnosed | Assessed comparison | Difference (95% CI) | Benjamini–Hochberg (BH) P |
|---|---|---|---|---|
| 1 anhedonia | 1.05 | 0.95 | 0.10 (0.02 to 0.19) | .03 |
| 2 depressed mood | 1.00 | 0.93 | 0.08 (−0.01 to 0.16) | .09 |
| 3 sleep | 1.45 | 1.31 | 0.14 (0.04 to 0.24) | .02 |
| 4 fatigue | 1.67 | 1.50 | 0.17 (0.08 to 0.26) | .002 |
| 5 appetite | 1.14 | 1.01 | 0.12 (0.02 to 0.22) | .03 |
| 6 worthlessness | 1.02 | 0.90 | 0.11 (0.02 to 0.21) | .03 |
| 7 concentration | 1.28 | 1.23 | 0.04 (−0.06 to 0.14) | .42 |
| 8 psychomotor | 0.58 | 0.44 | 0.14 (0.07 to 0.22) | .002 |

| Item | Diagnosed | Assessed comparison | Difference (95% CI) | Benjamini–Hochberg (BH) P |
|---|---|---|---|---|
| 9 death or self-harm thoughts | 0.20 | 0.16 | 0.04 (0.00 to 0.09) | .09 |

Percentiles use inverse empirical type 1 quantiles; BH adjustment covers the nine item-mean tests.

**Panel D. Observed PHQ-9 score distribution by cohort**

| Cohort and group | n | Mean (SD) | Score ≥10, n (%) | Floor 0, n (%) | Ceiling 27, n (%) |
|---|---|---|---|---|---|
| Original: diagnosed | 3,471 | 9.39 (5.77) | 1,503 (43.30) | 90 (2.59) | 5 (0.14) |
| Original: assessed comparison | 484 | 8.44 (5.67) | 175 (36.16) | 18 (3.72) | 1 (0.21) |
| Replication: diagnosed | 7,406 | 9.70 (5.69) | 3,361 (45.38) | 129 (1.74) | 19 (0.26) |
| Replication: assessed comparison | 1,194 | 8.90 (5.37) | 476 (39.87) | 33 (2.76) | 5 (0.42) |

Mean difference, diagnosed minus assessed comparison: original 0.95 points (Welch 95% CI 0.41–1.49), Hedges g 0.17 (0.07–0.26); replication 0.80 (0.47–1.13), g 0.14 (0.08–0.20).

#### Factor model fit

The diagnosed conventional bifactor model did not yield a reproducible solution across starting values (all 100 random starts converged to an inadmissible alternative). All 101 assessed comparison starts reproduced one proper solution. The S-1 results depend on their reference domain.

**Table S3.** Model fit

**Panel A. Diagnosed models**

| Model | Scaled $\chi^2$ and df | Scaled CFI / TLI / RMSEA | Robust CFI / TLI / RMSEA |
|---|---|---|---|
| One factor | 1237.51 (27) | 0.955 / 0.940 / 0.114 | 0.879 / 0.839 / 0.146 |
| Two factor | 844.21 (26) | 0.970 / 0.958 / 0.095 | 0.917 / 0.886 / 0.123 |
| Bifactor | 250.46 (18) | 0.991 / 0.983 / 0.061 | 0.798 / 0.597 / 0.231 |
| S-1 cognitive-affective reference | 676.80 (23) | 0.976 / 0.962 / 0.0905 | 0.931 / 0.892 / 0.120 |
| S-1 somatic reference | 458.17 (22) | 0.984 / 0.974 / 0.076 | 0.951 / 0.920 / 0.103 |

**Panel B. Assessed comparison models**

| Model | Scaled $\chi^2$ and df | Scaled CFI / TLI / RMSEA | Robust CFI / TLI / RMSEA |
|---|---|---|---|
| One factor | 212.33 (27) | 0.955 / 0.940 / 0.119 | 0.863 / 0.817 / 0.166 |
| Two factor | 146.03 (26) | 0.9706 / 0.959 / 0.098 | 0.9085 / 0.873 / 0.138 |
| Bifactor | 23.82 (18) | 0.999 / 0.997 / 0.026 | 0.991 / 0.983 / 0.051 |
| S-1 cognitive-affective reference | 130.23 (23) | 0.974 / 0.959 / 0.098 | 0.916 / 0.869 / 0.141 |
| S-1 somatic reference | 52.68 (22) | 0.993 / 0.988 / 0.054 | 0.978 / 0.965 / 0.073 |

SRMR (one-factor / two-factor / S-1 cognitive-affective reference / S-1 somatic reference): .062/.053/.046/.039 diagnosed and .071/.059/.055/.032 assessed comparison; conventional bifactor .028 and .022.

##### Panel C. Replication cohort one- and two-factor models

| Group | Model | Scaled $\chi^2$ (df) | Scaled CFI / RMSEA / SRMR | Robust CFI / RMSEA |
|---|---|---|---|---|
| Diagnosed | one factor | 2884.636 (27) | 0.944 / 0.120 / 0.068 | 0.856 / 0.154 |
| Diagnosed | two factor | 1957.408 (26) | 0.962 / 0.100 / 0.057 | 0.905 / 0.127 |
| Assessed comparison | one factor | 548.219 (27) | 0.930 / 0.127 / 0.077 | 0.823 / 0.166 |
| Assessed comparison | two factor | 423.479 (26) | 0.947 / 0.113 / 0.070 | 0.867 / 0.146 |

##### Panel D. Standardized primary two-factor loadings

| Item | Domain | Original diagnosed | Original assessed | Replication cohort diagnosed | Replication cohort assessed |
|---|---|---|---|---|---|
| 1 | Cognitive-affective | 0.843 | 0.856 | 0.827 | 0.843 |
| 2 | Cognitive-affective | 0.879 | 0.866 | 0.879 | 0.862 |
| 3 | Somatic | 0.690 | 0.630 | 0.667 | 0.699 |
| 4 | Somatic | 0.795 | 0.836 | 0.780 | 0.756 |
| 5 | Somatic | 0.716 | 0.765 | 0.693 | 0.703 |
| 6 | Cognitive-affective | 0.768 | 0.809 | 0.752 | 0.714 |
| 7 | Cognitive-affective | 0.654 | 0.676 | 0.636 | 0.623 |
| 8 | Somatic | 0.621 | 0.680 | 0.616 | 0.608 |
| 9 | Cognitive-affective | 0.712 | 0.788 | 0.713 | 0.576 |

Assessed denotes assessed comparison.

##### Panel E. Latent-response omega total and standardized factor correlations

| Statistic | Original diagnosed | Original assessed comparison | Replication diagnosed | Replication assessed comparison |
|---|---|---|---|---|
| One-factor latent-response ωT | 0.905 | 0.916 | 0.897 | 0.887 |
| Two-factor latent-response ωT | 0.912 | 0.923 | 0.905 | 0.895 |
| Standardized factor correlation (Wald 95% CI) | 0.852 (0.834–0.870) | 0.838 (0.786–0.890) | 0.825 (0.811–0.840) | 0.834 (0.800–0.868) |

ωT values are latent-response estimates, distinct from the Schmid–Leiman ωT in Table S4.

#### Reliability and dimensionality

**Table S4.** Reliability and dimensionality

##### Panel A. Reliability summaries

| Index | Original diagnosed | Original assessed comparison |
|---|---|---|
| Raw alpha | .857 | .862 |
| SL omega total | .926 | .933 |
| SL omega hierarchical | .775 | .772 |

| Index | Original diagnosed | Original assessed comparison |
|---|---|---|
| SL ECV | .690 | .689 |
| GRM reliability | .861 | .868 |

Replication (diagnosed / assessed comparison): alpha .847/.838; GRM sum-score reliability .849/.843; SL ωH .742/.696; general share .809/.763.

##### Panel B. Common factor parallel analysis

| Factor | Diagnosed observed | Diagnosed simulated mean | Assessed comparison observed | Assessed comparison simulated mean |
|---|---|---|---|---|
| 1 | 4.532 | 0.345 | 4.780 | 0.511 |
| 2 | 0.425 | 0.059 | 0.530 | 0.158 |
| 3 | 0.183 | 0.040 | 0.123 | 0.108 |
| 4 | 0.063 | 0.024 | 0.018 | 0.062 |
| 5 | −0.068 | 0.008 | −0.044 | 0.019 |
| 6 | −0.109 | −0.006 | −0.067 | −0.020 |
| 7 | −0.132 | −0.022 | −0.146 | −0.061 |
| 8 | −0.150 | −0.040 | −0.192 | −0.106 |
| 9 | −0.212 | −0.062 | −0.223 | −0.161 |

Simulated columns are mean eigenvalues across 500 simulations; 4 (diagnosed) and 3 (assessed comparison) observed eigenvalues exceeded them.

##### Panel C. Conventional confirmatory bifactor indices

| Index | Diagnosed | Assessed comparison |
|---|---|---|
| Omega total | 0.920 | 0.933 |
| Omega hierarchical | 0.850 | 0.844 |
| ECV | 0.794 | 0.761 |
| PUC | 0.556 | 0.556 |
| Omega hierarchical, cognitive-affective subscale | 0.118 | 0.145 |
| Omega hierarchical, somatic subscale | 0.120 | 0.161 |

SL denotes Schmid–Leiman.

##### Panel D. Replication cohort Schmid–Leiman decomposition

| Group | n | Omega total | Omega hierarchical | General share | ECV |
|---|---|---|---|---|---|
| Diagnosed | 7,406 | 0.917 | 0.742 | 0.809 | 0.656 |
| Assessed comparison | 1,194 | 0.912 | 0.696 | 0.763 | 0.598 |

##### Panel E. Replication cohort parallel analysis

| Group | Suggested common factors | Iterations |
|---|---|---|
| Diagnosed | 4 | 500 |
| Assessed comparison | 3 | 500 |

The four-factor diagnosed sensitivity converged without a Heywood case.

#### Item response parameters and Rasch sensitivity

**Table S5.** Graded response and Rasch results

##### Panel A. Diagnosed GRM

| Item | Discrimination | Thresholds 1 / 2 / 3 |
|---|---|---|
| 1 Anhedonia | 2.698 | −0.660 / 0.770 / 1.611 |
| 2 Depressed mood | 2.922 | −0.574 / 0.850 / 1.602 |
| 3 Sleep | 1.385 | −1.277 / 0.257 / 1.171 |
| 4 Fatigue | 1.860 | −1.662 / −0.030 / 0.817 |
| 5 Appetite | 1.515 | −0.581 / 0.634 / 1.518 |
| 6 Worthlessness | 1.956 | −0.485 / 0.822 / 1.600 |
| 7 Concentration | 1.435 | −0.943 / 0.519 / 1.446 |
| 8 Psychomotor change | 1.212 | 0.445 / 1.869 / 2.886 |
| 9 Death or self-harm thoughts | 1.684 | 1.453 / 2.627 / 3.390 |

##### Panel B. Assessed comparison GRM

| Item | Discrimination | Thresholds 1 / 2 / 3 |
|---|---|---|
| 1 Anhedonia | 2.927 | −0.489 / 0.965 / 1.568 |
| 2 Depressed mood | 2.524 | −0.477 / 0.976 / 1.767 |
| 3 Sleep | 1.210 | −1.186 / 0.551 / 1.515 |
| 4 Fatigue | 2.041 | −1.432 / 0.204 / 1.052 |
| 5 Appetite | 1.796 | −0.430 / 0.823 / 1.615 |
| 6 Worthlessness | 2.032 | −0.311 / 1.051 / 1.706 |
| 7 Concentration | 1.566 | −0.777 / 0.543 / 1.394 |
| 8 Psychomotor change | 1.446 | 0.833 / 1.882 / 2.862 |
| 9 Death or self-harm thoughts | 2.081 | 1.517 / 2.562 / 3.056 |

##### Panel C. Rasch fit sensitivity

| Item | Diagnosed infit | Diagnosed outfit | Assessed comparison infit | Assessed comparison outfit |
|---|---|---|---|---|
| 1 Anhedonia | 0.625 | 0.607 | 0.624 | 0.583 |
| 2 Depressed mood | 0.620 | 0.603 | 0.692 | 0.648 |
| 3 Sleep | 0.986 | 0.950 | 1.121 | 1.087 |
| 4 Fatigue | 0.716 | 0.698 | 0.641 | 0.620 |
| 5 Appetite | 1.045 | 0.994 | 0.989 | 0.900 |
| 6 Worthlessness | 0.900 | 0.856 | 0.912 | 0.867 |
| 7 Concentration | 1.002 | 0.980 | 1.046 | 0.997 |
| 8 Psychomotor change | 1.274 | 1.131 | 1.393 | 1.083 |
| 9 Death or self-harm thoughts | 1.267 | 0.891 | 1.256 | 0.741 |

Rasch marginal person reliability was 0.747 (diagnosed) and 0.700 (assessed comparison). Conditional reliability was at least .80 for model-expected totals of 3.1–22.3 and 2.7–22.9, respectively.

**Panel D. Replication cohort diagnosed graded response parameters**

| Item | Discrimination | Threshold 1 | Threshold 2 | Threshold 3 |
|---|---|---|---|---|
| 1 | 2.540 | −0.672 | 0.729 | 1.535 |
| 2 | 2.862 | −0.603 | 0.834 | 1.604 |
| 3 | 1.253 | −1.484 | 0.164 | 1.208 |
| 4 | 1.706 | −1.867 | −0.098 | 0.750 |
| 5 | 1.403 | −0.614 | 0.684 | 1.626 |
| 6 | 1.893 | −0.538 | 0.759 | 1.583 |
| 7 | 1.335 | −1.172 | 0.404 | 1.362 |
| 8 | 1.161 | 0.400 | 1.777 | 2.907 |
| 9 | 1.646 | 1.431 | 2.641 | 3.393 |

**Panel E. Replication cohort assessed comparison graded response parameters**

| Item | Discrimination | Threshold 1 | Threshold 2 | Threshold 3 |
|---|---|---|---|---|
| 1 | 2.678 | −0.654 | 0.828 | 1.560 |
| 2 | 2.651 | −0.618 | 0.912 | 1.725 |
| 3 | 1.357 | −1.235 | 0.365 | 1.374 |
| 4 | 1.710 | −1.821 | 0.141 | 0.966 |
| 5 | 1.521 | −0.426 | 0.876 | 1.798 |
| 6 | 1.713 | −0.524 | 0.952 | 1.832 |
| 7 | 1.253 | −1.094 | 0.610 | 1.633 |
| 8 | 1.121 | 0.804 | 2.271 | 3.516 |
| 9 | 1.206 | 1.823 | 3.378 | 4.138 |

**Panel F. Replication cohort information summary**

| Group | Items 1+2 share | Item 9 share | Peak expected total | Peak theta |
|---|---|---|---|---|
| Assessed comparison | 0.465 | 0.024 | 14.792 | 1.100 |
| Diagnosed | 0.457 | 0.041 | 15.313 | 1.000 |

Information shares are weighted at empirical EAP trait estimates.

#### Item fit

**Table S6.** Item fit

**Panel A. Diagnosed item fit**

| Item | S-X² (df) | Item RMSEA | P / adjusted P |
|---|---|---|---|
| 1 Anhedonia | 64.64 (45) | 0.011 | P=.03 / P=.04 |
| 2 Depressed mood | 116.25 (44) | 0.022 | P<.001 / P<.001 |
| 3 Sleep | 72.19 (54) | 0.010 | P=.0497 / P=.06 |
| 4 Fatigue | 43.75 (47) | 0.000 | P=.61 / P=.61 |
| 5 Appetite | 78.99 (55) | 0.011 | P=.02 / P=.03 |
| 6 Worthlessness | 92.98 (53) | 0.015 | P<.001 / P=.003 |
| 7 Concentration | 61.49 (55) | 0.006 | P=.26 / P=.29 |
| 8 Psychomotor change | 87.55 (59) | 0.012 | P=.009 / P=.02 |

| Item | S-X² (df) | Item RMSEA | P / adjusted P |
|---|---|---|---|
| 9 Death or self-harm thoughts | 74.75 (48) | 0.013 | P=.008 / P=.02 |

**Panel B. Assessed comparison item fit**

| Item | S-X² (df) | Item RMSEA | P / adjusted P |
|---|---|---|---|
| 1 Anhedonia | 25.35 (28) | 0.000 | P=.61 / P=.78 |
| 2 Depressed mood | 24.51 (30) | 0.000 | P=.75 / P=.84 |
| 3 Sleep | 45.08 (44) | 0.007 | P=.43 / P=.64 |
| 4 Fatigue | 37.39 (30) | 0.023 | P=.17 / P=.50 |
| 5 Appetite | 27.24 (39) | 0.000 | P=.92 / P=.92 |
| 6 Worthlessness | 37.69 (34) | 0.015 | P=.30 / P=.64 |
| 7 Concentration | 54.42 (39) | 0.029 | P=.051 / P=.23 |
| 8 Psychomotor change | 38.93 (37) | 0.010 | P=.38 / P=.64 |
| 9 Death or self-harm thoughts | 29.87 (14) | 0.048 | P=.008 / P=.07 |

Original C2 was 1,016.92 and 186.74 (27 df; RMSEA .103 and .111) and replication C2 was 2,545.32 and 466.37 (27 df; RMSEA .112 and .117) in diagnosed and assessed comparison clients, respectively.

**Panel C. Replication cohort diagnosed S-X² item fit**

| Item | S-X² (df) | Item RMSEA | P | BH P |
|---|---|---|---|---|
| 1 | 163.553 (51) | 0.017 | <.001 | <.001 |
| 2 | 350.771 (48) | 0.029 | <.001 | <.001 |
| 3 | 89.787 (57) | 0.009 | .004 | .004 |
| 4 | 128.972 (51) | 0.014 | <.001 | <.001 |
| 5 | 100.010 (59) | 0.010 | <.001 | <.001 |
| 6 | 172.300 (55) | 0.017 | <.001 | <.001 |
| 7 | 71.830 (58) | 0.006 | .105 | .105 |
| 8 | 112.746 (61) | 0.011 | <.001 | <.001 |
| 9 | 155.405 (53) | 0.016 | <.001 | <.001 |

**Panel D. Replication cohort assessed comparison S-X² item fit**

| Item | S-X² (df) | Item RMSEA | P | BH P |
|---|---|---|---|---|
| 1 | 58.219 (36) | 0.023 | .011 | .049 |
| 2 | 69.282 (36) | 0.028 | <.001 | .006 |
| 3 | 47.012 (46) | 0.004 | .431 | .554 |
| 4 | 51.103 (40) | 0.015 | .112 | .220 |
| 5 | 41.915 (46) | 0.000 | .644 | .724 |
| 6 | 60.524 (44) | 0.018 | .050 | .149 |
| 7 | 34.776 (47) | 0.000 | .907 | .907 |
| 8 | 65.137 (53) | 0.014 | .122 | .220 |
| 9 | 45.154 (37) | 0.014 | .168 | .252 |

#### Local dependence

No ordinal CFA residual reached absolute .20, whereas the unidimensional GRM showed residual dependence.

**Table S7.** Local dependence

**Panel A. Maximum adjusted Q3 values and bootstrap critical values**

| Group | Observed maximum | 95th percentile | 99th percentile | Pairs above the 95th percentile | Successful replicates |
|---|---|---|---|---|---|
| Diagnosed | 0.275 | 0.098 | 0.108 | 7 | 500 of 500 |
| Assessed comparison | 0.324 | 0.177 | 0.207 | 4 | 500 of 500 |

**Panel B. Flagged item pairs**

| Diagnosed: pair | Adjusted Q3 | Assessed comparison: pair | Adjusted Q3 |
|---|---|---|---|
| Sleep with fatigue | 0.275 | Worthlessness with depressed mood | 0.324 |
| Concentration with psychomotor change | 0.243 | Sleep with fatigue | 0.298 |
| Sleep with appetite | 0.182 | Item 9 with depressed mood | 0.225 |
| Fatigue with appetite | 0.143 | Item 9 with worthlessness | 0.179 |
| Item 9 with worthlessness | 0.137 | | |
| Worthlessness with depressed mood | 0.113 | | |
| Sleep with psychomotor change | 0.102 | | |

**Panel C. Residual summary for all item pairs**

| Pair | Diagnosed adjusted Q3 | Assessed comparison adjusted Q3 | Diagnosed two-factor CFA | Assessed comparison two-factor CFA |
|---|---|---|---|---|
| Items 1 and 2 | 0.017 | −0.002 | 0.020 | −0.007 |
| Items 1 and 3 | −0.074 | −0.028 | −0.052 | −0.087 |
| Items 1 and 4 | −0.066 | −0.119 | −0.014 | 0.031 |
| Items 1 and 5 | −0.130 | −0.145 | −0.038 | −0.076 |

| Pair | Diagnosed adjusted Q3 | Assessed comparison adjusted Q3 | Diagnosed two-factor CFA | Assessed comparison two-factor CFA |
|---|---|---|---|---|
| Items 1 and 6 | −0.144 | −0.173 | −0.002 | 0.013 |
| Items 1 and 7 | −0.016 | 0.058 | 0.030 | 0.033 |
| Items 1 and 8 | −0.076 | 0.071 | −0.008 | 0.029 |
| Items 1 and 9 | −0.026 | −0.104 | −0.006 | 0.077 |
| Items 2 and 3 | −0.183 | −0.154 | 0.045 | 0.070 |
| Items 2 and 4 | −0.163 | −0.197 | −0.095 | −0.131 |
| Items 2 and 5 | −0.188 | −0.166 | 0.038 | 0.091 |
| Items 2 and 6 | 0.113 | 0.324 | −0.061 | −0.092 |
| Items 2 and 7 | −0.165 | −0.211 | −0.023 | −0.060 |
| Items 2 and 8 | −0.125 | −0.138 | −0.036 | −0.025 |
| Items 2 and 9 | 0.096 | 0.225 | −0.039 | −0.080 |
| Items 3 and 4 | 0.275 | 0.298 | −0.070 | −0.067 |
| Items 3 and 5 | 0.182 | 0.120 | 0.075 | 0.053 |
| Items 3 and 6 | −0.107 | −0.118 | −0.063 | −0.085 |
| Items 3 and 7 | 0.057 | 0.030 | −0.042 | −0.055 |
| Items 3 and 8 | 0.102 | 0.150 | 0.002 | 0.002 |
| Items 3 and 9 | −0.030 | −0.031 | 0.048 | −0.016 |
| Items 4 and 5 | 0.143 | 0.113 | −0.081 | −0.129 |
| Items 4 and 6 | −0.126 | −0.135 | 0.027 | 0.012 |
| Items 4 and 7 | 0.064 | 0.076 | 0.049 | 0.063 |
| Items 4 and 8 | −0.017 | 0.001 | 0.060 | 0.082 |
| Items 4 and 9 | −0.052 | 0.033 | 0.176 | 0.127 |
| Items 5 and 6 | −0.043 | −0.078 | −0.083 | −0.067 |
| Items 5 and 7 | 0.087 | 0.069 | −0.090 | 0.006 |
| Items 5 and 8 | 0.094 | 0.113 | 0.014 | −0.054 |
| Items 5 and 9 | 0.037 | −0.058 | 0.049 | −0.041 |
| Items 6 and 7 | −0.058 | −0.051 | 0.063 | 0.066 |
| Items 6 and 8 | 0.067 | −0.020 | 0.021 | −0.005 |
| Items 6 and 9 | 0.137 | 0.179 | −0.038 | 0.017 |
| Items 7 and 8 | 0.243 | 0.119 | −0.010 | −0.027 |
| Items 7 and 9 | −0.004 | −0.013 | −0.129 | −0.081 |
| Items 8 and 9 | 0.079 | −0.039 | −0.022 | −0.018 |

Original mean raw Q3 values were −.097 and −.102; the bootstrap used 500 refitted datasets per group, all successful.

#### Panel D. Replication cohort Q3 values by item pair

| Item pair | Diagnosed raw | Diagnosed adjusted | Assessed raw | Assessed adjusted |
|---|---|---|---|---|
| 3–4 | 0.202 | 0.297 | 0.206 | 0.297 |
| 7–8 | 0.152 | 0.247 | 0.197 | 0.287 |
| 6–9 | 0.088 | 0.183 | 0.091 | 0.182 |
| 3–5 | 0.074 | 0.169 | 0.028 | 0.119 |
| 4–5 | 0.049 | 0.144 | −0.001 | 0.090 |
| 5–8 | 0.041 | 0.136 | 0.051 | 0.142 |
| 4–7 | 0.030 | 0.125 | 0.014 | 0.105 |
| 2–9 | 0.020 | 0.115 | 0.067 | 0.158 |

| Item pair | Diagnosed raw | Diagnosed adjusted | Assessed raw | Assessed adjusted |
|---|---|---|---|---|
| 3–7 | 0.011 | 0.106 | 0.061 | 0.151 |
| 2–6 | −0.007 | 0.088 | 0.035 | 0.126 |
| 5–7 | −0.007 | 0.088 | −0.013 | 0.078 |
| 3–8 | −0.017 | 0.078 | −0.011 | 0.080 |
| 8–9 | −0.056 | 0.039 | 0.005 | 0.096 |
| 1–2 | −0.057 | 0.038 | −0.024 | 0.067 |
| 4–8 | −0.064 | 0.031 | −0.082 | 0.009 |
| 6–8 | −0.065 | 0.030 | −0.029 | 0.062 |
| 7–9 | −0.109 | −0.014 | −0.084 | 0.007 |
| 5–9 | −0.112 | −0.017 | −0.070 | 0.021 |
| 6–7 | −0.118 | −0.023 | −0.084 | 0.006 |
| 3–9 | −0.125 | −0.030 | −0.090 | 0.001 |
| 1–9 | −0.133 | −0.038 | −0.148 | −0.057 |
| 5–6 | −0.139 | −0.044 | −0.109 | −0.018 |
| 1–4 | −0.145 | −0.050 | −0.145 | −0.054 |
| 4–9 | −0.149 | −0.054 | −0.147 | −0.056 |
| 1–7 | −0.152 | −0.057 | −0.214 | −0.123 |
| 1–8 | −0.172 | −0.077 | −0.201 | −0.110 |
| 1–3 | −0.176 | −0.081 | −0.230 | −0.139 |
| 3–6 | −0.192 | −0.097 | −0.222 | −0.132 |
| 1–5 | −0.210 | −0.115 | −0.236 | −0.145 |
| 2–8 | −0.227 | −0.132 | −0.227 | −0.136 |
| 4–6 | −0.238 | −0.143 | −0.220 | −0.129 |
| 1–6 | −0.261 | −0.166 | −0.254 | −0.163 |
| 2–3 | −0.273 | −0.178 | −0.261 | −0.170 |
| 2–5 | −0.275 | −0.180 | −0.270 | −0.179 |
| 2–7 | −0.293 | −0.198 | −0.303 | −0.212 |
| 2–4 | −0.314 | −0.219 | −0.349 | −0.259 |

Replication anchors 3 and 7 belonged to flagged pairs; anchor 6 did not.

Concentration–psychomotor change exceeded the descriptive mean-centered Q3 flag of 0.20 in three of four groups.

#### Analysis aims and evidence

**Table S8.** Analysis aims and supporting results

| Aim | Analytic evidence | Main evidence |
|---|---|---|
| Secondary: Internal structure and reliability | Model fit, factor correlations, alpha, omega, and explained common variance | Table 1; Tables S3–S4 |
| Secondary: Precision | Alpha-based SEM, information and model-based reliability | Information results; alpha-based SEM in Table 1 |
| Secondary: Association with anxiety | PHQ-9–GAD-7 correlations, joint ordinal CFAs, and HTMT | Table S9 |

| Aim | Analytic evidence | Main evidence |
|---|---|---|
| Primary: Expected scores by assessment outcome | Range of signed differences and largest upper bound on the average absolute difference across anchor sets, interpreted relative to the alpha-based SEM | Figures 2–3; Tables S14 and S16 |
| Secondary: Item 9 endorsement at the same PHQ-8 score | Assess whether the PHQ-8–adjusted group contrast has the same direction in both cohorts | Table 3; Table S18 |
| Exploratory: Score comparability within diagnosed subgroups | Describe original-scale effects and one-sided 95% upper confidence limits for average absolute differences | Tables S11–S15 and S19 |

#### Anxiety and wellbeing associations

**Table S9.** Anxiety and wellbeing associations

##### Panel A. HTMT uncertainty and diagnostics

| Quantity | Diagnosed | Assessed comparison |
|---|---|---|
| Point estimate | 0.811 | 0.841 |
| Two-sided 95% CI | 0.792 to 0.831 | 0.797 to 0.881 |
| One-sided 95% upper | 0.827 | 0.875 |
| Two-sided 90% CI | 0.795 to 0.827 | 0.804 to 0.875 |
| Attempted replications | 2,000 | 2,000 |
| Successful replications | 2,000 | 1,995 |
| Failed replications | 0 | 5 |
| Nonpositive definite matrices | 0 | 6 |

##### Panel B. Wellbeing correlations

| Correlate and method | Diagnosed (95% CI) | Assessed comparison (95% CI) |
|---|---|---|
| Stress and emotional functioning, Pearson | −0.533 (−0.556 to −0.509) | −0.595 (−0.650 to −0.534) |
| Stress and emotional functioning, Spearman | −0.543 (−0.568 to −0.518) | −0.618 (−0.675 to −0.554) |
| Overall wellbeing, Pearson | −0.498 (−0.523 to −0.473) | −0.414 (−0.485 to −0.337) |
| Overall wellbeing, Spearman | −0.538 (−0.563 to −0.512) | −0.462 (−0.533 to −0.385) |

##### Panel C. Joint depression and anxiety model

| Group and model | CFI | TLI | RMSEA | SRMR |
|---|---|---|---|---|
| Diagnosed two factors | .966 | .960 | .094 | .058 |
| Diagnosed one factor | .935 | .924 | .129 | .079 |
| Assessed comparison two factors | .966 | .960 | .095 | .067 |
| Assessed comparison one factor | .943 | .933 | .123 | .082 |

Original Pearson r .707 (.690–.723) and .742 (.699–.780); Spearman ρ .706 (.687–.724) and .719 (.667–.764); depression–anxiety latent r .790 (.773–.807) and .815 (.778–.853).

##### Panel D. Replication cohort observed-score correlations

| Group | Correlate | Method | n | r or rho | 95% CI |
|---|---|---|---|---|---|
| Assessed comparison | GAD-7 total | Pearson | 1,194 | 0.697 | 0.667 to 0.725 |
| Assessed comparison | GAD-7 total | Spearman | 1,194 | 0.683 | 0.648 to 0.715 |
| Assessed comparison | Stress/emotional functioning | Pearson | 1,194 | −0.514 | −0.554 to −0.471 |
| Assessed comparison | Stress/emotional functioning | Spearman | 1,194 | −0.521 | −0.564 to −0.475 |
| Assessed comparison | Overall wellbeing | Pearson | 1,194 | −0.374 | −0.422 to −0.324 |
| Assessed comparison | Overall wellbeing | Spearman | 1,194 | −0.418 | −0.465 to −0.367 |
| Diagnosed | GAD-7 total | Pearson | 7,406 | 0.687 | 0.675 to 0.699 |
| Diagnosed | GAD-7 total | Spearman | 7,406 | 0.680 | 0.666 to 0.693 |
| Diagnosed | Stress/emotional functioning | Pearson | 7,406 | −0.501 | −0.518 to −0.484 |
| Diagnosed | Stress/emotional functioning | Spearman | 7,406 | −0.509 | −0.527 to −0.491 |
| Diagnosed | Overall wellbeing | Pearson | 7,406 | −0.435 | −0.453 to −0.416 |
| Diagnosed | Overall wellbeing | Spearman | 7,406 | −0.475 | −0.494 to −0.457 |

##### Panel E. Replication cohort HTMT

| Group | n | HTMT | 95% percentile CI | Successful / attempted | Failed |
|---|---|---|---|---|---|
| Diagnosed | 7,406 | 0.795 | 0.781 to 0.808 | 2,000/2,000 | 0 |
| Assessed comparison | 1,194 | 0.823 | 0.790 to 0.856 | 2,000/2,000 | 0 |

##### Panel F. Replication cohort joint 16-item CFA

| Group | Model | Scaled CFI | Scaled RMSEA | SRMR | Robust CFI | Robust RMSEA |
|---|---|---|---|---|---|---|
| Diagnosed | One distress factor | 0.914 | 0.139 | 0.088 | 0.735 | 0.166 |
| Diagnosed | Two domain factors | 0.957 | 0.099 | 0.062 | 0.864 | 0.120 |
| Assessed comparison | One distress factor | 0.914 | 0.134 | 0.087 | 0.741 | 0.164 |
| Assessed comparison | Two domain factors | 0.951 | 0.102 | 0.069 | 0.845 | 0.128 |

##### Panel G. Replication cohort depression–anxiety latent correlations

| Group | Correlation | 95% CI |
|---|---|---|
| Diagnosed | 0.760 | 0.748 to 0.772 |
| Assessed comparison | 0.775 | 0.745 to 0.805 |

#### Assessment outcome invariance

**Table S10.** Assessment-outcome invariance

##### Panel A. Original invariance tests

| Step | Unscaled $\chi^2$ / df | Scaled CFI / RMSEA / SRMR | Nested $\chi^2$ and df / P |
|---|---|---|---|
| Configural | 552.35 / 52 | 0.970 / 0.095 / 0.054 | — |

| Step | Unscaled χ² / df | Scaled CFI / RMSEA / SRMR | Nested χ² and df / P |
|---|---|---|---|
| Thresholds | 555.75 / 60 | 0.969 / 0.090 / 0.054 | 9.27 (8) / P=.32 |
| Metric | 563.70 / 67 | 0.972 / 0.080 / 0.054 | 8.20 (7) / P=.315 |
| Scalar | 567.51 / 74 | 0.976 / 0.071 / 0.054 | 3.65 (7) / P=.82 |
| Strict | 590.59 / 83 | 0.979 / 0.063 / 0.055 | 20.60 (9) / P=.015 |

#### Panel B. Original fit changes

| Step | Δ scaled CFI | Δ scaled RMSEA | Δ SRMR |
|---|---|---|---|
| Thresholds | −0.0010 | −0.0051 | 0.0000 |
| Metric | 0.0035 | −0.0097 | 0.0001 |
| Scalar | 0.0040 | −0.0096 | 0.0000 |
| Strict | 0.0027 | −0.0078 | 0.0009 |

#### Panel C. Replication cohort full categories

| Step | Unscaled χ² / df | Scaled CFI / RMSEA / SRMR | Nested χ² and df / P |
|---|---|---|---|
| Configural | 1414.38 / 52 | 0.961 / 0.102 / 0.059 | — |
| Thresholds | 1417.53 / 61 | 0.959 / 0.096 / 0.059 | 8.39 (9) / P=.50 |
| Metric | 1426.53 / 68 | 0.965 / 0.085 / 0.059 | 8.15 (7) / P=.32 |
| Scalar | 1454.11 / 75 | 0.969 / 0.075 / 0.059 | 22.03 (7) / P=.003 |
| Strict | 1486.10 / 84 | 0.971 / 0.068 / 0.060 | 29.90 (9) / P<.001 |

#### Panel D. Replication cohort item 9 pooled

| Step | Scaled CFI | Scaled RMSEA | SRMR | Chi-square difference | df | P |
|---|---|---|---|---|---|---|
| Configural | 0.961 | 0.101 | 0.059 | | | |
| Thresholds | 0.959 | 0.096 | 0.059 | 6.94 | 8 | .54 |
| Metric | 0.964 | 0.086 | 0.059 | 8.90 | 7 | .26 |
| Scalar | 0.969 | 0.076 | 0.059 | 21.87 | 7 | .003 |
| Strict | 0.971 | 0.069 | 0.060 | 28.13 | 9 | <.001 |

Item 9 coded 0/1/2–3 in the original sequence and the replication pooled-category sensitivity; the replication primary sequence retains all four categories.

**Table S10, continued.** Nested-model audit for the standard assessment-outcome models

| Cohort | Step | Effective free parameters | df | Unscaled χ² | Scaled χ² | Unscaled Δχ² |
|---|---|---|---|---|---|---|
| Original | Configural | 72 | 52 | 552.35 | 979.68 | — |
| Original | Thresholds | 64 | 60 | 555.75 | 1019.47 | 3.40 |
| Original | Metric | 57 | 67 | 563.70 | 919.43 | 7.95 |
| Original | Scalar | 50 | 74 | 567.51 | 802.96 | 3.81 |
| Original | Strict | 41 | 83 | 590.59 | 729.66 | 23.08 |
| Replication | Configural | 74 | 52 | 1414.38 | 2361.39 | — |
| Replication | Thresholds | 65 | 61 | 1417.53 | 2471.53 | 3.15 |
| Replication | Metric | 58 | 68 | 1426.53 | 2159.87 | 9.00 |
| Replication | Scalar | 51 | 75 | 1454.11 | 1889.50 | 27.58 |

| Cohort | Step | Effective free parameters | df | Unscaled χ² | Scaled χ² | Unscaled Δχ² |
|---|---|---|---|---|---|---|
| Replication | Strict | 42 | 84 | 1486.10 | 1766.12 | 31.99 |

Effective free parameters = modeled polychoric correlations and thresholds (124 original, 126 replication) minus df.

#### Other invariance sequences

Original intake-only scalar P=.79, assessment-before-baseline P=.49, and ULSMV P=.82. Across 100 balanced subsamples all models converged; thresholds, loadings and scalar tests were significant in three, eight and zero runs. In 1,000 permutations, empirical χ²-change P values were .33, .344, and .840 at these steps.

**Table S11.** Subgroup and other invariance checks

**Panel A. Pairwise subgroup sample sizes**

| Comparison | Original, focal/reference n | Replication, focal/reference n |
|---|---|---|
| Men vs women | 755 / 2,167 | 1,557 / 4,946 |
| Gender-diverse vs women | 424 / 2,167 | 826 / 4,946 |
| Under 35 vs 35 or older | 1,629 / 1,841 | 3,353 / 4,051 |
| Black vs White | 331 / 2,443 | 840 / 5,276 |
| Multiracial vs White | 213 / 2,443 | 417 / 5,276 |
| Hispanic or Latine vs not Hispanic or Latine | 554 / 2,773 | 1,151 / 6,052 |
| Recorded ADHD code vs none among clients with treatment-plan records | 551 / 2,400 | 831 / 4,924 |

**Panel B. Original cohort subgroup invariance tests**

| Comparison | Configural CFI / RMSEA / SRMR | Scalar CFI / RMSEA / SRMR | Scalar additional evidence |
|---|---|---|---|
| Intake restricted comparison | 0.967 / 0.093 / 0.057 | 0.974 / 0.070 / 0.057 | Test: 3.15 (6) / P=.79 |
| Assessment by baseline comparison | 0.971 / 0.092 / 0.056 | 0.977 / 0.070 / 0.056 | Test: 5.41 (6) / P=.49 |
| Omnibus gender within diagnosed | 0.971 / 0.093 / 0.054 | 0.974 / 0.070 / 0.055 | Test: 60.82 (14) / P<.001 |
| Age within diagnosed | 0.968 / 0.096 / 0.055 | 0.974 / 0.073 / 0.057 | Test: 29.14 (7) / P<.001 |
| Omnibus race within diagnosed | 0.968 / 0.096 / 0.058 | 0.974 / 0.069 / 0.058 | Test: 19.02 (12) / P=.09 |
| ADHD code within diagnosed | 0.972 / 0.091 / 0.053 | 0.976 / 0.071 / 0.054 | Test: 32.74 (7) / P<.001 |
| Hispanic or Latine vs not within diagnosed | 0.968 / 0.097 / 0.055 | 0.974 / 0.073 / 0.055 | Δ fit: 0.0038 / −0.0091 / −0.0001 |
| Diagnosed vs no assessment record | 0.974 / 0.089 / 0.052 | 0.978 / 0.067 / 0.052 | Δ fit: 0.0023 / −0.0071 / 0.0001 |
| Diagnosed plus prior assessment vs assessed comparison | 0.970 / 0.095 / 0.054 | 0.976 / 0.071 / 0.054 | Δ fit: 0.0040 / −0.0096 / 0.0000 |

| Comparison | Configural CFI / RMSEA / SRMR | Scalar CFI / RMSEA / SRMR | Scalar additional evidence |
|---|---|---|---|
| Broader service cohort vs assessed comparison | 0.970 / 0.095 / 0.054 | 0.976 / 0.071 / 0.054 | Δ fit: 0.0039 / −0.0094 / 0.0000 |
| Men vs women | 0.970 / 0.094 / 0.054 | 0.973 / 0.074 / 0.055 | Δ fit: 0.0011 / −0.0054 / 0.0006 |
| Gender-diverse vs women | 0.970 / 0.092 / 0.054 | 0.974 / 0.071 / 0.055 | Δ fit: 0.0020 / −0.0065 / 0.0002 |
| Under 35 vs 35 or older | 0.968 / 0.096 / 0.055 | 0.974 / 0.073 / 0.056 | Δ fit: 0.0025 / −0.0072 / 0.0010 |
| Black vs White | 0.968 / 0.096 / 0.056 | 0.973 / 0.073 / 0.057 | Δ fit: 0.0038 / −0.0089 / 0.0001 |
| Multiracial vs White | 0.967 / 0.097 / 0.058 | 0.973 / 0.073 / 0.058 | Δ fit: −0.0006 / −0.0023 / −0.0004 |
| Anxiety code present vs absent within diagnosed | 0.971 / 0.094 / 0.054 | 0.977 / 0.069 / 0.054 | Δ fit: 0.0037 / −0.0092 / 0.0001 |
| GAD-7 at least 10 vs below 10 | 0.947 / 0.091 / 0.067 | 0.944 / 0.079 / 0.069 | Δ fit: −0.0092 / 0.0030 / 0.0019 |

Original omnibus gender, age, race, and ADHD sequences: largest CFI decrease .0015, largest SRMR increase .0016. Age intercept SRMR was .057 (original sequence) and .056 (additional sequence).

The threshold-step SRMR permutation P was .030 despite a change rounding to zero. Other chi-square-change permutation values were .33, .344, and .840. No loading- or scalar-step fit-change flags occurred in the balanced subsamples.

Partial-invariance checks released the item 9 loading for age (P=.08), concentration first threshold and loading for ADHD-code status (P=.34), and three constraints for gender-diverse versus women (P=.15). The Black-versus-White pairwise model changed from P=.03 to .09 after releasing the first appetite threshold; multiracial versus White required no release (P=.75). These data-guided pairwise checks differ from the omnibus sequences.

**Panel C. Replication cohort subgroup scalar tests and adjacent fit changes**

| Comparison | $\Delta\chi^2$ (df) | P | ΔCFI | ΔRMSEA | ΔSRMR |
|---|---|---|---|---|---|
| Men vs women | 106.801 (7) | <.001 | 0.0005 | −0.0046 | 0.0005 |
| Gender-diverse vs women | 32.705 (7) | <.001 | 0.0022 | −0.0065 | 0.0003 |
| Under 35 vs 35 or older | 79.141 (7) | <.001 | 0.0018 | −0.0062 | 0.0011 |
| Black vs White | 19.132 (7) | .008 | 0.0035 | −0.0081 | 0.0000 |
| Multiracial vs White | 9.979 (7) | .190 | 0.0045 | −0.0096 | 0.0000 |
| Hispanic or Latine vs not | 20.817 (7) | .004 | 0.0035 | −0.0081 | 0.0001 |
| ADHD code vs no code | 28.019 (7) | <.001 | 0.0031 | −0.0076 | 0.0000 |
| Omnibus gender group | 134.657 (14) | <.001 | 0.0003 | −0.0049 | 0.0007 |
| Omnibus race group | 27.252 (14) | .018 | 0.0043 | −0.0098 | 0.0001 |

Changes compare scalar with loading constraints.

#### DIF screening

**Table S12.** DIF screening

##### Panel A. Screening results by comparison

| Comparison | BH flagged items | Holm flagged items | Anchor items |
|---|---|---|---|
| Diagnosed plus prior assessment vs assessed comparison | None | None | 4, 6, 7 |
| Broader service cohort vs assessed comparison | None | None | 2, 4, 6 |
| Diagnosed vs assessed comparison | None | None | 4, 6, 7 |
| Assessment outcome by baseline | None | None | 3, 5, 7 |
| Diagnosed vs no assessment record | 1, 2, 6, 9 | 1, 2, 6, 9 | 3, 5, 8 |
| ADHD code present vs absent within diagnosed | 7 | 7 | 1, 5, 6 |
| Anxiety code present vs absent within diagnosed | None | None | 1, 2, 6 |
| GAD-7 at least 10 vs below 10 | 1, 2, 5, 6, 7, 8, 9 | 1, 2, 6, 7, 8, 9 | None |
| Black vs White | None | None | 6, 7, 8 |
| Gender-diverse vs women | 2, 4, 5, 9 | 2 | 1, 6, 8 |
| Men vs women | 3, 4, 5 | 3, 4, 5 | None |
| Multiracial vs White | None | None | 6, 7, 8 |
| Replication: diagnosed vs assessed comparison | 2, 4, 8 | 8 | 3, 6, 7 |
| Under 35 vs 35 or older | 9 | 9 | 2, 3, 4 |

Items are numbered 1–9. Replication men versus women had no eligible three-item anchor set (Table S19).

##### Panel B. Original ethnicity comparison, final DIF screen for all nine items

| Item | LR $\chi^2$ | df | P | BH P | Holm P |
|---|---|---|---|---|---|
| 1 | 4.262 | 4 | .372 | .478 | 1.000 |
| 2 | 2.249 | 4 | .690 | .776 | 1.000 |
| 3 | 9.248 | 4 | .055 | .248 | .442 |
| 4 | 4.614 | 4 | .329 | .478 | 1.000 |
| 5 | 5.178 | 4 | .269 | .478 | 1.000 |
| 6 | 7.227 | 4 | .124 | .373 | .871 |
| 7 | 1.255 | 4 | .869 | .869 | 1.000 |
| 8 | 22.305 | 4 | <.001 | .002 | .002 |
| 9 | 4.930 | 3 | .177 | .398 | 1.000 |

Item 8 was the only flagged item; BH and Holm P=.002.

##### Panel C. Final within-diagnosed BH flags in both cohorts

| Comparison | Original items | Count / 9 | Replication cohort items | Count / 9 |
|---|---|---|---|---|
| Men vs women | 3, 4, 5 | 3 | 3, 4, 5, 7 | 4 |
| Gender-diverse vs women | 2, 4, 5, 9 | 4 | 9 | 1 |
| Under 35 vs 35 or older | 9 | 1 | 3, 5, 8, 9 | 4 |
| Black vs White | None | 0 | None | 0 |

| Comparison | Original items | Count / 9 | Replication cohort items | Count / 9 |
|---|---|---|---|---|
| Multiracial vs White | None | 0 | None | 0 |
| Hispanic or Latine vs not | 8 | 1 | 3, 8 | 2 |
| ADHD code vs no code | 7 | 1 | 7 | 1 |

Totals: original 10/63; replication 12/63 (BH within each nine-item contrast).

#### Expected item score effects

**Table S13.** Item-level expected-score effects

##### Panel A. Original assessment comparison

| Item | Signed (95% CI) | Unsigned (95% CI) | Fixed |
|---|---|---|---|
| 1 Anhedonia | −0.017 (−0.094 to 0.052) | 0.021 (0.016 to 0.103) | No |
| 2 Depressed mood | −0.044 (−0.118 to 0.025) | 0.053 (0.018 to 0.126) | No |
| 3 Sleep | 0.037 (−0.058 to 0.122) | 0.070 (0.020 to 0.154) | No |
| 4 Fatigue | 0.000 (0.000 to 0.000) | 0.000 (0.000 to 0.000) | Yes |
| 5 Appetite | 0.011 (−0.082 to 0.102) | 0.025 (0.011 to 0.114) | No |
| 6 Worthlessness | 0.000 (0.000 to 0.000) | 0.000 (0.000 to 0.000) | Yes |
| 7 Concentration | 0.000 (0.000 to 0.000) | 0.000 (0.000 to 0.000) | Yes |
| 8 Psychomotor change | 0.080 (0.004 to 0.151) | 0.084 (0.036 to 0.154) | No |
| 9 Death or self-harm thoughts | 0.010 (−0.038 to 0.048) | 0.024 (0.007 to 0.062) | No |

##### Panel B. Replication cohort assessment comparison

| Item | Signed (95% CI) | Unsigned (95% CI) | Fixed |
|---|---|---|---|
| 1 Anhedonia | −0.069 (−0.119 to −0.020) | 0.069 (0.027 to 0.121) | No |
| 2 Depressed mood | −0.079 (−0.131 to −0.031) | 0.079 (0.036 to 0.131) | No |
| 3 Sleep | 0.000 (0.000 to 0.000) | 0.000 (0.000 to 0.000) | Yes |
| 4 Fatigue | 0.035 (−0.017 to 0.090) | 0.035 (0.011 to 0.092) | No |
| 5 Appetite | 0.033 (−0.027 to 0.092) | 0.048 (0.016 to 0.104) | No |
| 6 Worthlessness | 0.000 (0.000 to 0.000) | 0.000 (0.000 to 0.000) | Yes |
| 7 Concentration | 0.000 (0.000 to 0.000) | 0.000 (0.000 to 0.000) | Yes |
| 8 Psychomotor change | 0.098 (0.047 to 0.145) | 0.098 (0.050 to 0.145) | No |
| 9 Death or self-harm thoughts | −0.015 (−0.054 to 0.019) | 0.036 (0.016 to 0.067) | No |

##### Panel C. Selected item effects across contrasts

| Item | Signed difference | CI lower | CI upper (level) |
|---|---|---|---|
| **Gender-diverse vs women** | | | |
| Fatigue | −0.131 | −0.221 | −0.040 (95%) |
| Item 9 | 0.057 | 0.015 | 0.104 (95%) |
| Depressed mood | 0.051 | −0.016 | 0.117 (95%) |
| Appetite | −0.055 | −0.146 | 0.043 (95%) |
| **Under 35 vs 35 or older** | | | |
| Item 9 | 0.071 | 0.038 | 0.102 (95%) |
| **ADHD code present vs absent** | | | |

| Item | Signed difference | CI lower | CI upper (level) |
|---|---|---|---|
| Concentration | 0.186 | 0.103 | 0.266 (95%) |
| Item 9 | −0.060 | −0.092 | −0.021 (95%) |
| **Replication cohort: diagnosed vs assessed comparison** | | | |
| Depressed mood | −0.079 | −0.131 | −0.031 (95%) |
| Psychomotor change | 0.098 | 0.047 | 0.145 (95%) |
| Fatigue | 0.035 | −0.017 | 0.090 (95%) |

Fixed anchors have zero effect by construction.

##### Panel D. Replication cohort largest unsigned item effect under each fitted link

| Comparison and link | Item | Signed point effect | Unsigned point effect |
|---|---|---|---|
| ADHD code vs no code; original anchors | 7 | 0.155 | 0.155 |
| ADHD code vs no code; selected anchors | 7 | 0.153 | 0.153 |
| Black vs White; original anchors | 5 | 0.097 | 0.097 |
| Black vs White; selected anchors | 6 | −0.116 | 0.116 |
| Gender-diverse vs women; original anchors | 9 | 0.088 | 0.088 |
| Gender-diverse vs women; selected anchors | 9 | 0.097 | 0.097 |
| Hispanic or Latine vs not; original anchors | 8 | 0.097 | 0.108 |
| Hispanic or Latine vs not; selected anchors | 8 | 0.076 | 0.090 |
| Men vs women; items 1 and 7 | 4 | −0.245 | 0.245 |
| Men vs women; item 1 only | 4 | −0.282 | 0.282 |
| Men vs women; item 7 only | 2 | 0.185 | 0.185 |
| Multiracial vs White; original anchors | 5 | 0.167 | 0.167 |
| Multiracial vs White; selected anchors | 6 | −0.114 | 0.114 |
| Under 35 vs 35 or older; original anchors | 9 | 0.083 | 0.083 |
| Under 35 vs 35 or older; selected anchors | 5 | 0.083 | 0.086 |

**Table S13, continued.** Item effects for additional original-cohort models

##### Original-cohort item effects

| Original-cohort comparison and item | Signed estimate | 95% CI |
|---|---|---|
| **Gender-diverse vs women** | | |
| 1 | 0.000 | 0.000 to 0.000 |
| 2 | 0.051 | −0.016 to 0.117 |
| 3 | −0.023 | −0.121 to 0.068 |
| 4 | −0.131 | −0.221 to −0.040 |
| 5 | −0.055 | −0.146 to 0.043 |
| 6 | 0.000 | 0.000 to 0.000 |
| 7 | −0.115 | −0.210 to −0.024 |
| 8 | 0.000 | 0.000 to 0.000 |
| 9 | 0.057 | 0.015 to 0.104 |
| **Under 35 vs 35 or older** | | |
| 1 | −0.003 | −0.048 to 0.042 |
| 2 | 0.000 | 0.000 to 0.000 |
| 3 | 0.000 | 0.000 to 0.000 |
| 4 | 0.000 | 0.000 to 0.000 |

| Original-cohort comparison and item | Signed estimate | 95% CI |
|---|---|---|
| 5 | 0.073 | 0.010 to 0.135 |
| 6 | −0.015 | −0.072 to 0.036 |
| 7 | −0.035 | −0.097 to 0.025 |
| 8 | 0.080 | 0.026 to 0.133 |
| 9 | 0.071 | 0.038 to 0.102 |
| **Black vs White** | | |
| 1 | 0.088 | −0.002 to 0.179 |
| 2 | −0.022 | −0.107 to 0.067 |
| 3 | 0.035 | −0.083 to 0.146 |
| 4 | −0.094 | −0.191 to 0.005 |
| 5 | 0.169 | 0.055 to 0.283 |
| 6 | 0.000 | 0.000 to 0.000 |
| 7 | 0.000 | 0.000 to 0.000 |
| 8 | 0.000 | 0.000 to 0.000 |
| 9 | −0.022 | −0.055 to 0.017 |
| **Multiracial vs White** | | |
| 1 | 0.036 | −0.074 to 0.151 |
| 2 | −0.034 | −0.136 to 0.083 |
| 3 | 0.104 | −0.023 to 0.235 |
| 4 | 0.027 | −0.104 to 0.157 |
| 5 | 0.061 | −0.079 to 0.196 |
| 6 | 0.000 | 0.000 to 0.000 |
| 7 | 0.000 | 0.000 to 0.000 |
| 8 | 0.000 | 0.000 to 0.000 |
| 9 | 0.003 | −0.040 to 0.058 |
| **ADHD code vs no code** | | |
| 1 | 0.000 | 0.000 to 0.000 |
| 2 | −0.044 | −0.102 to 0.017 |
| 3 | 0.030 | −0.053 to 0.109 |
| 4 | −0.050 | −0.127 to 0.018 |
| 5 | 0.000 | 0.000 to 0.000 |
| 6 | 0.000 | 0.000 to 0.000 |
| 7 | 0.186 | 0.103 to 0.266 |
| 8 | 0.013 | −0.056 to 0.079 |
| 9 | −0.060 | −0.092 to −0.021 |

#### Integrated score differences

**Table S14.** Integrated score differences

##### Panel A. Original-cohort and selected-model estimates

| Comparison | Signed estimate and CI | Original-scale signed envelope | Original-scale unsigned upper |
|---|---|---|---|
| Diagnosed vs assessed comparison | 0.077 (95% CI −0.207 to 0.325) | −0.249 to 0.363 (95%) | 0.371 |

| Comparison | Signed estimate and CI | Original-scale signed envelope | Original-scale unsigned upper |
|---|---|---|---|
| Diagnosed vs no assessment record | −0.643 (95% CI −0.978 to −0.320) | −0.978 to −0.320 (95%) | 0.920 |
| Diagnosed plus prior assessment vs assessed comparison | 0.086 (95% CI −0.187 to 0.346) | −0.229 to 0.384 (95%) | 0.380 |
| Broader service cohort vs assessed comparison | 0.021 (95% CI −0.228 to 0.243) | −0.271 to 0.281 (95%) | 0.355 |
| Gender-diverse vs women | −0.216 (95% CI −0.498 to 0.064) | −0.534 to 0.110 (95%) | 0.501 |
| Under 35 vs 35 or older | 0.170 (95% CI −0.007 to 0.343) | −0.007 to 0.343 (95%) | 0.318 |
| Black vs White | 0.154 (95% CI −0.201 to 0.517) | −0.504 to 0.799 (95%) | 0.782 |
| Multiracial vs White | 0.198 (95% CI −0.231 to 0.622) | −0.538 to 0.975 (95%) | 0.931 |
| ADHD code present vs absent within diagnosed | 0.076 (95% CI −0.160 to 0.307) | −0.202 to 0.331 (95%) | 0.330 |
| Anxiety code present vs absent within diagnosed | 0.025 (95% CI −0.157 to 0.203) | −0.157 to 0.203 (95%) | 0.366 |
| Assessment outcome by baseline | −0.022 (95% CI −0.471 to 0.365) | −0.997 to 0.743 (95%) | 0.956 |
| Replication: diagnosed vs assessed comparison | 0.002 (95% CI −0.187 to 0.201) | −0.187 to 0.201 (95%) | 0.226 |
| Hispanic or Latine vs not within diagnosed | 0.065 (−0.114 to 0.257) [90%] | −0.151 to 0.303 (90%) | 0.531 |
| Men vs women (two eligible anchors) | −0.383 (−0.608 to −0.146) [90%] | −0.644 to −0.100 (90%) | 0.649 |

Focal minus reference. Signed intervals specify their coverage; unsigned upper bounds are one-sided 95%. Intervals use cohort-specific item 9 coding; common-coding values for the same anchor sets are in Table S16, Panel C. Rows labeled [90%] and the replication subgroup estimates use central 90% intervals, as originally specified; these are narrower than 95% intervals and should not be compared with them directly.

#### Panel B. Replication cohort-selected three-item subgroup estimates

| Comparison | Focal / reference n | Anchors | Signed (90% CI) | Original signed envelope | Unsigned upper |
|---|---|---|---|---|---|
| Gender-diverse vs women | 826 / 4,946 | 1 8 5 | 0.079 (−0.095 to 0.266) | −0.095 to 0.266 | 0.383 |
| Under 35 vs 35 or older | 3,353 / 4,051 | 2 6 4 | 0.185 (0.089 to 0.284) | 0.089 to 0.284 | 0.284 |
| Black vs White | 840 / 5,276 | 8 3 5 | −0.280 (−0.487 to −0.059) | −0.487 to −0.059 | 0.506 |
| Multiracial vs White | 417 / 5,276 | 7 4 2 | 0.076 (−0.133 to 0.304) | −0.171 to 0.345 | 0.372 |
| Hispanic or Latine vs not | 1,151 / 6,052 | 4 6 7 | 0.155 (0.005 to 0.312) | 0.005 to 0.312 | 0.315 |
| ADHD code vs no code | 831 / 4,924 | 5 4 1 | 0.245 (0.083 to 0.417) | 0.083 to 0.417 | 0.420 |

Multiracial comparison pooled item 9; other replication pairs used four categories.

##### Panel C. Replication cohort fixed-original-anchor subgroup sensitivities

| Comparison | Focal / reference n | Anchors | Signed (90% CI) | Original signed envelope | Unsigned upper |
|---|---|---|---|---|---|
| Gender-diverse vs women | 826 / 4,946 | 1 6 8 | −0.018 (−0.183 to 0.154) | −0.183 to 0.154 | 0.308 |
| Under 35 vs 35 or older | 3,353 / 4,051 | 2 3 4 | 0.158 (0.056 to 0.259) | 0.056 to 0.259 | 0.259 |
| Black vs White | 840 / 5,276 | 6 7 8 | 0.287 (0.086 to 0.488) | 0.086 to 0.488 | 0.491 |
| Multiracial vs White | 417 / 5,276 | 6 7 8 | 0.474 (0.213 to 0.747) | 0.176 to 0.794 | 0.807 |
| Hispanic or Latine vs not | 1,151 / 6,052 | 1 2 7 | 0.332 (0.199 to 0.469) | 0.199 to 0.469 | 0.474 |
| ADHD code vs no code | 831 / 4,924 | 1 5 6 | 0.257 (0.096 to 0.422) | 0.096 to 0.422 | 0.426 |

Multiracial comparison pooled item 9; other replication pairs used four categories.

##### Panel D. Latent distribution in the linked graded response models

| Cohort | Group | n | Mean | Variance | SD |
|---|---|---|---|---|---|
| Original | Assessed comparison | 484 | 0.000 | 1.000 | 1.000 |
| Original | Diagnosed | 3,471 | 0.175 | 0.935 | 0.967 |
| Replication cohort | Assessed comparison | 1,194 | 0.000 | 1.000 | 1.000 |
| Replication cohort | Diagnosed | 7,406 | 0.144 | 1.112 | 1.055 |

Reference means and variances were fixed at 0 and 1.

#### Anchors, coding, and draws

**Table S15.** Anchors, coding, and simulation draws

##### Panel A. Selected anchors and draws

| Comparison | Anchor items | Draws in selection run |
|---|---|---|
| Diagnosed vs assessed comparison | 4, 6, 7 | 8,000 |
| Diagnosed vs no assessment record | 3, 5, 8 | 16,000 |
| Diagnosed plus prior assessment vs assessed comparison | 4, 6, 7 | 8,000 |
| Broader service cohort vs assessed comparison | 2, 4, 6 | 8,000 |
| Gender-diverse vs women | 1, 6, 8 | 16,000 |
| Under 35 vs 35 or older | 2, 3, 4 | 8,000 |
| Black vs White | 6, 7, 8 | 32,000 |
| Multiracial vs White | 6, 7, 8 | 64,000 |
| ADHD code present vs absent within diagnosed | 1, 5, 6 | 8,000 |
| Anxiety code present vs absent within diagnosed | 1, 2, 6 | 8,000 |
| Assessment outcome by baseline | 3, 5, 7 | 32,000 (2 × 16,000) |
| Replication: diagnosed vs assessed comparison | 3, 6, 7 | 2,000 |

#### Panel B. Additional comparison anchors and draws

| Comparison | Anchor items | N reference / focal | Draws |
|---|---|---|---|
| Hispanic or Latine vs not within diagnosed | 7, 2, 1 | 2,773 / 554 | 4,000 |

#### Panel C. Category maps

| Comparison | Coding differing from 0 / 1 / 2 / 3 |
|---|---|
| Diagnosed plus prior assessment vs assessed comparison | Item 9 0 / 1 / 2–3 |
| Broader service cohort vs assessed comparison | Item 9 0 / 1 / 2–3 |
| Diagnosed vs assessed comparison | Item 9 0 / 1 / 2–3 |
| Assessment outcome by baseline | Item 8 0 / 1 / 2–3; Item 9 0 / 1–3 |
| Diagnosed vs no assessment record | All items 0 / 1 / 2 / 3 |
| ADHD code present vs absent within diagnosed | Item 9 0 / 1 / 2–3 |
| Anxiety code present vs absent within diagnosed | All items 0 / 1 / 2 / 3 |
| GAD-7 at least 10 vs below 10 | Item 9 0 / 1 / 2–3 |
| Black vs White | Item 9 0 / 1–3 |
| Gender-diverse vs women | Item 9 0 / 1 / 2–3 |
| Men vs women | Item 9 0 / 1 / 2–3 |
| Multiracial vs White | Item 9 0 / 1–3 |
| Replication: diagnosed vs assessed comparison | All items 0 / 1 / 2 / 3 |
| Under 35 vs 35 or older | All items 0 / 1 / 2 / 3 |

Multigroup linked graded response models with the anchor set selected by iterative purification: original C2=1,218.90 (64 df), RMSEA .068 (.064–.071), CFI .953, TLI .947 (maximum endpoint gap .008); replication full-category model C2=3,021.72 (64 df), RMSEA .073 (.071–.076), CFI .938, TLI .931. The men–women joint model used 8,000 draws; timing checks used two batches of 16,000.

#### Panel D. Ordinal CFA category maps by subgroup comparison

| Comparison | Original coding exception | Replication cohort coding exception |
|---|---|---|
| Men vs women | All items: 0 / 1 / 2 / 3 | All items: 0 / 1 / 2 / 3 |
| Gender-diverse vs women | item 9: 0 / 1 / 2–3 | All items: 0 / 1 / 2 / 3 |
| Under 35 vs 35 or older | All items: 0 / 1 / 2 / 3 | All items: 0 / 1 / 2 / 3 |
| Black vs White | item 9: 0 / 1 / 2–3 | All items: 0 / 1 / 2 / 3 |
| Multiracial vs White | item 9: 0 / 1–3 | item 9: 0 / 1 / 2–3 |
| Hispanic or Latine vs not | item 9: 0 / 1 / 2–3 | All items: 0 / 1 / 2 / 3 |
| ADHD code vs no code | item 9: 0 / 1 / 2–3 | All items: 0 / 1 / 2 / 3 |
| Omnibus gender | item 9: 0 / 1 / 2–3 | All items: 0 / 1 / 2 / 3 |
| Omnibus race | item 9: 0 / 1–3 | item 9: 0 / 1 / 2–3 |

Except where shown, items retain 0/1/2/3.

#### Panel E. GRM category maps by subgroup comparison

| Comparison | Original coding exception | Replication cohort coding exception |
|---|---|---|
| Men vs women | item 9: 0 / 1 / 2–3 | All items: 0 / 1 / 2 / 3 |
| Gender-diverse vs women | item 9: 0 / 1 / 2–3 | All items: 0 / 1 / 2 / 3 |
| Under 35 vs 35 or older | All items: 0 / 1 / 2 / 3 | All items: 0 / 1 / 2 / 3 |
| Black vs White | item 9: 0 / 1–3 | All items: 0 / 1 / 2 / 3 |
| Multiracial vs White | item 9: 0 / 1–3 | item 9: 0 / 1 / 2–3 |

| Comparison | Original coding exception | Replication cohort coding exception |
|---|---|---|
| Hispanic or Latine vs not | item 9: 0 / 1 / 2–3 | All items: 0 / 1 / 2 / 3 |
| ADHD code vs no code | item 9: 0 / 1 / 2–3 | All items: 0 / 1 / 2 / 3 |

**Panel F. Replication cohort parameter-draw stability**

| Comparison | Anchors | Total draws | Endpoint gap | Stable |
|---|---|---|---|---|
| Men vs women | 1 | 8,000 | 0.0069 | Yes |
| Men vs women | 7 | 32,000 | 0.0059 | Yes |
| Men vs women | 1 and 7 | 8,000 | 0.0035 | Yes |
| Gender-diverse vs women | 1 6 8 | 8,000 | 0.0054 | Yes |
| Gender-diverse vs women | 1 8 5 | 8,000 | 0.0041 | Yes |
| Under 35 vs 35 or older | 2 3 4 | 8,000 | 0.0023 | Yes |
| Under 35 vs 35 or older | 2 6 4 | 8,000 | 0.0061 | Yes |
| Black vs White | 6 7 8 | 8,000 | 0.0093 | Yes |
| Black vs White | 8 3 5 | 8,000 | 0.0055 | Yes |
| Multiracial vs White | 6 7 8 | 8,000 | 0.0049 | Yes |
| Multiracial vs White | 7 4 2 | 8,000 | 0.0025 | Yes |
| Hispanic or Latine vs not | 1 2 7 | 8,000 | 0.0095 | Yes |
| Hispanic or Latine vs not | 4 6 7 | 8,000 | 0.0048 | Yes |
| ADHD code vs no code | 1 5 6 | 8,000 | 0.0037 | Yes |
| ADHD code vs no code | 5 4 1 | 8,000 | 0.0094 | Yes |

#### All three-item anchor sets

All 56 common-coding fits in each cohort converged. Panel A gives point estimates under each cohort's native coding. Panel C gives common-coding estimates and central 95% intervals conditional on the corresponding model. The intervals are model-specific and have no simultaneous coverage interpretation.

**Table S16.** All three-item anchor sets

**Panel A. All corresponding anchor sets**

| Anchor items | Original signed | Replication cohort signed | Original unsigned envelope | Replication cohort screen flag in set |
|---|---|---|---|---|
| 3, 4, 8 | −0.458 | −0.472 | 0.502 | Yes |
| 3, 5, 8 | −0.347 | −0.474 | 0.388 | Yes |
| 3, 6, 8 | −0.323 | −0.207 | 0.367 | Yes |
| 4, 5, 8 | −0.314 | −0.444 | 0.352 | Yes |
| 4, 6, 8 | −0.296 | −0.215 | 0.334 | Yes |
| 5, 6, 8 | −0.215 | −0.193 | 0.251 | Yes |
| 3, 4, 5 | −0.211 | −0.228 | 0.261 | Yes |
| 3, 4, 6 | −0.200 | −0.057 | 0.262 | Yes |
| 1, 3, 8 | −0.155 | 0.023 | 0.193 | Yes |

| Anchor items | Original signed | Replication cohort signed | Original unsigned envelope | Replication cohort screen flag in set |
|---|---|---|---|---|
| 1, 4, 8 | −0.153 | 0.002 | 0.187 | Yes |
| 3, 7, 8 | −0.104 | −0.447 | 0.145 | Yes |
| 4, 7, 8 | −0.103 | −0.428 | 0.138 | Yes |
| 4, 5, 6 | −0.097 | −0.050 | 0.134 | Yes |
| 3, 5, 6 | −0.095 | −0.032 | 0.182 | No |
| 1, 6, 8 | −0.092 | 0.166 | 0.130 | Yes |
| 1, 5, 8 | −0.082 | 0.021 | 0.169 | Yes |
| 1, 3, 4 | −0.065 | 0.129 | 0.117 | Yes |
| 2, 4, 8 | −0.046 | 0.071 | 0.094 | Yes |
| 2, 3, 8 | −0.022 | 0.097 | 0.233 | Yes |
| 6, 7, 8 | −0.014 | −0.156 | 0.136 | Yes |
| 3, 4, 7 | −0.005 | −0.211 | 0.156 | Yes |
| 1, 4, 6 | −0.003 | 0.233 | 0.082 | Yes |
| 1, 3, 6 | 0.003 | 0.261 | 0.143 | Yes |
| 5, 7, 8 | 0.005 | −0.418 | 0.197 | Yes |
| 1, 4, 5 | 0.010 | 0.123 | 0.115 | Yes |
| 2, 3, 4 | 0.019 | 0.178 | 0.260 | Yes |
| 1, 3, 5 | 0.021 | 0.145 | 0.099 | Yes |
| 2, 6, 8 | 0.034 | 0.233 | 0.129 | Yes |
| 2, 5, 8 | 0.045 | 0.097 | 0.123 | Yes |
| 1, 5, 6 | 0.067 | 0.257 | 0.145 | Yes |
| 4, 6, 7 | 0.077 | −0.025 | 0.137 | Yes |
| 2, 4, 6 | 0.082 | 0.281 | 0.185 | Yes |
| 2, 4, 5 | 0.091 | 0.175 | 0.180 | Yes |
| 1, 7, 8 | 0.092 | 0.064 | 0.215 | Yes |
| 3, 6, 7 | 0.094 | 0.002 | 0.209 | No |
| 4, 5, 7 | 0.097 | −0.197 | 0.166 | Yes |
| 1, 2, 8 | 0.098 | 0.324 | 0.147 | Yes |
| 2, 3, 6 | 0.105 | 0.312 | 0.306 | Yes |
| 2, 3, 5 | 0.120 | 0.200 | 0.282 | Yes |
| 3, 5, 7 | 0.125 | −0.188 | 0.195 | No |
| 1, 2, 4 | 0.144 | 0.370 | 0.198 | Yes |
| 2, 5, 6 | 0.165 | 0.309 | 0.232 | Yes |
| 1, 4, 7 | 0.166 | 0.155 | 0.220 | Yes |
| 1, 2, 3 | 0.166 | 0.397 | 0.271 | Yes |
| 5, 6, 7 | 0.175 | 0.007 | 0.228 | No |
| 1, 3, 7 | 0.186 | 0.185 | 0.230 | Yes |
| 1, 2, 6 | 0.202 | 0.484 | 0.249 | Yes |
| 1, 2, 5 | 0.215 | 0.391 | 0.258 | Yes |
| 2, 7, 8 | 0.216 | 0.140 | 0.262 | Yes |
| 1, 6, 7 | 0.218 | 0.294 | 0.262 | Yes |
| 2, 4, 7 | 0.244 | 0.207 | 0.297 | Yes |
| 1, 5, 7 | 0.245 | 0.178 | 0.306 | Yes |
| 2, 3, 7 | 0.283 | 0.240 | 0.381 | Yes |
| 2, 6, 7 | 0.314 | 0.347 | 0.361 | Yes |
| 2, 5, 7 | 0.337 | 0.236 | 0.379 | Yes |

| Anchor items | Original signed | Replication cohort signed | Original unsigned envelope | Replication cohort screen flag in set |
|---|---|---|---|---|
| 1, 2, 7 | 0.354 | 0.428 | 0.393 | Yes |

#### Panel B. Domain composition

| Somatic anchors | Original median and range | Replication cohort median and range |
|---|---|---|
| 0 (4 sets) | 0.266 (0.202 to 0.354) | 0.387 (0.294 to 0.484) |
| 1 (24 sets) | 0.125 (−0.092 to 0.337) | 0.233 (−0.156 to 0.397) |
| 2 (24 sets) | −0.055 (−0.323 to 0.125) | −0.015 (−0.447 to 0.200) |
| 3 (4 sets) | −0.331 (−0.458 to −0.211) | −0.458 (−0.474 to −0.228) |

Somatic items are 3, 4, 5, and 8. Fitted range under native coding is 0–26 (original) and 0–27 (replication). Selected sets were 4/6/7 (original) and 3/6/7 (replication); 52/56 replication sets contained an item flagged during purification or final screening. Signed differences are diagnosed minus assessed comparison.

#### Panel C. Common-coding enumeration (item 9 pooled in both cohorts)

| Anchor items | Original signed | Replication signed | Original unsigned envelope | Replication unsigned envelope | Original 95% CI | Replication 95% CI |
|---|---|---|---|---|---|---|
| 3, 4, 8 | −0.46 | −0.46 | 0.502 | 0.508 | −0.80 to −0.12 | −0.68 to −0.25 |
| 3, 5, 8 | −0.35 | −0.47 | 0.388 | 0.517 | −0.68 to −0.04 | −0.68 to −0.26 |
| 3, 6, 8 | −0.32 | −0.20 | 0.367 | 0.245 | −0.64 to −0.03 | −0.40 to 0.00 |
| 4, 5, 8 | −0.31 | −0.44 | 0.352 | 0.480 | −0.62 to −0.01 | −0.64 to −0.24 |
| 4, 6, 8 | −0.30 | −0.21 | 0.334 | 0.287 | −0.59 to −0.02 | −0.41 to −0.02 |
| 5, 6, 8 | −0.21 | −0.19 | 0.251 | 0.242 | −0.53 to 0.08 | −0.39 to 0.01 |
| 3, 4, 5 | −0.21 | −0.22 | 0.261 | 0.292 | −0.53 to 0.08 | −0.43 to −0.04 |
| 3, 4, 6 | −0.20 | −0.05 | 0.262 | 0.110 | −0.50 to 0.07 | −0.24 to 0.12 |
| 1, 3, 8 | −0.16 | 0.03 | 0.193 | 0.120 | −0.45 to 0.11 | −0.15 to 0.20 |
| 1, 4, 8 | −0.15 | 0.01 | 0.187 | 0.071 | −0.44 to 0.10 | −0.17 to 0.18 |
| 3, 7, 8 | −0.10 | −0.44 | 0.145 | 0.483 | −0.47 to 0.23 | −0.66 to −0.21 |
| 4, 7, 8 | −0.10 | −0.42 | 0.138 | 0.471 | −0.41 to 0.21 | −0.63 to −0.21 |
| 4, 5, 6 | −0.10 | −0.05 | 0.134 | 0.115 | −0.36 to 0.16 | −0.22 to 0.12 |
| 3, 5, 6 | −0.10 | −0.03 | 0.182 | 0.104 | −0.38 to 0.17 | −0.21 to 0.15 |
| 1, 6, 8 | −0.09 | 0.17 | 0.130 | 0.210 | −0.35 to 0.17 | −0.01 to 0.34 |
| 1, 5, 8 | −0.08 | 0.02 | 0.169 | 0.094 | −0.36 to 0.18 | −0.14 to 0.19 |
| 1, 3, 4 | −0.06 | 0.13 | 0.117 | 0.188 | −0.34 to 0.18 | −0.03 to 0.28 |
| 2, 4, 8 | −0.05 | 0.07 | 0.094 | 0.126 | −0.31 to 0.22 | −0.09 to 0.24 |
| 2, 3, 8 | −0.02 | 0.10 | 0.233 | 0.139 | −0.31 to 0.25 | −0.08 to 0.27 |
| 6, 7, 8 | −0.01 | −0.15 | 0.136 | 0.282 | −0.33 to 0.28 | −0.35 to 0.05 |
| 3, 4, 7 | 0.00 | −0.20 | 0.156 | 0.255 | −0.32 to 0.28 | −0.41 to −0.02 |
| 1, 4, 6 | 0.00 | 0.24 | 0.082 | 0.274 | −0.26 to 0.23 | 0.07 to 0.39 |
| 1, 3, 6 | 0.00 | 0.26 | 0.143 | 0.302 | −0.26 to 0.25 | 0.10 to 0.41 |
| 5, 7, 8 | 0.00 | −0.41 | 0.197 | 0.454 | −0.31 to 0.31 | −0.63 to −0.19 |
| 1, 4, 5 | 0.01 | 0.13 | 0.115 | 0.171 | −0.25 to 0.26 | −0.04 to 0.28 |
| 2, 3, 4 | 0.02 | 0.18 | 0.260 | 0.219 | −0.24 to 0.28 | 0.03 to 0.33 |
| 1, 3, 5 | 0.02 | 0.15 | 0.099 | 0.254 | −0.24 to 0.27 | −0.01 to 0.30 |
| 2, 6, 8 | 0.03 | 0.24 | 0.129 | 0.293 | −0.24 to 0.30 | 0.07 to 0.40 |
| 2, 5, 8 | 0.05 | 0.10 | 0.123 | 0.138 | −0.24 to 0.30 | −0.08 to 0.27 |

| Anchor items | Original signed | Replication signed | Original unsigned envelope | Replication unsigned envelope | Original 95% CI | Replication 95% CI |
|---|---|---|---|---|---|---|
| 1, 5, 6 | 0.07 | 0.26 | 0.145 | 0.298 | −0.19 to 0.30 | 0.10 to 0.40 |
| 4, 6, 7 | 0.08 | −0.02 | 0.137 | 0.160 | −0.20 to 0.34 | −0.19 to 0.14 |
| 2, 4, 6 | 0.08 | 0.28 | 0.185 | 0.323 | −0.16 to 0.31 | 0.13 to 0.43 |
| 2, 4, 5 | 0.09 | 0.18 | 0.180 | 0.216 | −0.16 to 0.33 | 0.02 to 0.33 |
| 1, 7, 8 | 0.09 | 0.07 | 0.215 | 0.111 | −0.19 to 0.34 | −0.11 to 0.24 |
| 3, 6, 7 | 0.09 | 0.01 | 0.209 | 0.091 | −0.20 to 0.38 | −0.17 to 0.18 |
| 4, 5, 7 | 0.10 | −0.19 | 0.166 | 0.245 | −0.20 to 0.37 | −0.39 to −0.01 |
| 1, 2, 8 | 0.10 | 0.33 | 0.147 | 0.365 | −0.16 to 0.34 | 0.17 to 0.48 |
| 2, 3, 6 | 0.10 | 0.32 | 0.306 | 0.353 | −0.16 to 0.35 | 0.16 to 0.47 |
| 2, 3, 5 | 0.12 | 0.20 | 0.282 | 0.247 | −0.15 to 0.36 | 0.05 to 0.36 |
| 3, 5, 7 | 0.12 | −0.18 | 0.195 | 0.254 | −0.19 to 0.41 | −0.39 to 0.01 |
| 1, 2, 4 | 0.14 | 0.37 | 0.198 | 0.410 | −0.10 to 0.37 | 0.22 to 0.51 |
| 2, 5, 6 | 0.16 | 0.31 | 0.232 | 0.351 | −0.09 to 0.40 | 0.16 to 0.46 |
| 1, 4, 7 | 0.17 | 0.16 | 0.220 | 0.196 | −0.09 to 0.41 | −0.01 to 0.31 |
| 1, 2, 3 | 0.17 | 0.40 | 0.271 | 0.437 | −0.08 to 0.39 | 0.24 to 0.55 |
| 5, 6, 7 | 0.18 | 0.01 | 0.228 | 0.112 | −0.12 to 0.42 | −0.18 to 0.19 |
| 1, 3, 7 | 0.19 | 0.19 | 0.230 | 0.226 | −0.07 to 0.42 | 0.03 to 0.35 |
| 1, 2, 6 | 0.20 | 0.49 | 0.249 | 0.524 | −0.04 to 0.43 | 0.34 to 0.63 |
| 1, 2, 5 | 0.22 | 0.39 | 0.258 | 0.431 | −0.04 to 0.44 | 0.25 to 0.54 |
| 2, 7, 8 | 0.22 | 0.14 | 0.262 | 0.201 | −0.05 to 0.49 | −0.04 to 0.33 |
| 1, 6, 7 | 0.22 | 0.30 | 0.262 | 0.335 | −0.02 to 0.46 | 0.13 to 0.45 |
| 2, 4, 7 | 0.24 | 0.21 | 0.297 | 0.248 | 0.00 to 0.48 | 0.04 to 0.36 |
| 1, 5, 7 | 0.24 | 0.18 | 0.306 | 0.219 | 0.00 to 0.48 | 0.02 to 0.34 |
| 2, 3, 7 | 0.28 | 0.24 | 0.381 | 0.281 | 0.02 to 0.52 | 0.07 to 0.40 |
| 2, 6, 7 | 0.31 | 0.35 | 0.361 | 0.388 | 0.05 to 0.55 | 0.20 to 0.51 |
| 2, 5, 7 | 0.34 | 0.24 | 0.379 | 0.277 | 0.08 to 0.58 | 0.07 to 0.39 |
| 1, 2, 7 | 0.35 | 0.43 | 0.393 | 0.469 | 0.12 to 0.59 | 0.28 to 0.57 |

Item 9 responses 2 and 3 pooled in both cohorts (fitted range 0–26). Unsigned envelopes are evaluated on the original 0–27 scale at fitted parameters without draws; four exceeded 0.50 (original: items 3, 4, 8; replication: items 3, 4, 8; 3, 5, 8; and 1, 2, 6). These envelopes are the upper bounds reported in the main text. Table S14 reports one-sided 95% upper bounds from parameter draws; the envelopes here are evaluated at the fitted parameters without draws, so the two are not comparable (for items 4, 6, and 7 in the original cohort: 0.371 in Table S14 and 0.137 here).

#### Targeted alternative anchors

##### Panel D. Item-membership contrasts across all anchor sets

| Anchor item | Original contrast | Replication cohort contrast |
|---|---|---|
| 1 | 0.084 | 0.258 |
| 2 | 0.201 | 0.325 |
| 3 | −0.106 | −0.106 |
| 4 | −0.118 | −0.126 |
| 5 | −0.013 | −0.101 |
| 6 | −0.025 | 0.077 |
| 7 | 0.188 | −0.063 |

| Anchor item | Original contrast | Replication cohort contrast |
|---|---|---|
| 8 | −0.211 | −0.264 |

Contrast = mean estimate in the 21 sets containing the item minus the mean in the 35 sets excluding it.

**Table S17.** Targeted alternative anchors

| Comparison | Fits | Signed point range | Unsigned point range |
|---|---|---|---|
| Diagnosed vs assessed comparison | 5 | −0.281 to 0.650 | 0.085 to 0.654 |
| Diagnosed vs no assessment record | 3 | −0.876 to −0.502 | 0.513 to 0.876 |
| Diagnosed plus prior assessment vs assessed comparison | 5 | −0.268 to 0.655 | 0.076 to 0.659 |
| Broader service cohort vs assessed comparison | 5 | −0.177 to 0.220 | 0.130 to 0.302 |
| Men vs women | 2 | −0.619 to 0.086 | 0.091 to 0.623 |
| Gender-diverse vs women | 5 | −0.354 to 0.079 | 0.128 to 0.522 |
| Under 35 vs 35 or older | 5 | −0.026 to 0.295 | 0.057 to 0.295 |
| Black vs White | 5 | −0.750 to 0.401 | 0.136 to 0.752 |
| Multiracial vs White | 5 | −0.225 to 0.530 | 0.041 to 0.572 |
| ADHD code present vs absent within diagnosed | 5 | −0.055 to 0.219 | 0.058 to 0.273 |
| Anxiety code present vs absent within diagnosed | 5 | −0.128 to 0.081 | 0.133 to 0.329 |
| Assessment outcome by baseline | 5 | −0.530 to 0.844 | 0.262 to 0.874 |
| Replication: diagnosed vs assessed comparison | 5 | −0.214 to 0.261 | 0.077 to 0.342 |

Values are ranges of targeted point fits. Original single-anchor signed estimates: −0.281 (item 4), −0.028 (item 6), 0.650 (item 7); alternative triples gave −0.200 and 0.094.

#### Item 9 analyses

**Table S18.** Item 9 analyses

##### Panel A. Binary endorsement model

| Cohort | PHQ-8 | Odds ratio (95% CI) | Risk difference, percentage points (95% CI) |
|---|---|---|---|
| Original | 5 | 1.42 (0.77 to 2.61) | 1.2 (−1.1 to 2.8) |
| Original | 10 | 1.24 (0.85 to 1.81) | 2.1 (−1.8 to 5.1) |
| Original | 15 | 1.08 (0.76 to 1.54) | 1.6 (−5.9 to 8.2) |
| Replication cohort | 5 | 0.66 (0.49 to 0.88) | −2.2 (−4.2 to −0.6) |
| Replication cohort | 10 | 0.85 (0.70 to 1.04) | −1.8 (−4.2 to 0.4) |
| Replication cohort | 15 | 1.11 (0.89 to 1.37) | 2.0 (−2.4 to 6.1) |
| Pooled | 5 | 0.78 (0.60 to 1.02) | −1.2 (−2.7 to 0.1) |
| Pooled | 10 | 0.92 (0.78 to 1.10) | −0.9 (−2.9 to 1.0) |
| Pooled | 15 | 1.09 (0.91 to 1.31) | 1.7 (−2.0 to 5.3) |

##### Panel B. Standardized item 9 loadings (post hoc; identical coding in both cohorts)

| Cohort | Assessed comparison loading (standardized) | Diagnosed loading (standardized) | Standardized difference (diagnosed minus assessed comparison) and 95% CI | Unstandardized difference and 95% CI |
|---|---|---|---|---|
| Original | 0.787 | 0.708 | −0.078 (−0.177 to 0.021) | −0.154 (−0.573 to 0.264) |

| Cohort | Assessed comparison loading (standardized) | Diagnosed loading (standardized) | Standardized difference (diagnosed minus assessed comparison) and 95% CI | Unstandardized difference and 95% CI |
|---|---|---|---|---|
| Replication | 0.573 | 0.709 | 0.136 (0.058 to 0.215) | 0.204 (0.015 to 0.394) |
| Difference between cohorts (replication minus original, from unrounded values) | — | — | 0.215 (0.088 to 0.341), P<.001 | 0.359 (−0.101 to 0.818), P=.13 |

Ordinal two-factor models with item 9 responses 2 and 3 pooled and thresholds constrained across groups; scaled RMSEA 0.090 (original) and 0.096 (replication).

##### Panel C. Raw item 9 endorsement at PHQ-8=5 and within PHQ-8 scores 3–7 (post hoc)

| Cohort | Group | PHQ-8 window | Clients, n | Endorsers (responses 1–3), n | Nonendorsers (response 0), n | Endorsement, % |
|---|---|---|---|---|---|---|
| Original | Diagnosed | 5 | 234 | 7 | 227 | 3.0 |
| Original | Assessed comparison | 5 | 32 | 2 | 30 | 6.3 |
| Original | Diagnosed | 3–7 | 1,149 | 45 | 1,104 | 3.9 |
| Original | Assessed comparison | 3–7 | 182 | 6 | 176 | 3.3 |
| Replication | Diagnosed | 5 | 458 | 13 | 445 | 2.8 |
| Replication | Assessed comparison | 5 | 95 | 6 | 89 | 6.3 |
| Replication | Diagnosed | 3–7 | 2,401 | 96 | 2,305 | 4.0 |
| Replication | Assessed comparison | 3–7 | 444 | 24 | 420 | 5.4 |

Counts use the first eligible paired administration; PHQ-8 is the sum of items 1–8.

##### Panel D. Group-by-PHQ-8 interaction and between-cohort tests (post hoc except the pooled test)

| Test or contrast | Ratio of odds ratios per 5 PHQ-8 points (95% CI) | Ratio of odds ratios per 1 PHQ-8 point (95% CI) | Statistic and P |
|---|---|---|---|
| Original interaction (1 df) | 0.87 (0.63 to 1.20) | 0.973 (0.912 to 1.037) | $\chi^2(1)=0.74$, P=.39 |
| Replication interaction (1 df) | 1.30 (1.09 to 1.55) | 1.054 (1.018 to 1.091) | $\chi^2(1)=8.41$, P=.004 |
| Group and group × PHQ-8 terms together (2 df) | — | — | Original $\chi^2(2)=1.44$, P=.49; replication $\chi^2(2)=8.76$, P=.01 |
| Pooled heterogeneity (cohort × group and cohort × group × PHQ-8) | — | — | $\chi^2(2)=5.78$, P=.06 |
| Score-5 contrast, original divided by replication (exploratory) | 2.17 (1.10 to 4.27) | — | Wald P=.03 |

McFadden $\Delta R^2$ for the two group terms was 0.0004 (original) and 0.0012 (replication).

##### Panel E. Post hoc sensitivity models for the odds ratio at PHQ-8=5, 10, and 15

| PHQ-8 | Model | Original: OR (95% CI) | Replication: OR (95% CI) |
|---|---|---|---|
| 5 | Linear, maximum likelihood (primary) | 1.42 (0.77 to 2.61) | 0.66 (0.49 to 0.88) |
| 5 | Quadratic PHQ-8 term | 1.20 (0.59 to 2.42) | 0.61 (0.42 to 0.87) |
| 5 | Natural spline | 1.21 (0.58 to 2.50) | 0.60 (0.41 to 0.87) |
| 5 | Linear, Firth penalized likelihood | 1.38 (0.78 to 2.61) | 0.65 (0.49 to 0.88) |
| 10 | Linear, maximum likelihood (primary) | 1.24 (0.85 to 1.81) | 0.85 (0.70 to 1.04) |

| PHQ-8 | Model | Original: OR (95% CI) | Replication: OR (95% CI) |
|---|---|---|---|
| 10 | Quadratic PHQ-8 term | 1.33 (0.87 to 2.05) | 0.83 (0.65 to 1.04) |
| 10 | Natural spline | 1.20 (0.70 to 2.07) | 0.82 (0.62 to 1.08) |
| 10 | Linear, Firth penalized likelihood | 1.22 (0.85 to 1.81) | 0.85 (0.70 to 1.03) |
| 15 | Linear, maximum likelihood (primary) | 1.08 (0.76 to 1.54) | 1.11 (0.89 to 1.37) |
| 15 | Quadratic PHQ-8 term | 1.19 (0.79 to 1.80) | 1.12 (0.88 to 1.43) |
| 15 | Natural spline | 1.31 (0.79 to 2.18) | 1.14 (0.83 to 1.57) |
| 15 | Linear, Firth penalized likelihood | 1.08 (0.76 to 1.53) | 1.10 (0.89 to 1.37) |

Quadratic term $\chi^2(2)=7.26$, P=.03 (original) and 17.43, P<.001 (replication); natural spline $\chi^2(4)=9.54$, P=.05 and 19.06, P<.001. Firth models used logistf 1.26.1 with penalized profile-likelihood intervals.

**Panel F. Average fitted differences in item 9 endorsement probability (exploratory)**

| Cohort | Scope | Average signed difference, percentage points (95% CI) | Average absolute difference, percentage points (95% CI) | Diagnosed clients represented, % |
|---|---|---|---|---|
| Original | Adequate local support | 1.35 (−2.18 to 4.48) | 1.56 (0.48 to 4.63) | 88.6 |
| Original | Shared observed range | 1.22 (−2.03 to 4.13) | 1.49 (0.47 to 4.37) | 100.0 |
| Replication | Adequate local support | −0.19 (−2.55 to 1.98) | 2.61 (1.14 to 4.59) | 98.2 |
| Replication | Shared observed range | −0.22 (−2.54 to 1.92) | 2.60 (1.13 to 4.56) | 100.0 |

Supported scores ranged from 3 to 22 (original) and 1 to 24 (replication), covering 88.6% and 98.2% of diagnosed clients; intervals use 20,000 coefficient draws.

**Panel G. Repeat item 9 endorsement among baseline endorsers (descriptive)**

| Cohort | Group | Baseline endorsers, n | With a second eligible submission, n (%) | Endorsed again, n/N (%) | Median days between administrations (IQR) |
|---|---|---|---|---|---|
| Original | Diagnosed | 531 | 279 (52.5) | 153/279 (54.8) | 32 (28–39) |
| Original | Assessed comparison | 58 | 29 (50.0) | 17/29 (58.6) | 33 (29–37) |
| Replication | Diagnosed | 1,172 | 646 (55.1) | 304/646 (47.1) | 31 (29–37) |
| Replication | Assessed comparison | 171 | 85 (49.7) | 36/85 (42.4) | 31 (29–39) |

Baseline endorsers endorsed item 9 (responses 1–3) at the first administration; the denominator is those with a second eligible paired submission.

Original logistic reference-group OR per PHQ-8 point was 1.284 (1.208–1.364), and interaction OR .973 (.912–1.037). Original reference/diagnosed predicted probabilities were .030/.042 at 5, .098/.119 at 10, and .275/.290 at 15. Replication cohort interaction OR was 1.054 (1.018–1.091). Original endorsement categories 1/2/3 contained 405/83/43 diagnosed and 45/7/6 assessed comparison clients. The pooled heterogeneity test was $\chi^2=5.780$ (2 df), P=.06. Replication cohort item 9 categories 0/1/2/3 contained 6,234/894/178/100 diagnosed clients and

1,023/135/20/16 assessed comparison clients; any endorsement occurred in 1,172/7,406 (15.83%) and 171/1,194 (14.32%).

Pooled ordinal diagnostics rejected proportional PHQ-8 effects under both original and collapsed coding (both P<.001). Under collapsed coding, the group-by-PHQ-8 term also departed from proportionality (P<.001). Follow-up models therefore allowed group, PHQ-8, and interaction effects to vary across thresholds; the remaining cohort proportionality tests were nonsignificant (P=.89 and P=.75). Original-coding group and group-by-PHQ-8 term-specific diagnostic fits remained unresolved. In the linked graded response models, the item 9 signed expected-score effect was 0.010 modeled points (90% CI −0.031 to 0.044) in the original comparison, with unsigned effect 0.024 (0.010 to 0.058). Replication cohort gave signed effect −0.015 (−0.048 to 0.014) and unsigned effect 0.036 (0.020 to 0.063). These were empirical-distribution-integrated, anchor-conditional effects; the original item used pooled upper categories and the replication-cohort item used all four categories.

The assessment-by-baseline standardized difference was −0.041 (−0.222 to 0.140). Continuous-model dMACS estimates were 0.028 (0.012-0.194) in the original cohort, 0.098 (0.048-0.182) in the replication cohort, and 0.122 (0.027-0.409) in the timing subset.

**Panel H. Item 9 observed endorsement counts**

| Cohort | Group | n | Response 1 | Response 2 | Response 3 | Any endorsement, n (%) |
|---|---|---|---|---|---|---|
| Original | Diagnosed | 3,471 | 405 | 83 | 43 | 531 (15.30) |
| Original | Assessed comparison | 484 | 45 | 7 | 6 | 58 (11.98) |
| Replication cohort | Diagnosed | 7,406 | 894 | 178 | 100 | 1,172 (15.83) |
| Replication cohort | Assessed comparison | 1,194 | 135 | 20 | 16 | 171 (14.32) |

Any endorsement is responses 1–3 versus 0.

#### Men and women anchor models

**Table S19.** Men-versus-women anchor models

##### Panel A. Original cohort: single-anchor estimates

| Single anchor | Signed (90% CI) | Original signed envelope | Original unsigned upper |
|---|---|---|---|
| Item 1 | −0.619 (−0.902 to −0.315) | −0.936 to −0.271 | 0.944 |
| Item 7 | 0.086 (−0.382 to 0.546) | −0.417 to 0.597 | 0.668 |

##### Panel B. Original cohort: joint-anchor estimates

| Quantity | Value (8,000 draws) |
|---|---|
| Signed impact (PHQ-9 points) | −0.383 |
| Signed 90% CI lower | −0.608 |
| Signed 90% CI upper | −0.146 |
| Unsigned impact | 0.384 |
| Unsigned 90% CI lower | 0.187 |
| Unsigned 90% CI upper | 0.613 |
| Original-scale signed envelope, lower | −0.644 |
| Original-scale signed envelope, upper | −0.100 |
| Original-scale unsigned upper bound | 0.649 |
| Maximum endpoint gap between batches | 0.009 |
| Endpoint rule (gap ≤ 0.01) | Met |

##### Panel C. Original cohort: nested likelihood-ratio tests

| Freed item (other item held equal) | Likelihood-ratio $\chi^2$ | df | P |
|---|---|---|---|
| Item 7 (item 1 equal) | 5.97 | 4 | .202 |
| Item 1 (item 7 equal) | 5.19 | 4 | .268 |

##### Panel D. Original cohort: information criteria

| Model | AIC | BIC |
|---|---|---|
| Joint anchors 1 and 7 | 53095.4 | 53478.2 |
| Item 1 anchor only | 53097.5 | 53504.1 |
| Item 7 anchor only | 53098.3 | 53504.9 |

N=2,167 women and 755 men. Joint-model C2=894.29 (60 df), RMSEA .069 (90% CI .065–.073), CFI .954; two 4,000-draw batches agreed within .009. Intervals are central 90%, as originally specified.

##### Panel E. Replication cohort men–women fixed-link estimates

| Anchors | Signed (90% CI) | Unsigned upper | Total draws |
|---|---|---|---|
| 1 | −0.647 (−0.847 to −0.446) | 0.850 | 8,000 |
| 7 | 0.460 (0.118 to 0.799) | 0.802 | 32,000 |
| 1 and 7 | −0.305 (−0.466 to −0.145) | 0.471 | 8,000 |

Women n=4,946; men n=1,557. Fixed-link checks used 8,000 draws (item 1 and joint) and 32,000 (item 7).

#### Timing, composition, and coding checks

**Table S20.** Timing, composition, and coding

##### Panel A. Effects of the timing, composition, and coding checks

| Analysis | Anchor items | Signed estimate and CI | Original-scale unsigned upper bound |
|---|---|---|---|
| Women only (2,167 vs 350) | 4, 6, 7 | −0.054 (−0.347 to 0.204) [90%] | 0.428 |
| Pooled cohorts (10,877 vs 1,678), item 9 four categories | 4, 6, 7 | 0.007 (−0.120 to 0.129) [90%] | 0.198 |
| Pooled cohorts, second anchor set | 3, 6, 7 | 0.032 (−0.097 to 0.161) [90%] | 0.200 |
| Replication cohort (7,406 vs 1,194), item 9 pooled | 3, 6, 7 | 0.007 (−0.156 to 0.156) [90%] | 0.262 |
| Timing-restricted (2,047 vs 225) | 3, 5, 7 | −0.022 (95% CI −0.471 to 0.365) | 0.956 |

##### Panel B. Numerical diagnostics

| Analysis | Endpoint gap | Draws per batch | RMSEA (C2) |
|---|---|---|---|
| Women only (2,167 vs 350) | 0.008 | 2,000 | 0.067 |
| Pooled cohorts (10,877 vs 1,678), item 9 four categories | 0.008 | 2,000 | 0.071 |
| Pooled cohorts, second anchor set | 0.010 | 2,000 | 0.071 |
| Replication cohort (7,406 vs 1,194), item 9 pooled | 0.007 | 2,000 | 0.073 |
| Timing-restricted stability run (2,047 vs 225) | 0.004 | 16,000 | 0.065 |

Pooled cohorts: 10,877 diagnosed and 1,678 assessed comparison clients. Timing-restricted: 2,047 and 225 clients, original means 8.85 and 7.41, difference 1.43 (95% CI 0.71–2.16), g=0.253. Intake-only: 2,219 and 290 clients. Intervals are central 90%, as originally specified, except the timing-restricted row (95%).

##### Panel C. Replication cohort primary and pooled-category linked-model fit

| Model | C2 (df) | RMSEA | 95% RMSEA CI | CFI |
|---|---|---|---|---|
| Replication cohort primary four categories | 3021.719 (64) | 0.073 | 0.071 to 0.076 | 0.938 |
| Replication cohort pooled item 9 sensitivity | 3035.829 (64) | 0.073 | 0.071 to 0.076 | 0.938 |

The pooled-category row changes response coding under fixed anchors 3/6/7.

#### Detection simulations

**Table S21.** Detection simulations

##### Panel A. Detection of injected differences by test step

| Condition | Label error | Test step | Detected, % (Monte Carlo SE) | 95% CI | Fit-change flag, % |
|---|---|---|---|---|---|
| Loading increase 25% (total-score shift 0.09) | None | Loadings | 87.8 (1.5) | 84.6 to 90.4 | 11.4 |
| Loading increase 25% (total-score shift 0.09) | 2% symmetric | Loadings | 80.2 (1.8) | 76.5 to 83.5 | 9.2 |

| Condition | Label error | Test step | Detected, % (Monte Carlo SE) | 95% CI | Fit-change flag, % |
|---|---|---|---|---|---|
| Loading increase 25% (total-score shift 0.09) | 11.8% / 18.8% | Loadings | 44.0 (2.2) | 39.7 to 48.4 | 0.8 |
| Loading increase 25% (total-score shift 0.09) | 21.5% / 35.3% | Loadings | 18.4 (1.7) | 15.2 to 22.0 | 0.2 |
| Threshold shift 0.30 SD (total-score shift −0.51) | None | Intercepts | 100.0 (0.0) | 99.2 to 100.0 | 96.4 |
| Threshold shift 0.30 SD (total-score shift −0.51) | 2% symmetric | Intercepts | 100.0 (0.0) | 99.2 to 100.0 | 91.6 |
| Threshold shift 0.30 SD (total-score shift −0.51) | 11.8% / 18.8% | Intercepts | 99.8 (0.2) | 98.9 to 100.0 | 44.6 |
| Threshold shift 0.30 SD (total-score shift −0.51) | 21.5% / 35.3% | Intercepts | 74.0 (2.0) | 70.0 to 77.7 | 2.0 |
| Threshold shift 0.15 SD (total-score shift −0.26) | 11.8% / 18.8% | Intercepts | 53.8 (2.2) | 49.4 to 58.1 | 0.8 |
| No injected difference | None | Thresholds | 5.0 (1.0) | 3.4 to 7.3 | 0.0 |
| No injected difference | None | Loadings | 5.6 (1.0) | 3.9 to 8.0 | 0.2 |
| No injected difference | None | Intercepts | 3.8 (0.9) | 2.4 to 5.9 | 0.0 |
| No injected difference | 11.8% / 18.8% | Thresholds | 4.8 (1.0) | 3.2 to 7.0 | 0.0 |
| No injected difference | 11.8% / 18.8% | Loadings | 5.4 (1.0) | 3.7 to 7.7 | 0.0 |
| No injected difference | 11.8% / 18.8% | Intercepts | 5.0 (1.0) | 3.4 to 7.3 | 0.0 |

**Panel B. Threshold-step diagnostics**

| Condition | Label error | Test step | Detected, % (Monte Carlo SE) | 95% CI | Fit-change flag, % |
|---|---|---|---|---|---|
| Threshold shift 0.30 SD | None | thresholds | 4.4 (0.9) | 2.9 to 6.6 | 0.0 |
| Threshold shift 0.30 SD | 2% symmetric | thresholds | 6.8 (1.1) | 4.9 to 9.4 | 0.0 |
| Threshold shift 0.30 SD | 11.8% / 18.8% | thresholds | 4.0 (0.9) | 2.6 to 6.1 | 0.0 |
| Threshold shift 0.30 SD | 21.5% / 35.3% | thresholds | 3.8 (0.9) | 2.4 to 5.9 | 0.0 |
| Threshold shift 0.15 SD | 11.8% / 18.8% | thresholds | 3.4 (0.8) | 2.1 to 5.4 | 0.0 |

500 replicates per condition; injections affected items 2 and 5; modeled total-score shifts were −.51 (.30 SD), −.26 (.15 SD), and +.09 (loading increase).

#### Additional clinical comparisons

These supplementary clinical correlates and service-cohort checks are distinct analyses. Recorded codes are not treated as independent depression diagnoses.

**Table S22.** Additional clinical comparisons

**Panel A. Diagnosed clinical code mean contrasts**

| Code and timing sample | N with code / total | Mean with / without and Hedges g |
|---|---|---|
| Depressive code F32/F33; all clients with codes | 954 / 2,951 | 11.02 / 8.49; g 0.45 |

| Code and timing sample | N with code / total | Mean with / without and Hedges g |
|---|---|---|
| Depressive code F32/F33; earliest plan on or before baseline | 364 / 1,078 | 9.89 / 7.44; g 0.44 |
| Depressive code F32/F33 or dysthymia F34.1; all clients with codes | 978 / 2,951 | 11.01 / 8.46; g 0.45 |
| Depressive code F32/F33 or dysthymia F34.1; earliest plan on or before baseline | 372 / 1,078 | 9.84 / 7.44; g 0.43 |
| GAD code F41.1 (exploratory); all clients with codes | 1,512 / 2,951 | 9.55 / 9.05; g 0.09 |
| GAD code F41.1 (exploratory); earliest plan on or before baseline | 606 / 1,078 | 8.54 / 7.91; g 0.11 |
| Any anxiety code (exploratory); all clients with codes | 1,752 / 2,951 | 9.44 / 9.11; g 0.06 |
| Any anxiety code (exploratory); earliest plan on or before baseline | 691 / 1,078 | 8.50 / 7.84; g 0.12 |

**Panel B. Assessed comparison clinical code mean contrasts**

| Code and timing sample | N with code / total | Mean with / without and Hedges g |
|---|---|---|
| Depressive code F32/F33; all clients with codes | 134 / 405 | 10.25 / 7.30; g 0.55 |
| Depressive code F32/F33; earliest plan on or before baseline | 55 / 170 | 8.07 / 6.58; g 0.30 |
| Depressive code F32/F33 or dysthymia F34.1; all clients with codes | 136 / 405 | 10.32 / 7.24; g 0.57 |
| Depressive code F32/F33 or dysthymia F34.1; earliest plan on or before baseline | 57 / 170 | 8.32 / 6.43; g 0.38 |
| GAD code F41.1 (exploratory); all clients with codes | 209 / 405 | 8.29 / 8.26; g 0.01 |
| GAD code F41.1 (exploratory); earliest plan on or before baseline | 98 / 170 | 6.88 / 7.32; g −0.09 |
| Any anxiety code (exploratory); all clients with codes | 245 / 405 | 8.41 / 8.06; g 0.06 |
| Any anxiety code (exploratory); earliest plan on or before baseline | 111 / 170 | 6.95 / 7.29; g −0.07 |

Broader-cohort alpha was .856 (adding prior-assessment clients) and .857 (also adding not-completed and no-record clients; GRM reliability .861). Signed total effects were 0.086 and 0.021 (Table S14). The response-pattern screen flagged 241/5,077 profiles (4.7%); observed mean contrasts were 0.99 (flagged excluded), 0.88 (all-zero excluded), and 0.95 (principal analysis).

Response flags comprised 148 of 3,471 diagnosed and 17 of 484 assessed comparison clients (165 of 3,955 in the primary comparison); the separate all-zero count was 148 of 5,077 in the full original cohort (90 diagnosed and 18 assessed comparison). Excluding flags left 3,323 and 467 clients (means 9.17/8.19); excluding all-zero responses left 3,381 and 466 (9.64/8.76). The broader 4,591-client cohort had alpha .857, GRM reliability .861, one-factor omega .904, and two-factor total omega .911; the 3,512-client diagnosed-plus-prior-assessment cohort had .856, .861, .905, and .912.

#### Domain factor means

**Table S23.** Domain factor means

**Panel A. Domain factor means**

| Quantity | Estimate | SE | 95% CI |
|---|---|---|---|
| Cognitive-affective factor | −0.137 | 0.056 | −0.247 to −0.028 |
| Somatic factor | −0.208 | 0.057 | −0.319 to −0.097 |
| Somatic minus cognitive-affective | −0.071 | 0.047 | −0.164 to 0.022 |

Assessed comparison relative to diagnosed (reference), in diagnosed latent SD units; signs are opposite to the diagnosed-minus-assessed convention (cognitive-affective +0.137, somatic +0.208, difference +0.071).

#### Recorded depressive-disorder codes

**Table S24.** Recorded depressive-disorder codes

| Cohort and assessment group | Clients, n | AUC (DeLong 95% CI) | Sensitivity at ≥10 (95% CI) | Specificity at ≥10 (95% CI) | Code positive, n/N (%) |
|---|---|---|---|---|---|
| Original: diagnosed | 2,951 | 0.628 (0.606 to 0.649) | 526/954 (55.1; 51.9–58.3) | 1,261/1,997 (63.1; 61.0–65.3) | 954/2,951 (32.3) |
| Original: assessed comparison | 405 | 0.650 (0.593 to 0.707) | 65/134 (48.5; 39.8–57.3) | 196/271 (72.3; 66.6–77.6) | 134/405 (33.1) |
| Replication: diagnosed | 5,755 | 0.647 (0.631 to 0.662) | 906/1,482 (61.1; 58.6–63.6) | 2,574/4,273 (60.2; 58.8–61.7) | 1,482/5,755 (25.8) |
| Replication: assessed comparison | 863 | 0.598 (0.555 to 0.641) | 118/229 (51.5; 44.9–58.2) | 409/634 (64.5; 60.6–68.2) | 229/863 (26.5) |

Code positive = F32/F33-series code in any position on any available treatment plan (F34.1 excluded; remission codes included; no date restriction). Sensitivity and specificity use exact binomial intervals. Code status was available for 9,974 of 12,555 clients.

#### Supplementary figure

**Figure S1.** Test information and standard error. Replication cohort curves from the single-group graded response models; the horizontal axis is the model-expected PHQ-9 total and the standard error is in latent severity units.

#### Item response frequencies and polychoric correlations

**Table S25.** Item response distributions and polychoric correlations

Each client contributed the first eligible paired administration. The tabulated frequencies combine responses 2 and 3 on item 9 in both cohorts. The descriptive polychoric matrices use the original four response categories, with the marginal cumulative proportions determining the fixed thresholds.

**Original cohort, diagnosed (n=3,471): item distribution**

| Item | 0 | 1 | 2 (2–3 for item 9) | 3 |
|---|---|---|---|---|
| 1 | 1,004 | 1,592 | 562 | 313 |
| 2 | 1,085 | 1,601 | 479 | 306 |
| 3 | 730 | 1,240 | 706 | 795 |
| 4 | 383 | 1,318 | 822 | 948 |
| 5 | 1,222 | 1,102 | 603 | 544 |
| 6 | 1,255 | 1,307 | 503 | 406 |
| 7 | 936 | 1,258 | 663 | 614 |
| 8 | 2,111 | 882 | 295 | 183 |
| 9 | 2,940 | 405 | 126 | |

**Original cohort, diagnosed (n=3,471): polychoric correlations**

| Item | 1 | 2 | 3 | 4 | 5 | 6 | 7 | 8 | 9 |
|---|---|---|---|---|---|---|---|---|---|
| 1 | 1.000 | | | | | | | | |
| 2 | 0.761 | 1.000 | | | | | | | |
| 3 | 0.494 | 0.456 | 1.000 | | | | | | |
| 4 | 0.601 | 0.572 | 0.612 | 1.000 | | | | | |
| 5 | 0.506 | 0.500 | 0.514 | 0.559 | 1.000 | | | | |
| 6 | 0.595 | 0.720 | 0.388 | 0.478 | 0.470 | 1.000 | | | |
| 7 | 0.538 | 0.480 | 0.411 | 0.493 | 0.459 | 0.433 | 1.000 | | |
| 8 | 0.440 | 0.426 | 0.390 | 0.364 | 0.422 | 0.454 | 0.522 | 1.000 | |
| 9 | 0.561 | 0.663 | 0.335 | 0.392 | 0.448 | 0.621 | 0.385 | 0.425 | 1.000 |

**Original cohort, assessed comparison (n=484): item distribution**

| Item | 0 | 1 | 2 (2–3 for item 9) | 3 |
|---|---|---|---|---|
| 1 | 164 | 223 | 54 | 43 |
| 2 | 170 | 216 | 62 | 36 |
| 3 | 120 | 186 | 86 | 92 |
| 4 | 66 | 209 | 108 | 101 |
| 5 | 184 | 168 | 73 | 59 |
| 6 | 198 | 184 | 53 | 49 |
| 7 | 144 | 167 | 89 | 84 |
| 8 | 344 | 86 | 36 | 18 |
| 9 | 426 | 45 | 13 | |

**Original cohort, assessed comparison (n=484): polychoric correlations**

| Item | 1 | 2 | 3 | 4 | 5 | 6 | 7 | 8 | 9 |
|---|---|---|---|---|---|---|---|---|---|
| 1 | 1.000 | | | | | | | | |
| 2 | 0.734 | 1.000 | | | | | | | |
| 3 | 0.464 | 0.365 | 1.000 | | | | | | |
| 4 | 0.632 | 0.547 | 0.593 | 1.000 | | | | | |
| 5 | 0.578 | 0.530 | 0.477 | 0.613 | 1.000 | | | | |
| 6 | 0.606 | 0.770 | 0.342 | 0.513 | 0.520 | 1.000 | | | |
| 7 | 0.609 | 0.454 | 0.368 | 0.536 | 0.515 | 0.480 | 1.000 | | |
| 8 | 0.565 | 0.414 | 0.445 | 0.487 | 0.503 | 0.445 | 0.512 | 1.000 | |
| 9 | 0.598 | 0.773 | 0.348 | 0.558 | 0.452 | 0.691 | 0.403 | 0.408 | 1.000 |

**Replication cohort, diagnosed (n=7,406): item distribution**

| Item | 0 | 1 | 2 (2–3 for item 9) | 3 |
|---|---|---|---|---|
| 1 | 2,151 | 3,289 | 1,189 | 777 |
| 2 | 2,266 | 3,447 | 1,034 | 659 |
| 3 | 1,399 | 2,618 | 1,651 | 1,738 |
| 4 | 698 | 2,795 | 1,713 | 2,200 |
| 5 | 2,590 | 2,400 | 1,296 | 1,120 |
| 6 | 2,589 | 2,758 | 1,152 | 907 |
| 7 | 1,728 | 2,714 | 1,474 | 1,490 |
| 8 | 4,414 | 1,843 | 732 | 417 |
| 9 | 6,234 | 894 | 278 | |

**Replication cohort, diagnosed (n=7,406): polychoric correlations**

| Item | 1 | 2 | 3 | 4 | 5 | 6 | 7 | 8 | 9 |
|---|---|---|---|---|---|---|---|---|---|
| 1 | 1.000 | | | | | | | | |
| 2 | 0.757 | 1.000 | | | | | | | |
| 3 | 0.443 | 0.422 | 1.000 | | | | | | |
| 4 | 0.569 | 0.514 | 0.589 | 1.000 | | | | | |
| 5 | 0.478 | 0.473 | 0.468 | 0.525 | 1.000 | | | | |
| 6 | 0.563 | 0.701 | 0.362 | 0.431 | 0.437 | 1.000 | | | |
| 7 | 0.481 | 0.437 | 0.408 | 0.489 | 0.426 | 0.426 | 1.000 | | |
| 8 | 0.412 | 0.406 | 0.354 | 0.381 | 0.431 | 0.412 | 0.506 | 1.000 | |
| 9 | 0.538 | 0.671 | 0.319 | 0.370 | 0.376 | 0.643 | 0.354 | 0.382 | 1.000 |

**Replication cohort, assessed comparison (n=1,194): item distribution**

| Item | 0 | 1 | 2 (2–3 for item 9) | 3 |
|---|---|---|---|---|
| 1 | 347 | 565 | 168 | 114 |
| 2 | 361 | 577 | 164 | 92 |
| 3 | 262 | 449 | 251 | 232 |
| 4 | 119 | 527 | 256 | 292 |
| 5 | 469 | 403 | 183 | 139 |
| 6 | 431 | 476 | 168 | 119 |
| 7 | 307 | 462 | 225 | 200 |
| 8 | 812 | 258 | 85 | 39 |
| 9 | 1,023 | 135 | 36 | |

**Replication cohort, assessed comparison (n=1,194): polychoric correlations**

| Item | 1 | 2 | 3 | 4 | 5 | 6 | 7 | 8 | 9 |
|---|---|---|---|---|---|---|---|---|---|
| 1 | 1.000 | | | | | | | | |
| 2 | 0.763 | 1.000 | | | | | | | |
| 3 | 0.462 | 0.436 | 1.000 | | | | | | |

| Item | 1 | 2 | 3 | 4 | 5 | 6 | 7 | 8 | 9 |
|---|---|---|---|---|---|---|---|---|---|
| 4 | 0.571 | 0.471 | 0.605 | 1.000 | | | | | |
| 5 | 0.505 | 0.481 | 0.476 | 0.506 | 1.000 | | | | |
| 6 | 0.543 | 0.672 | 0.333 | 0.401 | 0.440 | 1.000 | | | |
| 7 | 0.442 | 0.382 | 0.447 | 0.465 | 0.422 | 0.397 | 1.000 | | |
| 8 | 0.385 | 0.371 | 0.372 | 0.338 | 0.439 | 0.404 | 0.542 | 1.000 | |
| 9 | 0.409 | 0.574 | 0.286 | 0.235 | 0.335 | 0.533 | 0.267 | 0.320 | 1.000 |

**Table S26.** Item 9 local support counts

| Cohort | PHQ-8 target | PHQ-8 window (inclusive) | Assessment group | Clients in window, n | Item 9 endorsers (responses 1–3), n | Item 9 nonendorsers (response 0), n |
|---|---|---|---|---|---|---|
| Original | 5 | 3–7 | Diagnosed | 1,149 | 45 | 1,104 |
| Original | 5 | 3–7 | Assessed comparison | 182 | 6 | 176 |
| Original | 10 | 8–12 | Diagnosed | 1,055 | 125 | 930 |
| Original | 10 | 8–12 | Assessed comparison | 128 | 16 | 112 |
| Original | 15 | 13–17 | Diagnosed | 587 | 179 | 408 |
| Original | 15 | 13–17 | Assessed comparison | 75 | 19 | 56 |
| Replication | 5 | 3–7 | Diagnosed | 2,401 | 96 | 2,305 |
| Replication | 5 | 3–7 | Assessed comparison | 444 | 24 | 420 |
| Replication | 10 | 8–12 | Diagnosed | 2,324 | 308 | 2,016 |
| Replication | 10 | 8–12 | Assessed comparison | 377 | 62 | 315 |
| Replication | 15 | 13–17 | Diagnosed | 1,372 | 393 | 979 |
| Replication | 15 | 13–17 | Assessed comparison | 187 | 50 | 137 |

Support rule: at least five endorsers and five nonendorsers within two PHQ-8 points; all 12 cells met it (smallest endorser count 6).

#### Extended methods and results

The following sections provide additional detail on the methods and results summarized in the main manuscript.

#### Extended methods

##### Analytic frameworks and aims

The primary aim was to compare expected PHQ-9 totals by assessment outcome using item response theory. Secondary analyses addressed internal structure, reliability, precision, association with anxiety, and item 9 endorsement at the same PHQ-8 score. Subgroup comparisons within diagnosed clients were exploratory. The original cohort included 5,077 clients and 3,955 in the assessment-group comparison. The replication cohort from the same service included 10,579 clients and 8,600 in the comparison. Common-coding anchor enumeration was the main analysis of expected total-score differences. The study was not preregistered. Additional models and analyses are identified in Table S8 and the sections below.

**Factor structure**

In each cohort, one-factor and correlated two-factor confirmatory factor analysis (CFA) models were fitted to the four-category items using weighted least squares mean- and variance-adjusted (WLSMV) estimation [5,6] and theta parameterization [7]. The two-factor model followed Tibubos and colleagues [8], with items 1, 2, 6, 7, and 9 on the cognitive-affective factor and items 3, 4, 5, and 8 on the somatic factor; published assignments vary [9-11]. In the original cohort, models were compared on convergence, admissibility, standardized factor loadings, the correlation between factors, overall fit, and change in fit across models. Chi-square, CFI, TLI, RMSEA, and SRMR [12,13] were reported, with scaled and robust versions of chi-square, CFI, TLI, and RMSEA reported separately. Fit indices for the one-factor and correlated two-factor models in the replication cohort are reported in Table S3.

Conventional fit indices were summarized alongside evidence of misfit [14]. Conventional WLSMV cutoffs can under-detect misfit [15,16]. Interpretation of the fit indices considered absolute fit and the pattern of residual dependence.

**Reliability of the total score and general-factor saturation**

Reliability of the total score was estimated with omega total for the underlying continuous item responses from the one- and two-factor models [17]. General-factor saturation was assessed with an exploratory Schmid–Leiman decomposition [18]. Parallel analysis (common-factor, polychoric correlations, 500 simulated replications) [19] was conducted in each assessment group of the original cohort. Four observed common-factor eigenvalues exceeded the simulated means in the diagnosed group and three did so in the assessed comparison group. The Schmid–Leiman decomposition specified three group factors in both cohorts. A two-group-factor Schmid–Leiman solution requires equality constraints on the higher-order loadings for identification. No such constraints were imposed. The confirmatory two-factor model tests the published item assignment, whereas the exploratory decomposition estimates how much reliable variance a general factor carries without imposing an assignment; the Schmid–Leiman estimates were therefore used as the primary basis for judging general-factor saturation.

Omega total, omega hierarchical ($\omega H$), explained common variance (ECV), and the general factor's share of reliable total-score variance were reported descriptively [20-22]. The percentage of uncontaminated correlations (PUC) was .556 under the two-domain CFA assignment and .722 under the dominant-loading grouping of the Schmid–Leiman solution. Interfactor correlations were considered when assessing support for separate domain scores.

**Precision**

Unidimensional graded response models were fitted separately in each assessment group in both cohorts using marginal maximum likelihood with the expectation-maximization algorithm [1,23]. Local dependence was examined as described in the next subsection. Overall model fit was assessed with the limited-information C2 statistic and its associated RMSEA [24] and interpreted alongside item fit. Item fit was assessed with the S-$X^2$ statistic [25]. Items were flagged when the Benjamini–Hochberg–adjusted P value was <.05, with adjustment across the nine items within each assessment group and cohort. Item-level RMSEA was reported to describe the magnitude of misfit. Item discrimination and category thresholds were estimated in both cohorts, and item and test information were used to describe precision. Peak information was defined as the severity (θ) at which test information was highest on the evaluated grid. The conditional standard error of estimated severity was the inverse square root of test information [26].

Marginal sum-score reliability was estimated in both cohorts as the variance of the model-expected PHQ-9 total divided by the sum of that variance and the mean conditional variance of the observed total, integrating over a standard normal severity distribution [26]. In the original cohort, conditional reliability of the severity estimate was calculated as $1 - 1/I(\theta)$, where $I(\theta)$ is test information from the separate-group model. Conditional reliability was evaluated from $\theta = -4$ to 4 in steps of 0.10 to identify the range in which reliability was at least 0.80, corresponding to a conditional standard error of approximately 0.45 or less. Because each group's severity scale was standardized separately, the endpoints were also expressed as model-expected PHQ-9 totals from the test characteristic curve. In the original cohort, a Rasch rating-scale sensitivity analysis provided marginal person reliability and item infit and outfit statistics [27,28].

**Local dependence**

Local dependence was examined because residual associations between items can inflate test information and reliability estimates [29]. In each assessment group and cohort, Yen's Q3 [30] was computed from the group's graded response model using expected a posteriori severity estimates [31]. Because Q3 tends to be negative under local independence, adjusted Q3 was calculated by subtracting the group's mean Q3 across all 36 item pairs from each pair's Q3. Adjusted Q3 values above 0.20 were flagged descriptively in both cohorts [2]. In the original cohort, 500 data sets per assessment group were simulated under local independence from the fitted unidimensional graded response model. The same model specification was refitted to each simulated data set. The group-specific bootstrap critical value was defined as the 95th percentile of the maximum adjusted Q3 across simulations [2]. In the replication cohort, only the descriptive adjusted Q3 flag was applied. As supplementary checks in the original cohort, unadjusted Q3 values and residual correlations from the one-factor and correlated two-factor ordinal confirmatory factor analyses were examined against the COSMIN reference criteria of Q3 <.37 or residual correlations <.20 [14].

**Associations with anxiety and wellbeing**

Exploratory wellbeing ratings included the stress and emotional challenges item from the service's internally developed quality-of-life questionnaire and a separate overall wellbeing item. Higher scores indicated better wellbeing on both ratings. The internally developed wellbeing instrument is proprietary. Public English versions of the PHQ-9 and GAD-7 are available from the American Psychological Association (https://www.apa.org/depression-guideline/patient-health-questionnaire.pdf) and the Agency for Healthcare Research and Quality (https://integrationacademy.ahrq.gov/sites/default/files/2020-07/GAD-7.pdf), respectively.

Pearson and Spearman correlations of PHQ-9 total scores with GAD-7 total scores and each of the two wellbeing ratings were estimated from first available responses, separately by assessment group within each cohort. For Pearson correlations, absolute coefficients of 0.50 for GAD-7 totals and 0.30 for each wellbeing rating were used as descriptive benchmarks for magnitude [32]. Signed coefficients were reported to show the direction of each association.

To examine whether depression and anxiety were empirically distinguishable, joint ordinal confirmatory factor analyses of all 16 PHQ-9 and GAD-7 items were fitted separately within each assessment group and cohort. A single general distress factor and two correlated factors representing depression and anxiety were compared descriptively using model fit indices. The latent correlation between depression and anxiety was estimated from the two-factor model. The polychoric heterotrait–monotrait ratio (HTMT) [33] was calculated with 95% percentile intervals based on 2,000 attempted client-level bootstrap resamples within each assessment group and cohort [34]. An HTMT of 0.85 was used as a descriptive reference for the point estimate, and the upper limit of the 95% interval was compared with 0.90 [33,35].

**Group definition and response-category coding**

Measurement invariance, differential item functioning (DIF), and expected total-score differences were evaluated separately in the original and replication cohorts. In the graded response model (GRM) analyses [1,23], clients who were assessed but did not meet autism diagnostic criteria formed the reference group, and diagnosed clients formed the focal group. In the GRM, each threshold corresponds to the severity level at which the probability of endorsing a given response category or higher is 0.50 [1,23]. When few clients in a group endorse the upper categories, the corresponding thresholds can be estimated imprecisely, making group differences in item functioning harder to distinguish from sampling variability. To reduce reliance on sparse cells, adjacent response categories were pooled before model fitting whenever any group-by-category cell contained fewer than 10 clients, with at least two categories modeled per item. The category-pooling rule required a larger cell count than the minimum of five observations per response category used in a previous PHQ-9 DIF analysis.

In the original-cohort assessed comparison group, each of the highest two item 9 response categories had fewer than 10 clients. The two highest response categories were pooled in both original-cohort groups (modeled range 0–26). The standard replication-cohort models used all original categories (modeled range 0–27), and the common-coding anchor analysis pooled the highest two item 9 categories in both cohorts.

**Measurement invariance**

The correlated two-factor ordinal model was used to evaluate measurement invariance across assessment groups in each cohort. Models were fitted using WLSMV estimation and theta parameterization [7,36]. Under the Wu and Estabrook identification scheme [37], equality constraints were added sequentially for thresholds, loadings, latent-response intercepts, and residual variances. Configural fit was evaluated using CFI, RMSEA, and SRMR. Adjacent models were compared using scaled and shifted chi-square difference tests, with $P<.05$ indicating rejection of the added constraints [38]. The full sequence was fitted, and later tests were interpreted conditional on the constraints already imposed.

Changes in approximate fit were interpreted descriptively using benchmarks of a CFI decrease of ≥.010, an RMSEA increase of ≥.015, and an SRMR increase of ≥.030 at the loading step or ≥.010 at the intercept and residual-variance steps [3,39]. These benchmarks were developed for continuous indicators [3,39]. No SRMR benchmark was applied at the threshold step. The supplementary tables report the scaled and shifted chi-square difference statistics, degrees of freedom, P values, and unscaled chi-square statistics for the fitted models.

In the original cohort, exploratory follow-ups assessed joint equality of thresholds, loadings, and intercepts against the configural model. For comparisons with a rejected joint test, up to three constraints were released one at a time in descending order of univariate score-test $\chi^2$, stopping at $P\geq.05$. The Black-versus-White and multiracial-versus-White pairwise checks used the joint test of thresholds, loadings, and intercepts. The ethnicity comparison yielded $P=.103$ on its pairwise joint test and no constraints were released. In the replication cohort, the subgroup follow-ups used the adjacent intercept-equality test. When rejected, intercepts were freed sequentially by decreasing score-test $\chi^2$ with the threshold-and-loading model as comparator. Table S11 identifies which results come from omnibus or pairwise tests.

Regardless of the invariance test results, differential item functioning (DIF) and expected item- and total-score differences were evaluated using a unidimensional graded response model to examine the PHQ-9 as a single total score [40,41]. Expected-score differences were expressed in PHQ-9 points.

**DIF screening and anchor selection**

DIF was assessed by comparing item responses between groups at the same modeled depression severity. Anchor items placed the groups on a common scale by holding their discrimination and thresholds equal across groups. The reference group's latent mean and variance were fixed at 0 and 1, and both were estimated in the focal group. Anchor-based linking assumes that the anchor items function equivalently across groups [42-44].

Multigroup graded response models were fitted [1]. For each item, a likelihood-ratio test compared models that constrained or freed its discrimination and thresholds while retaining the other anchor constraints. Tests had 4 degrees of freedom for four-category items and 3 for three-category items. The initial anchor pool comprised items 1–8 for both primary cohort comparisons and the original-cohort subgroup screens, and items 1–9 for the replication cohort subgroup screens.

During purification [43], each candidate was tested against the other current anchors. Benjamini–Hochberg adjustment was applied within each round, with adjusted $P<.05$ flagging an item [45]. The flagged candidate with the smallest unadjusted P value was removed, with ties resolved by item number. Removed items stayed out of the pool and were free across groups in tests of other items. Purification stopped when no candidate was flagged. At three remaining candidates, the primary-cohort and original-subgroup procedures allowed one final removal if a candidate was flagged, whereas the replication cohort subgroup procedure stopped at three. All nine items were then tested in a separate final screen. Holm-adjusted P values were calculated for each round and the final screen as a sensitivity check [46].

For expected total-score estimation, three anchors were selected separately for each contrast and cohort [42,43]. Items qualified if they were unflagged throughout purification and in the final screen. Item 9 was excluded from final linking because its upper categories were sparsely endorsed in the original cohort. The three eligible items with the largest unadjusted final-screen P values were selected, with ties resolved by item number. These P values ranked candidates for the data-driven selection. DIF magnitude was evaluated using expected-score differences in PHQ-9 points. When fewer than three items qualified, the primary three-anchor estimate was unavailable. For men versus women, sensitivity models used item 1 alone, item 7 alone, and both together. For the primary assessment-group comparisons, sensitivity analyses examined all three-item anchor sets drawn from items 1–8.

**Expected total-score differences**

For each comparison, a unidimensional multigroup graded response model was fitted by marginal maximum likelihood. Dimensionality and model fit are summarized in Tables S4, S6, and S15. Anchor-item discriminations and thresholds were constrained to equality across groups, and the remaining item parameters were estimated separately by group [1]. The reference group’s latent mean and variance were fixed at 0 and 1, respectively, and both were estimated in the focal group. For the assessment-outcome comparisons, the diagnosed group was the focal group and the assessed comparison group was the reference group. Expected PHQ-9 totals were compared at the same modeled severity using the differential response function method [40,41,47]. Differences were evaluated at 61 equally spaced severity nodes from −6 to 6. Each node was weighted by the mean posterior probability across clients in both groups, calculated using each client's group-specific item parameters and latent distribution. The weights were normalized to sum to 1 across nodes. Each client contributed equally, so the weights reflected the relative group sizes in each cohort. The signed difference was the weighted average of the focal-minus-reference expected total across nodes. In the assessment-outcome comparisons, positive values indicated higher expected totals in the diagnosed group at the same modeled severity, on average. The unsigned difference was the weighted average of the absolute total-score difference across nodes. Opposing item differences could offset one another within a node. Taking the absolute value before averaging prevented cancellation across nodes. Parameter vectors were drawn from a multivariate normal distribution centered on the maximum likelihood estimates, with covariance estimated using the Oakes method [48]. Invalid parameter vectors were redrawn, with the redraw limit set to 100. Expected scores and node weights were recalculated for each accepted draw. Central 95% percentile intervals are used for the primary and selected sensitivity models. The unsigned bounds use the 95th percentile as a one-sided limit. Central 90% intervals are labeled for other sensitivity models. These intervals and bounds quantify parameter uncertainty conditional on the fitted model and selected anchors. Uncertainty from anchor selection and model specification falls outside their scope [42,43]. Selection runs used two batches of 4,000 draws in the original cohort and two batches of 1,000 in the replication cohort, which gives the totals in Table S15, Panel A (8,000 and 2,000). The central 95% intervals for the screening-selected models use the 2,000 draws retained for each model. The main severity curves show pointwise 95% intervals. When response categories were pooled, a worst-case envelope was calculated on the original 0–27 scale. At each severity node, the lower bound assigned each pooled category its lowest possible original score in the focal group and its highest in the reference group. The upper bound reversed those assignments. For example, when item 9 responses of 2 and 3 were pooled, the lower bound assigned that category a score of 2 in the diagnosed group and 3 in the reference group. For each draw, the lower and upper bounds were averaged across nodes using the same weights as the modeled-score estimates. The signed envelope used the percentile bounds of the draw-specific lower and upper bounds, with the interval level stated for the source run. For the unsigned upper bound, the larger absolute bound at each node was averaged using the same weights, and the 95th percentile of those draw-specific averages was reported.

**Item 9**

In addition to the nine-item DIF screen, item 9 (thoughts of death or self-harm) was examined separately because responses have been associated with subsequent suicide attempts and deaths [49] and because Robeson et al. reported loading noninvariance between autistic and nonautistic university students [50]. Because the upper categories were sparse, any endorsement (responses 1–3) was compared with no endorsement (response 0). The PHQ-8 score, the sum of items 1–8 (range 0–24), was used so that item 9 did not contribute to its own matching score [51]. Logistic regression models included assessment group, PHQ-8 score, and their interaction [52]. PHQ-8 scores were centered at 10, so the group coefficient represented the difference in log odds of endorsement at that score. The interaction assessed whether the between-group difference in log odds varied with PHQ-8 score. Because the PHQ-8 measures severity with error and can include items flagged for DIF, clients with the same observed score may differ in latent severity. The item 9 graded response estimates provided a complementary comparison at the same modeled latent severity. The primary estimates were obtained separately for the original and replication cohorts. Odds ratios and risk differences for diagnosed versus assessed comparison clients were estimated at PHQ-8 scores of 5, 10, and 15. The 95% odds-ratio intervals were Wald intervals based on the coefficient covariance matrix, and the 95% risk-difference intervals were obtained from multivariate normal coefficient draws [53]. Under a study-specific support rule, estimates were flagged for cautious interpretation if either group had fewer than five endorsers or five nonendorsers within two PHQ-8 points of the target score. In the combined-cohort analysis, a two-degree-of-freedom likelihood-ratio test jointly evaluated the group-by-cohort and group-by-PHQ-8-by-cohort terms. Combined risk differences were standardized to the observed cohort proportions. Supplementary ordinal CFAs estimated group-specific loadings of item 9 on the cognitive-affective factor with item thresholds constrained across groups. Item 9 responses of 2 and 3 were combined in both cohorts. Standardized loading differences were estimated with delta-method intervals. As a separate sensitivity analysis, the differential mean and covariance structure effect size (dMACS) was estimated using a one-factor robust maximum likelihood model that treated the original 0–3 responses as continuous [54]. Exploratory cumulative-logit analyses combined the cohorts and examined both four-category coding and coding with responses 2 and 3 combined. Proportional odds were examined using term-specific likelihood-ratio comparisons. Subsequent models allowed assessment group, PHQ-8 score, and their interaction to vary by threshold. Predicted category probabilities were checked over the observed score range within each group and cohort. Upper-threshold estimates were interpreted cautiously because of sparse responses. Table S18 reports the binary and ordinal analyses.

***Post hoc item 9 analyses***

Follow-up analyses compared raw endorsement at PHQ-8=5 and within PHQ-8 scores of 3–7, estimated the group-by-PHQ-8 interaction as a ratio of odds ratios per 5 PHQ-8 points, and compared the score-5 contrast between cohorts with a Wald test. Sensitivity models added quadratic and natural-spline PHQ-8 terms or used Firth penalized likelihood (logistf version 1.26.1, with penalized profile-likelihood intervals). Absolute and signed fitted differences in endorsement probability were averaged over the diagnosed PHQ-8 distribution at supported scores. McFadden $\Delta R^2$ values were reported descriptively. Item 9 loadings were estimated in ordinal two-factor models with item 9 responses 2 and 3 pooled and thresholds constrained across groups in both cohorts, and the cross-cohort contrast was computed from the two cohort-specific standardized differences. A descriptive follow-up (Panel G) summarized repeat item 9 endorsement and elapsed days among baseline endorsers with a second eligible paired submission in the same cohort window. These analyses were not part of the primary plan (Table S18).

**Comparisons within diagnosed clients**

Exploratory analyses evaluated measurement invariance, DIF, and expected total-score differences for seven pairwise comparisons among diagnosed clients in both cohorts. The comparisons were men versus women, gender-diverse clients versus women, age under 35 versus 35 or older, Black versus White clients, multiracial versus White clients, Hispanic or Latine versus non-Hispanic or Latine clients, and clients with versus without a recorded ADHD code. All seven comparisons included at least 100 clients per group. The larger group served as the reference, so positive differences indicate higher expected totals in the smaller group at the same modeled severity. Follow-up releases were based on pairwise tests and are reported in Table S11.

The omnibus gender model included women, men, and gender-diverse clients. The omnibus race model included White, Black, and multiracial clients. Race and ethnicity were self-reported on the clinical intake form, which included an “Other” response option. They were analyzed separately to examine whether PHQ-9 items and totals functioned differently across the recorded groups. The Asian, Native American, and Native Hawaiian groups each had fewer than 100 diagnosed clients and were excluded from the original-cohort race comparisons. Recorded categories and missing responses are summarized in Table S1. The ADHD comparison was restricted to clients with available treatment-plan records and was defined by recorded code status. Age was divided at 35 years, the median among diagnosed clients with recorded age, to permit the same multigroup analysis used for the other contrasts. There were 1,629 clients younger than 35 and 1,841 aged 35 or older. One client lacked a recorded age. The cutpoint was sample-specific and had no clinical interpretation. Each characteristic was examined separately, so DIF associated with one characteristic could reflect its association with another. Benjamini–Hochberg adjustment was applied to the nine item tests within each contrast. Error rates were not controlled across contrasts. Response-category coding is reported in Table S15. For men versus women, only items 1 and 7 met the anchor criteria, so a screening-selected three-anchor estimate was unavailable. Sensitivity estimates were obtained with each item as the sole anchor and, after inspection of those estimates, with both items together. Likelihood-ratio tests, AIC, and BIC were used descriptively to compare the two-anchor model with each single-anchor model [55,56]. Each likelihood-ratio comparison evaluated one anchor constraint conditional on retaining the other (Table S19).

**Sensitivity analyses**

Women accounted for 2,167 of 3,471 diagnosed clients (62.4%) and 350 of 484 assessed comparison clients (72.3%). To examine sensitivity to the difference in the proportion of women between assessment groups, the expected total-score difference was re-estimated among women using the original assessment-group anchors (items 4, 6, and 7), held fixed across analyses. The two nonoverlapping cohorts were also combined to estimate expected total-score differences with all four item 9 categories modeled. In the combined assessed comparison group, 27 clients selected category 2 and 22 selected category 3, meeting the 10-client minimum. Separate models used the anchor set selected by iterative purification in each cohort. The combined analysis included the participants from the cohort-specific analyses and was interpreted as a sensitivity check. To limit variation in therapy exposure before measurement, the ordinal invariance sequence was repeated among clients whose first PHQ-9 was linked to therapy intake (2,219 diagnosed and 290 assessed comparison). To assess sensitivity to classification based on reports issued after baseline, the invariance sequence and expected total-score analysis were repeated among clients whose assessment report was dated on or before the first administration (2,047 diagnosed and 225 assessed comparison). The remaining 259 assessed comparison clients had reports dated after baseline. Item 9 was coded as any endorsement versus none in both restricted CFA samples. The assessment-timing analysis also combined item 8 responses of 2 and 3 (Tables S11, S15, and S20). The ordinal invariance sequence was repeated using unweighted least squares with mean- and variance-adjusted test statistics (ULSMV) [57] to assess sensitivity to the diagonal fitting weights used in the primary WLSMV models. Full-information graded response models provided complementary estimates of DIF and expected-score differences. Additional sensitivity analyses included excluding records flagged by response-quality screens and separately excluding all-zero profiles, balanced subsampling to examine unequal group sizes [58], label permutation to obtain an empirical reference for invariance tests [59], and simulations to assess detection of known item differences under assessment-label error. Broader service-group definitions examined sensitivity to the inclusion of clients with prior outside assessments, incomplete assessments, or no in-service assessment record. A domain-mean model provided a supplementary assessment-group comparison.

***Exploratory treatment-plan code analysis***

PHQ-9 scores at the first eligible check-in were compared with the presence of any F32- or F33-series code at any position on available treatment plans. The comparison included clients with treatment-plan records. Estimates included group means, Hedges g, the area under the receiver operating characteristic curve (with DeLong 95% CIs), and sensitivity and specificity for a total of 10 or higher relative to recorded codes. Comparisons of areas under the curve used unadjusted two-sided tests. A sensitivity analysis added F34.1. The extraction included remission codes and did not impose a timing restriction on treatment plans (Table S24).

**Study setting details**

Recorded client addresses represented 28 states in the original analytic cohort and 35 in the replication cohort. The analytic extracts did not include payer information. Participation in the Prosper Health research program is voluntary. The organization maintains clinical and research advisory structures that include autism specialists and autistic advocates or community members who provide input on research priorities and designs. Clinicians used explicit, concrete language and adjusted the pace and delivery of therapy to each client's sensory needs and communication preferences. Gilmore et al [60] describe related adaptations for mental healthcare with autistic adults.

##### Conventional bifactor, bifactor S-1, factorability, and parallel analysis

A conventional bifactor model with a general depression factor and two domain-specific factors was also fitted in each original-cohort assessment group [61]. Estimation used 100 random starts and one standard start. The converged solution with the lowest WLSMV fitting-function value was selected, and its admissibility was assessed separately. Two bifactor S-1 models were fitted, with each symptom domain serving in turn as the reference that defined the general factor [62]. Factorability was assessed with the Kaiser–Meyer–Olkin (KMO) measure and Bartlett's test [63,64]. Common-factor parallel analysis used polychoric correlations and 500 simulated replications [19]. Original-cohort model comparisons considered convergence, admissibility, factor loadings, correlations between factors, overall fit, and changes in fit across models. Because RMSEA from ordinal WLSMV models depends on the number of indicators, the estimator, the scaling correction, and the degrees of freedom [13,15,16], it was interpreted alongside the other fit indices. Each model was evaluated against the COSMIN criteria to determine whether it met at least one fit criterion (CFI or TLI >.95, RMSEA <.06, or SRMR <.08) [14]. The conventional bifactor and bifactor S-1 models were original-cohort sensitivity analyses. Their results, including instability across random starts in the diagnosed group, are reported in Tables S3–S4.

#### Extended results

##### Exploratory analyses within diagnosed clients: samples and invariance

Original-cohort exclusions were 424 gender-diverse clients, 22 with unclassified gender, and 103 with missing gender for men versus women; 755 men and the same 125 for gender-diverse versus women; 1 client with missing age; 126 clients in racial groups below the 100-client minimum, 135 who preferred not to state race, 120 who self-described race only, 103 with missing race, and the other focal race group (213 multiracial clients or 331 Black clients); 144 with missing ethnicity; and 520 without an ADHD-code record. Replication cohort exclusions were 826 gender-diverse, 45 unclassified, and 32 with missing gender for men versus women; 1,557 men plus the same 77 for gender-diverse versus women; 2 with missing age; 1,290 excluded from Black versus White and 1,713 from multiracial versus White; 203 with missing ethnicity; and 1,651 without an ADHD-code record. Table S1 gives the replication cohort race-category breakdowns and the group counts.

In original-cohort pairwise comparisons, scaled and shifted $\chi^2$ difference tests of the fully constrained model against the configural model rejected equality for men versus women ($\chi^2(22)=71.80$, $P<.001$), gender-diverse versus women ($\chi^2(22)=54.42$, $P<.001$), age ($\chi^2(23)=51.98$, $P<.001$), Black versus White ($\chi^2(21)=35.27$, $P=.026$), and ADHD-code status ($\chi^2(22)=49.51$, $P<.001$). The men-versus-women model released the psychomotor loading, fatigue threshold 3, and concentration threshold 1; the final test remained rejected, $\chi^2(19)=35.82$, $P=.011$. Gender-diverse versus women released the appetite loading, first concentration threshold, and third fatigue threshold; after release, $\chi^2(19)=25.21$, $P=.154$. Age released the item 9 loading; after release, $\chi^2(22)=31.96$, $P=.078$. Black versus White released the first appetite threshold; after release, $\chi^2(20)=29.04$, $P=.087$. ADHD-code status released the first concentration threshold and concentration loading; after release, $\chi^2(20)=22.11$, $P=.335$. The multiracial-versus-White comparison required no release ($\chi^2(21)=16.42$, $P=.746$). The fully constrained-versus-configural ethnicity test was not rejected ($\chi^2(22)=30.68$, $P=.103$), so no partial-release sequence was fitted for that contrast.

#### Additional results

In the original diagnosed group, anhedonia and depressed mood had the largest discrimination estimates (2.70 and 2.92) and together contributed 44% of information at clients' expected a posteriori severity; item 9 contributed 4%.

#### Sensitivity analyses

##### *Replication cohort partial scalar invariance*

The full intercept test in the replication cohort gave P=.003. After releasing the intercepts for item 2 (depressed mood) and then item 1 (anhedonia), the comparison with the threshold-and-loading model gave $\Delta\chi^2(5)=10.03$, P=.074. For the replication cohort pairwise comparisons, partial models fitted after the following releases (final P): men versus women, items 8, 7, and 4 (.39); gender-diverse versus women, item 9 (.42); age, items 9, 4, and 6 (.076); Black versus White, item 6 (.086); Hispanic or Latine versus not, item 4 (.0503); ADHD code, item 7 (.76). All releases were exploratory and used the threshold-and-loading model as the comparator. For the Hispanic or Latine comparison, the scaled-shifted difference test after releasing the fatigue intercept was $\Delta\chi^2(6)=12.57$, P=.0503. The remaining intercept constraints were not rejected at the P<.05 threshold.

##### *Fit changes under added constraints*

Unscaled $\chi^2$ increased at every adjacent step for the primary comparisons. The original-cohort Black-versus-White and multiracial-versus-White intercept comparisons produced nesting warnings, so those two difference tests are not interpreted.

##### *Power check*

In 500 replication cohort subsamples drawn at the original group sizes (484 reference and 3,471 focal clients), the intercept equality test rejected in 193 (38.6%; exact binomial 95% CI

34.3% to 43.0%). The rejection frequency describes the replication cohort data at the original sample sizes; it cannot identify the causes of the cohort difference.

***Expected totals at the screening cutoff***

At the severity where the reference group's expected total was 10, the diagnosed group's expected total was 10.18 in the original cohort and 10.03 in the replication cohort under the screening-selected anchors. Both these estimates and the ranges across the 56 anchor sets used cohort-specific coding (original range 0–26, replication range 0–27). Across the 56 sets, the ranges were 9.57 to 10.51 and 9.52 to 10.53.

***Descriptive error band for two scores***

With standard errors of measurement of 2.11 to 2.23 points (Table 1 of the main manuscript), $1.96 \times \sqrt{2} \times \text{SEM}$ was 5.8 to 6.2 points (reported in the main text as the 95% minimal detectable change implied by single-administration reliability) under an assumption of independent errors of equal variance. The error band was based on internal consistency at a single administration. SEMs based on graded response model sum-score reliability were similar (2.06 to 2.21 points; $1.96 \times \sqrt{2} \times \text{SEM}$, 5.7 to 6.1 points).

***Interval endpoint precision***

An endpoint precision check used 8,000 parameter draws (seed 20262160) at the original-cohort link defined by items 3, 4, and 8. The lower endpoints from the same draws were −0.749 at 90% and −0.808 at 95%. The Monte Carlo uncertainty interval for the 95% lower endpoint was −0.818 to −0.797.

***Factor-mean difference***

These contrasts use the assessed comparison group as the reference, whereas Table S23 uses the diagnosed group as the reference. The diagnosed-minus-comparison difference in factor means, in reference-group SD units, was 0.131 on the cognitive-affective factor and 0.207 on the somatic factor in the original cohort (somatic minus cognitive-affective 0.076, SE 0.045, 95% CI −0.013 to 0.165; Wald $\chi^2(1)=2.82$, P=.093). In the replication cohort the fully constrained model gave 0.072 and 0.202 (difference 0.130, 95% CI 0.069 to 0.191; P<.001), but its intercept step was rejected; the partial scalar model gave 0.146 and 0.202 (difference 0.056, 95% CI −0.018 to 0.129; P=.139). The partial scalar model is used for interpretation.

***Local-dependence sensitivity***

Seven-item totals were used for item-deletion checks (score range 0–20 in the original cohort, where item 9 responses 2 and 3 were pooled, and 0–21 in the replication cohort). In the original cohort, dropping items 4 and 8 gave −0.027 points (95% CI −0.248 to 0.163), and dropping items 3 and 7 gave −0.029 points (95% CI −0.227 to 0.144). In the replication cohort, dropping items 4 and 8 gave −0.135 points (95% CI −0.284 to 0.002). The replication-cohort model dropping items 3 and 7 was not fitted because fewer than three eligible anchor items remained. Where a dropped item was an anchor, the model used a different link from the anchor set selected by iterative purification (original cohort: anchors 6, 7, and 3 after dropping items 4 and 8, and 4, 6, and 5 after dropping items 3 and 7; replication cohort: anchors 3, 6, and 7). Intervals are central 95% percentile intervals from 2,000 parameter draws per fit. The two original-cohort estimates were nearly identical, so the result did not depend on which member of the dependent pair was retained. These estimates are on seven-item scales and are not directly comparable with the nine-item anchor-set range.

***Latent means across anchor sets***

The diagnosed group’s estimated mean was positive under all 56 anchor sets in each cohort. Estimated differences ranged from 0.118 to 0.263 reference-group SDs in the original cohort and from 0.046 to 0.250 in replication. Model-specific 95% Wald intervals excluded zero for 56 of 56 original-cohort sets and 49 of 56 replication-cohort sets. The estimates under the anchor sets selected by iterative purification were 0.175 SD (95% CI 0.062–0.289) in the original cohort and 0.144 SD (95% CI 0.066–0.223) in replication.

***Dominant-loading PUC***

The dominant-loading assignment of the three-group-factor Schmid–Leiman solution was identical in all four groups (items 1, 2, 6, and 9; items 3, 4, and 5; items 7 and 8), with PUC=.722 and 26 cross-domain pairs. The exploratory decomposition retains cross-loadings, so the dominant-loading assignment is a descriptive proxy.

***Treatment-plan code comparison***

The AUC difference between assessment groups (assessed comparison minus diagnosed) was 0.022 in the original cohort (unadjusted two-sided P=.475) and −0.048 in the replication cohort (P=.039). With F34.1 added to the code definition, AUCs were 0.628 (diagnosed) and 0.651 (assessed comparison) in the original cohort and 0.641 and 0.605 in the replication cohort (differences 0.022, P=.475; −0.037, P=.109). Hedges g for PHQ-9 totals with versus without an F32/F33 code was 0.45 (95% CI 0.37 to 0.53) in original diagnosed and 0.55 (0.34 to 0.76) in original assessed comparison clients, and 0.53 (0.47 to 0.59) and 0.35 (0.19 to 0.50) in the replication cohort.

***Two-factor residual-covariance sensitivity***

Freeing the sleep–fatigue and concentration–psychomotor residual covariances changed fit as follows (standard model to model with the two covariances): original assessed comparison (n=484): scaled CFI 0.971 to 0.976, scaled RMSEA 0.097 to 0.091, SRMR 0.058 to 0.052; original diagnosed (n=3,471): scaled CFI 0.970 to 0.981, scaled RMSEA 0.095 to 0.078, SRMR

0.054 to 0.042; replication cohort assessed comparison (n=1,194): scaled CFI 0.947 to 0.967, scaled RMSEA 0.113 to 0.092, SRMR 0.070 to 0.055; replication cohort diagnosed (n=7,406): scaled CFI 0.962 to 0.975, scaled RMSEA 0.100 to 0.085, SRMR 0.057 to 0.046. Identical category coding was used with and without the covariances. Configural scaled RMSEA fell by .016 in both cohorts, which met the .010 threshold for rerunning the invariance sequence; the intercept-equality decision was unchanged (not rejected in the original cohort; rejected in the replication cohort).

***Severity weighting***

Re-averaging the fixed-anchor expected-score difference over pooled, reference-group, and focal-group severity distributions gave signed estimates of pooled sample size weighted 0.077, reference 0.069, focal 0.078 points in the original cohort (anchors 4, 6, and 7) and pooled sample size weighted 0.002, reference 0.006, focal 0.002 points in the replication cohort (anchors 3, 6, and 7).

***Fixed original anchors in the replication cohort***

With the original anchors (items 4, 6, and 7) and item 9 pooled in both cohorts, the replication cohort signed difference was pooled sample size weighted −0.019, reference −0.004, focal −0.022 points.

***Retest analysis***

Among clients in the two assessment-outcome groups, only one original-cohort pair (diagnosed clients) and no replication-cohort pairs had first and second administrations 5 to 14 days apart, so no ICC, retest standard error, or retest-based change threshold could be estimated. The eligible administration exports included 10,766 records across 5,077 original-cohort clients and 21,759 records across 10,579 replication-cohort clients. Only the first eligible check-in per client entered the main analyses.
